# Association of Multiple Trait Polygenic Risk Score with Obesity and Cardiometabolic Diseases in Korean population

**DOI:** 10.1101/2025.04.13.25325699

**Authors:** Jinyeon Jo, Nayoung Ha, Yunmi Ji, Ahra Do, Je Hyun Seo, Bumjo Oh, Sungkyoung Choi, Eun Kyung Choe, Woojoo Lee, Jang Won Son, Sungho Won

## Abstract

We conducted a comprehensive genetic investigation of obesity in a cohort of 93,673 Korean individuals, categorized by both body mass index and waist circumference using Korean-specific and international criteria. To explore the genetic architecture of obesity and its comorbidities, we performed genome-wide association studies and constructed polygenic risk scores (PRSs) using both conventional single trait and advanced multiple-trait models, including the PRSsum approach.

Our analyses identified genome-wide significant loci and demonstrated higher heritability for general obesity than abdominal obesity, and for moderate compared to severe obesity. Notably, East Asian populations showed stronger genetic correlations between abdominal obesity and obesity-related diseases. Both single trait and multiple trait PRSs stratified individuals by risk, with low PRS individuals exhibiting reduced risk for obesity, hypertension, and type 2 diabetes, while high PRS individuals displayed elevated risk, particularly under the multiple trait model.

Additionally, interaction and mediation analyses revealed distinct genetic pathways through which obesity contributes to disease development. Collectively, our findings uncover key loci and shared genetic mechanisms linking obesity and its comorbidities in the Korean population. These insights highlight the value of multiple trait PRS models and underscore the importance of ancestry-specific genetic research for addressing the obesity epidemic.

## Introduction

Obesity remains a major global health challenge, with its prevalence steadily increasing over the past decades. According to the World Obesity Atlas 2023, approximately 38% of the global population is classified as overweight or obese [1]. This growing trend substantially heightens the risk of various chronic conditions, including type 2 diabetes (T2D), cardiovascular diseases (CVD), hypertension (HT), dyslipidemia, and certain cancers, thereby exacerbating the global healthcare burden [2−8].

The clinical presentation and health impact of obesity vary considerably across ethnic groups. In particular, East Asian (EAS) populations tend to develop obesity-related cardiometabolic disorders at lower body mass index (BMI) thresholds than individuals of European ancestry [9, 10]. Recognizing these population-specific differences, the Asia-Pacific World Health Organization (WHO) and the Korean Society for the Study of Obesity have proposed lower BMI cutoffs for defining obesity in EAS populations [11−14]. Moreover, EAS individuals often exhibit discordant obesity metrics, such as low BMI but relatively higher waist circumference (WC) [15], along with a higher prevalence of T2D [16]. These unique patterns underscore the need for population-specific approach to obesity research, especially in contrast to studies focused on non-Hispanic White (NHW) populations from European ancestry.

Although genome-wide association studies (GWASs) have identified numerous single nucleotide polymorphisms (SNPs) associated with obesity-related indicators [17−23], most of these studies have concentrated on NHW populations and primarily focused on BMI [24−28]. Consequently, their applicability to EAS population remain limited [29−32]. In addition, existing generic risk prediction models, typically based on single trait polygenic risk scores (PRS), often fail to fully capture the complex interrelated genetic structures connecting obesity with its related disorders.

In this study, we aim to address these gaps by investigating shared genetic components underlying obesity and obesity-related diseases including cardiometabolic disorders using a large-scale Korean cohort. Unlike prior Korean research that focused on genetic heterogeneity between obesity subtypes stratified by metabolic health status [33], our research applies advanced methodologies, notably employing multiple trait PRS models, including the integrative PRSsum approach. This framework enables us to identify shared genetic contributors to obesity and its comorbidities while improving risk prediction accuracy. By leveraging multiple trait PRS strategies tailored to the EAS population, our findings offer actionable insights for developing precise, population-specific preventive strategies to combat the growing obesity epidemic.

## Results

### Demographic characteristics of research participants

Our discovery dataset comprised 85,947 Korean individuals, consisting of 51,317 females and 34,630 males. **Table 1** summarizes demographic characteristics for this dataset, including the proportions of individuals classified as obese groups. General and abdominal obesity were assessed based on BMI- and WC-based criteria, respectively, as described in the Materials and methods section. Specifically, BMI-based classifications included BMI25 (≥25 kg/m²), BMI30 (≥30 kg/m²), and BMI40 (≥40 kg/m²), while WC-based classifications included WC1 (≥85 cm for females / 90 cm for males) and WC2 (≥95 cm for females / 100 cm for males). As shown in Table 1A, 33.29% of participants were classified as obese and 2.96% as severely obese based on BMI. Similarly, Table 1B shows that 25.49% exhibited abdominal obesity, with 3.68% classified as severely obese based on WC.

**Table 1.**
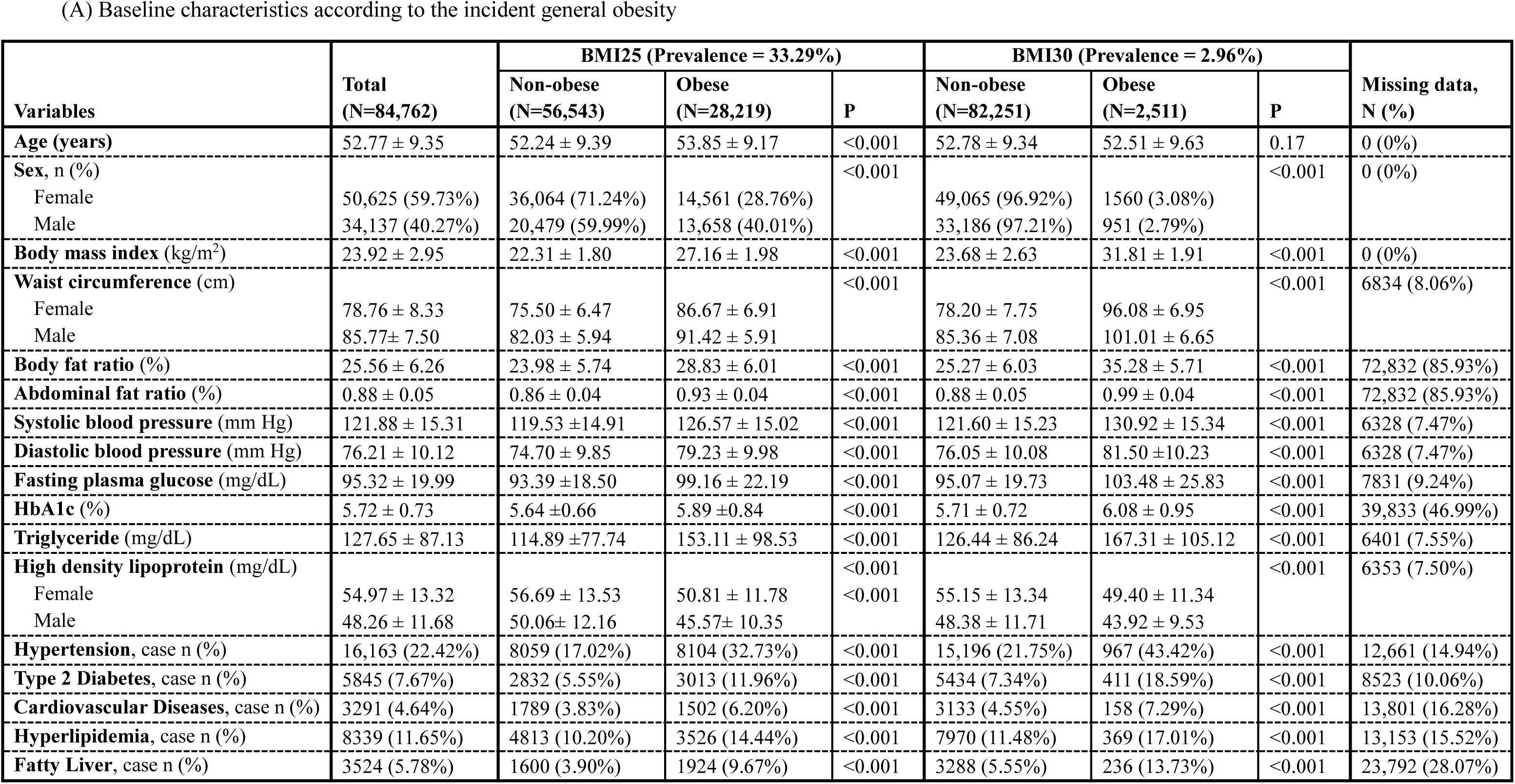
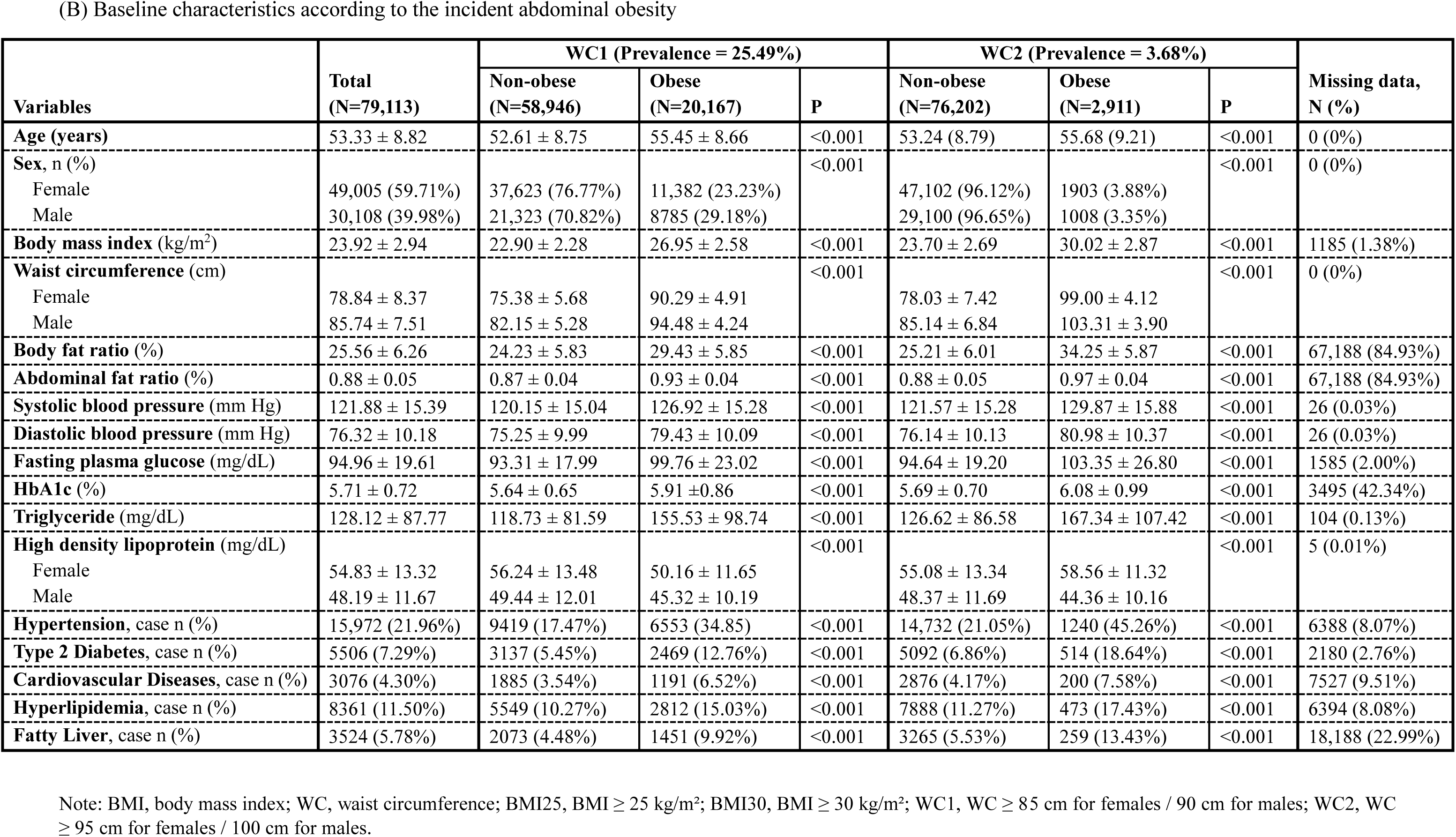
Baseline characteristics according to the incident obesity on discovery dataset.

(A) Baseline characteristics according to the incident general obesity
| Variables | Total<br>(N=84,762) | BMI25 (Prevalence = 33.29%) |  |  | BMI30 (Prevalence = 2.96%) |  |  | Missing data,<br>N (%) |
| --- | --- | --- | --- | --- | --- | --- | --- | --- |
|  |  | Non-obese<br>(N=56,543) | Obese<br>(N=28,219) | P | Non-obese<br>(N=82,251) | Obese<br>(N=2,511) | P |  |
| **Age (years)** | 52.77 ± 9.35 | 52.24 ± 9.39 | 53.85 ± 9.17 | <0.001 | 52.78 ± 9.34 | 52.51 ± 9.63 | 0.17 | 0 (0%) |
| **Sex, n (%)** |  |  |  | <0.001 |  |  | <0.001 | 0 (0%) |
| Female | 50,625 (59.73%) | 36,064 (71.24%) | 14,561 (28.76%) |  | 49,065 (96.92%) | 1560 (3.08%) |  |  |
| Male | 34,137 (40.27%) | 20,479 (59.99%) | 13,658 (40.01%) |  | 33,186 (97.21%) | 951 (2.79%) |  |  |
| **Body mass index (kg/m<sup>2</sup>)** | 23.92 ± 2.95 | 22.31 ± 1.80 | 27.16 ± 1.98 | <0.001 | 23.68 ± 2.63 | 31.81 ± 1.91 | <0.001 | 0 (0%) |
| **Waist circumference (cm)** |  |  |  | <0.001 |  |  | <0.001 | 6834 (8.06%) |
| Female | 78.76 ± 8.33 | 75.50 ± 6.47 | 86.67 ± 6.91 |  | 78.20 ± 7.75 | 96.08 ± 6.95 |  |  |
| Male | 85.77 ± 7.50 | 82.03 ± 5.94 | 91.42 ± 5.91 |  | 85.36 ± 7.08 | 101.01 ± 6.65 |  |  |
| **Body fat ratio (%)** | 25.56 ± 6.26 | 23.98 ± 5.74 | 28.83 ± 6.01 | <0.001 | 25.27 ± 6.03 | 35.28 ± 5.71 | <0.001 | 72,832 (85.93%) |
| **Abdominal fat ratio (%)** | 0.88 ± 0.05 | 0.86 ± 0.04 | 0.93 ± 0.04 | <0.001 | 0.88 ± 0.05 | 0.99 ± 0.04 | <0.001 | 72,832 (85.93%) |
| **Systolic blood pressure (mm Hg)** | 121.88 ± 15.31 | 119.53 ± 14.91 | 126.57 ± 15.02 | <0.001 | 121.60 ± 15.23 | 130.92 ± 15.34 | <0.001 | 6328 (7.47%) |
| **Diastolic blood pressure (mm Hg)** | 76.21 ± 10.12 | 74.70 ± 9.85 | 79.23 ± 9.98 | <0.001 | 76.05 ± 10.08 | 81.50 ± 10.23 | <0.001 | 6328 (7.47%) |
| **Fasting plasma glucose (mg/dL)** | 95.32 ± 19.99 | 93.39 ± 18.50 | 99.16 ± 22.19 | <0.001 | 95.07 ± 19.73 | 103.48 ± 25.83 | <0.001 | 7831 (9.24%) |
| **HbA1c (%)** | 5.72 ± 0.73 | 5.64 ± 0.66 | 5.89 ± 0.84 | <0.001 | 5.71 ± 0.72 | 6.08 ± 0.95 | <0.001 | 39,833 (46.99%) |
| **Triglyceride (mg/dL)** | 127.65 ± 87.13 | 114.89 ± 77.74 | 153.11 ± 98.53 | <0.001 | 126.44 ± 86.24 | 167.31 ± 105.12 | <0.001 | 6401 (7.55%) |
| **High density lipoprotein (mg/dL)** |  |  |  | <0.001 |  |  | <0.001 | 6353 (7.50%) |
| Female | 54.97 ± 13.32 | 56.69 ± 13.53 | 50.81 ± 11.78 | <0.001 | 55.15 ± 13.34 | 49.40 ± 11.34 |  |  |
| Male | 48.26 ± 11.68 | 50.06 ± 12.16 | 45.57 ± 10.35 |  | 48.38 ± 11.71 | 43.92 ± 9.53 |  |  |
| **Hypertension, case n (%)** | 16,163 (22.42%) | 8059 (17.02%) | 8104 (32.73%) | <0.001 | 15,196 (21.75%) | 967 (43.42%) | <0.001 | 12,661 (14.94%) |
| **Type 2 Diabetes, case n (%)** | 5845 (7.67%) | 2832 (5.55%) | 3013 (11.96%) | <0.001 | 5434 (7.34%) | 411 (18.59%) | <0.001 | 8523 (10.06%) |
| **Cardiovascular Diseases, case n (%)** | 3291 (4.64%) | 1789 (3.83%) | 1502 (6.20%) | <0.001 | 3133 (4.55%) | 158 (7.29%) | <0.001 | 13,801 (16.28%) |
| **Hyperlipidemia, case n (%)** | 8339 (11.65%) | 4813 (10.20%) | 3526 (14.44%) | <0.001 | 7970 (11.48%) | 369 (17.01%) | <0.001 | 13,153 (15.52%) |
| **Fatty Liver, case n (%)** | 3524 (5.78%) | 1600 (3.90%) | 1924 (9.67%) | <0.001 | 3288 (5.55%) | 236 (13.73%) | <0.001 | 23,792 (28.07%) |

(B) Baseline characteristics according to the incident abdominal obesity
| Variables | Total<br>(N=79,113) | WC1 (Prevalence = 25.49%) |  |  | WC2 (Prevalence = 3.68%) |  |  | Missing data,<br>N (%) |
| --- | --- | --- | --- | --- | --- | --- | --- | --- |
|  |  | Non-obese<br>(N=58,946) | Obese<br>(N=20,167) | P | Non-obese<br>(N=76,202) | Obese<br>(N=2,911) | P |  |
| **Age (years)** | 53.33 ± 8.82 | 52.61 ± 8.75 | 55.45 ± 8.66 | <0.001 | 53.24 (8.79) | 55.68 (9.21) | <0.001 | 0 (0%) |
| **Sex, n (%)** |  |  |  | <0.001 |  |  | <0.001 | 0 (0%) |
| Female | 49,005 (59.71%) | 37,623 (76.77%) | 11,382 (23.23%) |  | 47,102 (96.12%) | 1903 (3.88%) |  |  |
| Male | 30,108 (39.98%) | 21,323 (70.82%) | 8785 (29.18%) |  | 29,100 (96.65%) | 1008 (3.35%) |  |  |
| **Body mass index (kg/m<sup>2</sup>)** | 23.92 ± 2.94 | 22.90 ± 2.28 | 26.95 ± 2.58 | <0.001 | 23.70 ± 2.69 | 30.02 ± 2.87 | <0.001 | 1185 (1.38%) |
| **Waist circumference (cm)** |  |  |  | <0.001 |  |  | <0.001 | 0 (0%) |
| Female | 78.84 ± 8.37 | 75.38 ± 5.68 | 90.29 ± 4.91 |  | 78.03 ± 7.42 | 99.00 ± 4.12 |  |  |
| Male | 85.74 ± 7.51 | 82.15 ± 5.28 | 94.48 ± 4.24 |  | 85.14 ± 6.84 | 103.31 ± 3.90 |  |  |
| **Body fat ratio (%)** | 25.56 ± 6.26 | 24.23 ± 5.83 | 29.43 ± 5.85 | <0.001 | 25.21 ± 6.01 | 34.25 ± 5.87 | <0.001 | 67,188 (84.93%) |
| **Abdominal fat ratio (%)** | 0.88 ± 0.05 | 0.87 ± 0.04 | 0.93 ± 0.04 | <0.001 | 0.88 ± 0.05 | 0.97 ± 0.04 | <0.001 | 67,188 (84.93%) |
| **Systolic blood pressure (mm Hg)** | 121.88 ± 15.39 | 120.15 ± 15.04 | 126.92 ± 15.28 | <0.001 | 121.57 ± 15.28 | 129.87 ± 15.88 | <0.001 | 26 (0.03%) |
| **Diastolic blood pressure (mm Hg)** | 76.32 ± 10.18 | 75.25 ± 9.99 | 79.43 ± 10.09 | <0.001 | 76.14 ± 10.13 | 80.98 ± 10.37 | <0.001 | 26 (0.03%) |
| **Fasting plasma glucose (mg/dL)** | 94.96 ± 19.61 | 93.31 ± 17.99 | 99.76 ± 23.02 | <0.001 | 94.64 ± 19.20 | 103.35 ± 26.80 | <0.001 | 1585 (2.00%) |
| **HbA1c (%)** | 5.71 ± 0.72 | 5.64 ± 0.65 | 5.91 ± 0.86 | <0.001 | 5.69 ± 0.70 | 6.08 ± 0.99 | <0.001 | 3495 (42.34%) |
| **Triglyceride (mg/dL)** | 128.12 ± 87.77 | 118.73 ± 81.59 | 155.53 ± 98.74 | <0.001 | 126.62 ± 86.58 | 167.34 ± 107.42 | <0.001 | 104 (0.13%) |
| **High density lipoprotein (mg/dL)** |  |  |  | <0.001 |  |  | <0.001 | 5 (0.01%) |
| Female | 54.83 ± 13.32 | 56.24 ± 13.48 | 50.16 ± 11.65 |  | 55.08 ± 13.34 | 58.56 ± 11.32 |  |  |
| Male | 48.19 ± 11.67 | 49.44 ± 12.01 | 45.32 ± 10.19 |  | 48.37 ± 11.69 | 44.36 ± 10.16 |  |  |
| **Hypertension, case n (%)** | 15,972 (21.96%) | 9419 (17.47%) | 6553 (34.85) | <0.001 | 14,732 (21.05%) | 1240 (45.26%) | <0.001 | 6388 (8.07%) |
| **Type 2 Diabetes, case n (%)** | 5506 (7.29%) | 3137 (5.45%) | 2469 (12.76%) | <0.001 | 5092 (6.86%) | 514 (18.64%) | <0.001 | 2180 (2.76%) |
| **Cardiovascular Diseases, case n (%)** | 3076 (4.30%) | 1885 (3.54%) | 1191 (6.52%) | <0.001 | 2876 (4.17%) | 200 (7.58%) | <0.001 | 7527 (9.51%) |
| **Hyperlipidemia, case n (%)** | 8361 (11.50%) | 5549 (10.27%) | 2812 (15.03%) | <0.001 | 7888 (11.27%) | 473 (17.43%) | <0.001 | 6394 (8.08%) |
| **Fatty Liver, case n (%)** | 3524 (5.78%) | 2073 (4.48%) | 1451 (9.92%) | <0.001 | 3265 (5.53%) | 259 (13.43%) | <0.001 | 18,188 (22.99%) |

Note: BMI, body mass index; WC, waist circumference; BMI25, BMI ≥ 25 kg/m<sup>2</sup>; BMI30, BMI ≥ 30 kg/m<sup>2</sup>; WC1, WC ≥ 85 cm for females / 90 cm for males; WC2, WC ≥ 95 cm for females / 100 cm for males.

Compared to the non-obese, obese individuals tended to be older, and the proportion of males with moderate obesity was notably higher. The obese group also displayed elevated levels in anthropometric measures (BMI, WC, body fat ratio, and abdominal fat ratio), as well as less favorable biochemical profiles, including elevated blood pressures of systolic blood pressure (SBP)/diastolic blood pressure (DBP), increased blood sugar (BS) of fasting plasma glucose (FPG), and unhealthy blood lipids of triglyceride (TG)/high density lipoprotein (HDL). Moreover, a higher prevalence of obesity-related diseases was observed in this group. These trends were consistently observed across the replication datasets of Korean, Chinese and NHW populations, namely REP1_Kor_, REP2_Chi_, and REP3_NHW_, as presented in Table S1, Table S2, and Table S3. An overview of the study and analysis framework is provided in **Figure 1**.

**Figure 1.**
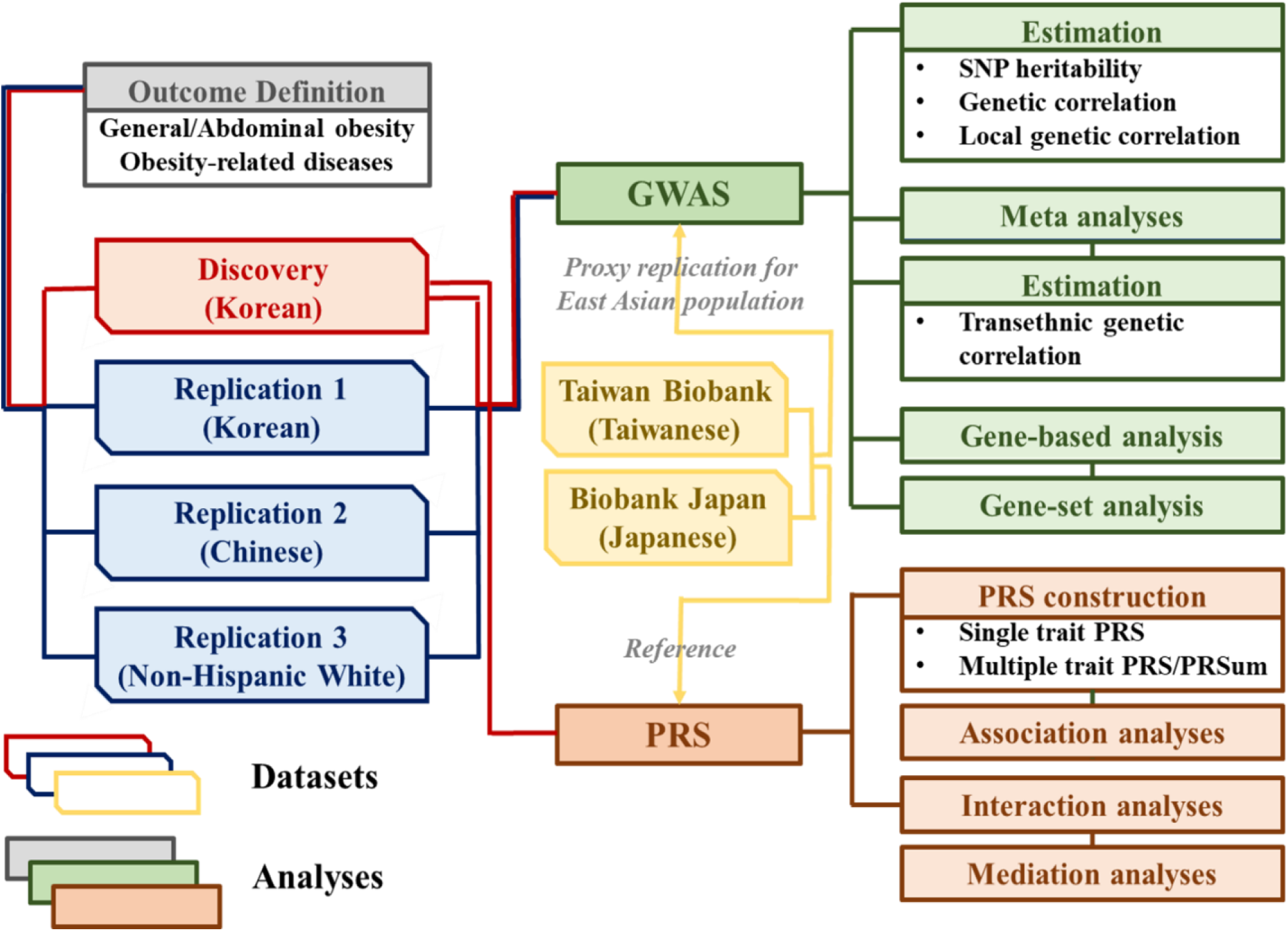
Analyses scheme This figure outlines the analysis flow including genome-wide association study (GWAS) and polygenic risk score (PRS) construction in the discovery and replication datasets.

### Genome-wide association studies, meta analyses and correlation analyses

We conducted GWAS for obesity using the discovery datasets and three replication datasets, with Manhattan plots shown in **Figure 2** and Figure S1, Figure S2, and Figure S3. From GWAS on general and abdominal obesity, we identified 22 genome-wide significant SNPs, of which eleven were replicated in REP1_Kor_, REP2_Chi_, or REP3_NHW_, presented in **Table 2**. Additionally, we included P values from Biobank Japan (BBJ) and Taiwan Biobank (TWB) as proxy replications to assess generalizability to other EAS populations, despite differences in outcome definitions. Most of SNPs still met genome-wide significance thresholds. Notably, all 22 genome-wide significant SNPs had been previously reported as associated with obesity-related indicators such as BMI or WC in prior studies, including those on European ancestry population.

**Figure 2.**
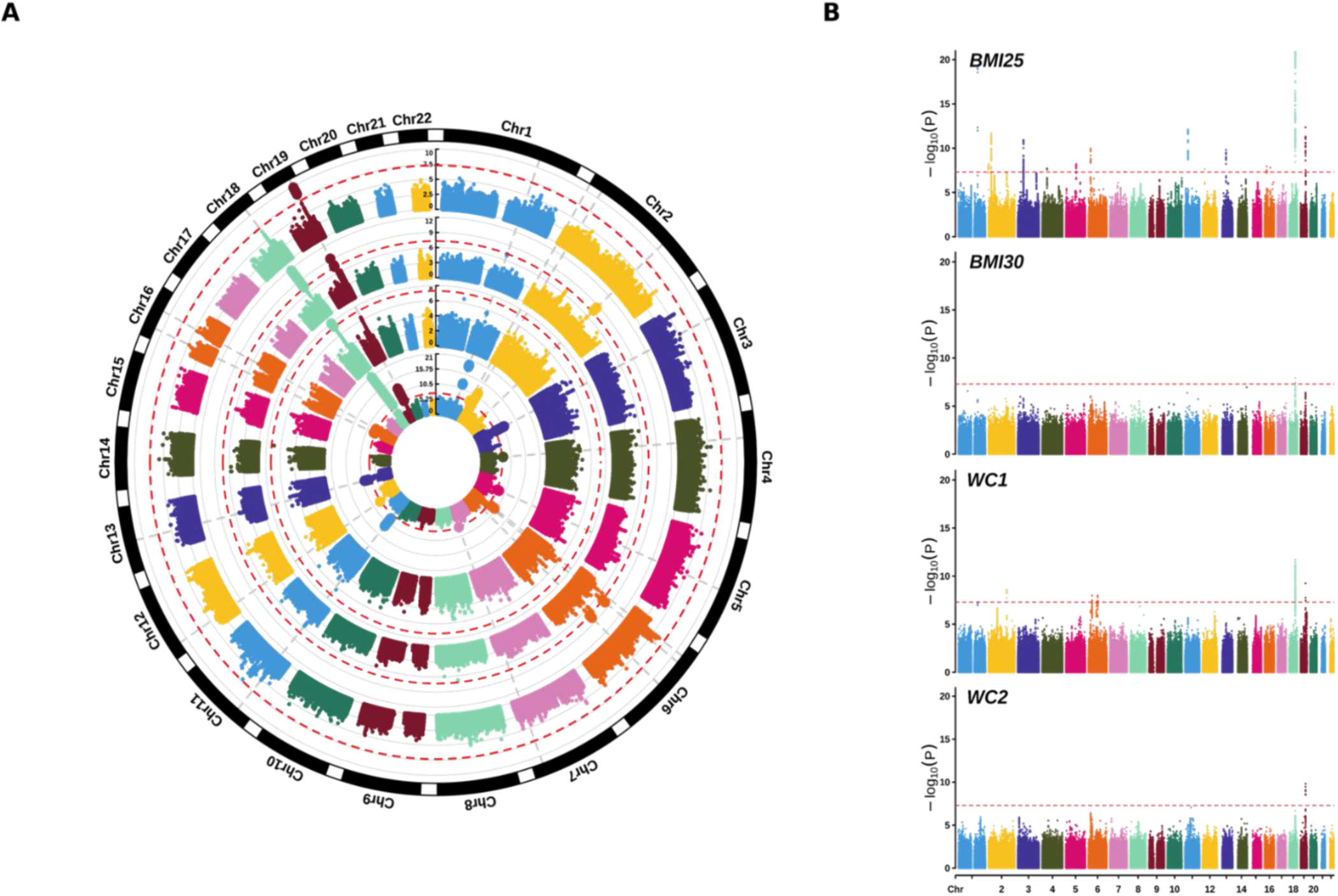
Manhattan plots constructed from GWAS results for obesity in the discovery dataset Circular and standard Manhattan plots are shown for four obesity phenotypes: BMI25 (body mass index ≥ 25 kg/m²), BMI30 (≥ 30 kg/m²), WC1 (waist circumference ≥ 85 cm for females / 90 cm for males), and WC2 (≥ 95 cm for females / 100 cm for males). The red dashed line indicates the genome-wide significance threshold. Panel A presents circular plots arranged from inner to outer rings. Panel B displays the corresponding standard Manhattan plots.

**Table 2.**
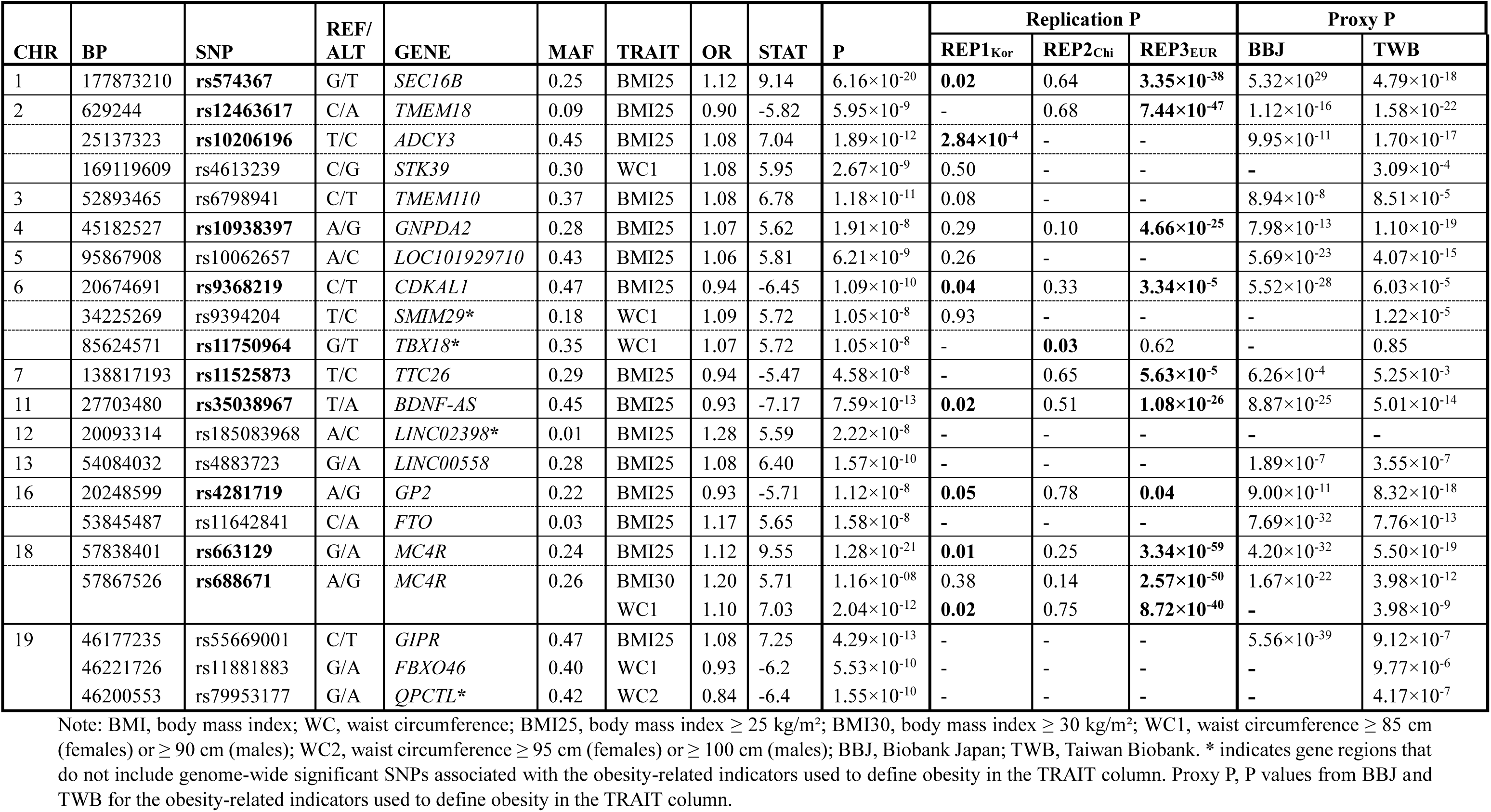
Significant SNPs identified from GWASs of obesity.

For obesity-related indicators, we identified 50 genome-wide significant SNPs (Table S4), 47 of which were replicated in at least one of the external datasets (REP1_Kor_, REP2_Chi_, REP3_NHW_, BBJ or TWB). Four SNPs (rs220063, rs7209484, rs11881883, and rs4817972) appeared to be novel. As visualized in Figure S4, GWASs on continuous outcomes showed broader ranges of significance and more widespread associated regions across chromosomes than dichotomous obesity. Nevertheless, some SNPs, such as rs9394204, were only significant in dichotomous outcome GWASs.

For obesity-related diseases, we identified 59 genome-wide significant SNPs: 27 for HT, 14 for T2D, 1 for CVD, 16 for hyperlipidemia (HL), and 1 for fatty liver (FL), as shown in Figure S5 and Table S5. Among these, 23 SNPs were successfully replicated: 4 for HT, 12 for T2D, 0 for CVD, 6 for HL, and 1 for FL. Two SNPs (rs1268653 and rs6499554) associated with HL, were novel, while two replicated SNPs (rs35612982 in *CDKAL1* and rs66723169 in *MC4R)* overlapped with obesity-associated gene regions. Most SNPs relevant to obesity-related indicators, obesity, and obesity-related diseases in Korean populations also generalized well to Chinese, Japanese, and Taiwanese of EAS populations.

As shown in Table S6A and Table S6B, genomic inflation factors (*λ*_*GC*_) exceeded 1 for all obesity-related indicators, suggesting polygenicity or population structure. However, LD score regression intercepts (*λ*_*LDSC*_) were close to 1, indicating no major confounding due to population stratification, while those from continuous traits in REP3_NHW_ were higher likely due to the large sample size and polygenicity. SNP Heritability estimates were significant for all traits except FL. Notably, moderate obesity definitions showed higher SNP heritability in discovery, REP1_Kor_ and REP3_NHW_ datasets. In contrast, estimates in REP2_Chi_ were negative or insignificant likely due to small sample size.

Regarding genetic correlations, obesity showed strong associations with its target obesity-related indicators. For BMI-based traits, *GC_BMI-BMI25_* = 1.01 (P = 0) and *GC_BMI-BMI30_* = 0.95 (P = 4.22 ×10^-57^), and for WC-based traits, *GC_WC-WC1_* = 0.98 (P = 0) and *GC_WC-WC2_* = 0.90 (P = 1.99 ×10^-45^), indicating overlapping genetic architectures. Similarly, significant correlations were found between obesity and its related diseases in Table S6C. Among Korean individuals, abdominal obesity showed stronger genetic correlations with related diseases, while in NHW populations, general obesity showed stronger correlations.

Local genetic correlation patterns, visualized in Figure S6 and detailed in Table S7, revealed population-specific differences. Transethnic genetic correlations analyses between the EAS meta-analyses and UKB_NHW_ showed strong correlations for moderate levels of general obesity (0.83 with P = 5.02 × 10^−3^) and abdominal obesity (0.76 with P = = 3.41 × 10^−3^), suggesting that many SNPs relevant to obesity in Koreans are also informative in NHW populations.

### Gene-based and gene-set analyses

Gene-based analyses were performed using GWAS results for obesity and related diseases in the discovery dataset. Table S8A and Table S8B present the genes that reached statistical significance level (α = 2.71 × 10^−6^) adjusted by Bonferroni-correction under various definitions of obesity and five obesity-related diseases. While no novel genes were identified for obesity, 13 novel genes were associated with two diseases: 9 for HT (*SPTBN1*, *LRRIQ4*, *STXBP1*, *PTRH1*, *TTC16*, *ARL3*, *SFXN2*, *INA*, and *PDCD11*) and 4 for HL (*APOA4*, *TXNL4B*, *HP*, and *APOE*). No gene reached significance for CVD. Notably, the majority of these genes were also replicated in the UKB_NHW_ dataset, supporting shared genetic mechanisms across populations of different ancestries.

For gene-set analyses, we constructed two sets: one comprising 33 genes related to obesity, and another including 68 genes associated with obesity-related diseases. CVD was excluded due to the lack of suitable GEO target data. As summarized in Table S9, five out of 24 gene-set combinations showed significant associations, primarily from the diseases-related gene-sets. In contrast, the obesity gene-set based on significant genes from the gene-based analyses, did not exhibit significant enrichment in any tested combinations. Further details for HT, HL and FL are provided in Table S10. These results suggest that significant gene-sets are mainly linked to lipid metabolism pathways driven by specific genes. This indicates that, at the pathway level, obesity gene-sets may not directly contribute to the development of associated diseases. These findings highlight the distinction between gene-level and pathway-level associations, suggesting that shared genetic etiology may manifest differently across biological contexts.

### Construction and evaluation of single trait polygenic risk scores for obesity

The prevalence patterns of obesity and obesity-related diseases were consistent between the validation and test datasets, as shown in Table S11. To explore genetic similarities among EAS populations, we estimated SNP heritability and genetic correlations for obesity-related indicators and metabolic traits across Japanese, Taiwanese, and Koreans. As presented in Table S12A, all traits were heritable in EAS populations, with most exhibiting significantly high genetic correlations. Table S12B further showed strong cross-population correlations for key indicators such as BMI and WC (*GC_BBJ-KOR_* = 0.90 with P = 0, *GC_TWB-KOR_* = 0.92 with P = 1.23 × 10^-243^ for BMI, and *GC_TWB-KOR_* = 0.94 with P = 3.59 × 10^-128^ for WC), supporting the suitability of PRS derivation based on these populations for use in Korean dataset.

In the PRS analyses, for obesity, results from the validation phase (Table S13) indicated that models built using LDpred auto algorithm achieved the best performance across all selection metrics. In addition, we constructed PRSs for metabolic health indicators. The best-performing models for each trait were selected based on validation results, as shown in Table S14.

### Construction and performance comparison of single trait and multiple trait polygenic risk score models for obesity and obesity-related diseases

The best-performing PRS models selected during validation were subsequently applied to the test dataset. As shown in Table S15, the single trait PRS model for BMI (M1) outperformed the WC-based PRS model (M2) in predicting general obesity, with significantly higher AUCs for both BMI25 (DeLong P = 1.64 × 10^-18^) and BMI30 (DeLong P = 7.76 × 10^-7^). In contrast, the WC PRS did not exhibit significant improvements in predictive performance for any obesity outcome or obesity-related diseases, except for T2D (DeLong P = 2.40 × 10^-4^).

By comparison, the multiple trait PRS model, which incorporated both obesity metrics (BMI and WC), achieved significantly improved AUCs for all obesity and for HT, compared to the single trait PRS models. Furthermore, an extended multiple trait PRS model (M4), which included additional PRSs for metabolic health indicators, demonstrated improved predictive performance for obesity-related diseases, except for CVD, but did not enhance prediction for obesity outcomes.

### Associations analyses of polygenic risk scores on obesity and obesity-related diseases

The associations between PRSs and both obesity and related diseases are summarized in **Table 3** and **Table 4**. These results showed consistent trends in risk across PRS categories (low (L), medium (M), and high (H)) for both single trait and multiple trait models.

**Table 3.** Effect of single trait PRS on obesity and obesity-related diseases.

| Trait | PRS group | BMI PRS |  |  | WC PRS |  |  |
| --- | --- | --- | --- | --- | --- | --- | --- |
|  |  | Case N (Prev.) | OR (95% CI) | P | Case N (Prev.) | OR (95% CI) | P |
| BMI25 | L | 1494 (21.43%) | 0.54 (0.51, 0.58) | **3.61×10<sup>-87</sup>** | 1726 (24.76%) | 0.65 (0.61, 0.68) | **1.71×10<sup>-49</sup>** |
|  | M (ref.) | 18,508 (33.16%) | - | - | 18,613 (33.35%) | - | - |
|  | H | 3353 (48.06%) | 1.90 (1.80, 2.00) | **4.00×10<sup>-135</sup>** | 3016 (43.18%) | 1.54 (1.46, 1.62) | **1.12×10<sup>-60</sup>** |
| BMI30 | L | 80 (1.15%) | 0.41 (0.32, 0.51) | **6.39×10<sup>-15</sup>** | 103 (1.48%) | 0.50 (0.41, 0.61) | **2.11×10<sup>-11</sup>** |
|  | M (ref.) | 1549 (2.78%) | - | - | 1598 (2.86%) | - | - |
|  | H | 441 (6.32%) | 2.36 (2.11, 2.63) | **7.55×10<sup>-54</sup>** | 369 (5.28%) | 1.91 (1.70, 2.15) | **1.12×10<sup>-27</sup>** |
| WC1 | L | 1188 (18.22%) | 0.65 (0.61, 0.70) | **1.15×10<sup>-36</sup>** | 1192 (18.21%) | 0.64 (0.60, 0.68) | **4.05×10<sup>-39</sup>** |
|  | M (ref.) | 13247 (25.33%) | - | - | 13,250 (25.32%) | - | - |
|  | H | 2279 (34.71%) | 1.59 (1.51, 1.68) | **5.43×10<sup>-61</sup>** | 2272 (34.88%) | 1.61 (1.52, 1.70) | **9.37×10<sup>-63</sup>** |
| WC2 | L | 121 (1.86%) | 0.52 (0.43, 0.62) | **2.88×10<sup>-12</sup>** | 126 (1.92%) | 0.52 (0.43, 0.62) | **1.37×10<sup>-12</sup>** |
|  | M (ref.) | 1844 (3.53%) | - | - | 1868 (3.57%) | - | - |
|  | H | 436 (6.64%) | 1.96 (1.75, 2.18) | **4.90×10<sup>-34</sup>** | 407 (6.25%) | 1.83 (1.64, 2.05) | **1.39×10<sup>-26</sup>** |
| HT | L+M (ref.) | 12,022 (22.07%) | - | - | 12,028 (22.04%) | - | - |
|  | H | 1507 (24.4%) | 1.18 (1.10, 1.25) | **8.94×10<sup>-7</sup>** | 1501 (24.7%) | 1.20 (1.12, 1.28) | **5.03×10<sup>-8</sup>** |
| T2D | L+M (ref.) | 4478 (7.78%) | - | - | 4429 (7.69%) | - | - |
|  | H | 537 (8.36%) | 1.10 (1.00, 1.21) | **0.05** | 586 (9.21%) | 1.23 (1.12, 1.35) | **1.25×10<sup>-5</sup>** |
| CVD | L+M (ref.) | 2504 (4.67%) | - | - | 2490 (4.63%) | - | - |
|  | H | 289 (4.74%) | 1.05 (0.92, 1.19) | 0.47 | 303 (5.07%) | 1.10 (0.97, 1.25) | 0.13 |
| HL | L+M (ref.) | 6166 (11.39%) | - | - | 6136 (11.32%) | - | - |
|  | H | 650 (10.63%) | 0.93 (0.85, 1.01) | 0.11 | 680 (11.29%) | 1.01 (0.93, 1.10) | 0.73 |
| FL | L+M (ref.) | 2553 (5.73%) | - | - | 2563 (5.73%) | - | - |
|  | H | 324 (6.4%) | 1.13 (1.00, 1.28) | **0.04** | 314 (6.39%) | 1.11 (0.98, 1.26) | 0.08 |

Note: BMI, body mass index; WC, waist circumference; BMI25, body mass index $\geq 25$ kg/m<sup>2</sup>; BMI30, body mass index $\geq 30$ kg/m<sup>2</sup>; WC1, waist circumference $\geq 85$ cm (females) or $\geq 90$ cm (males); WC2, waist circumference $\geq 95$ cm (females) or $\geq 100$ cm (males); HT, hypertension; T2D, type 2 diabetes; CVD, cardiovascular disease; HL, hyperlipidemia; FL, fatty liver; L., M. and H., Low, Medium and high PRS group of bottom decile, 2<sup>nd</sup>-9<sup>th</sup> decile, and top decile; OR. and P., odds ratio and significance level of polygenic risk score (PRS) group from logistic model on obesity, adjusted by age, sex, PRS group and the first five principle components.

**Table 4.**
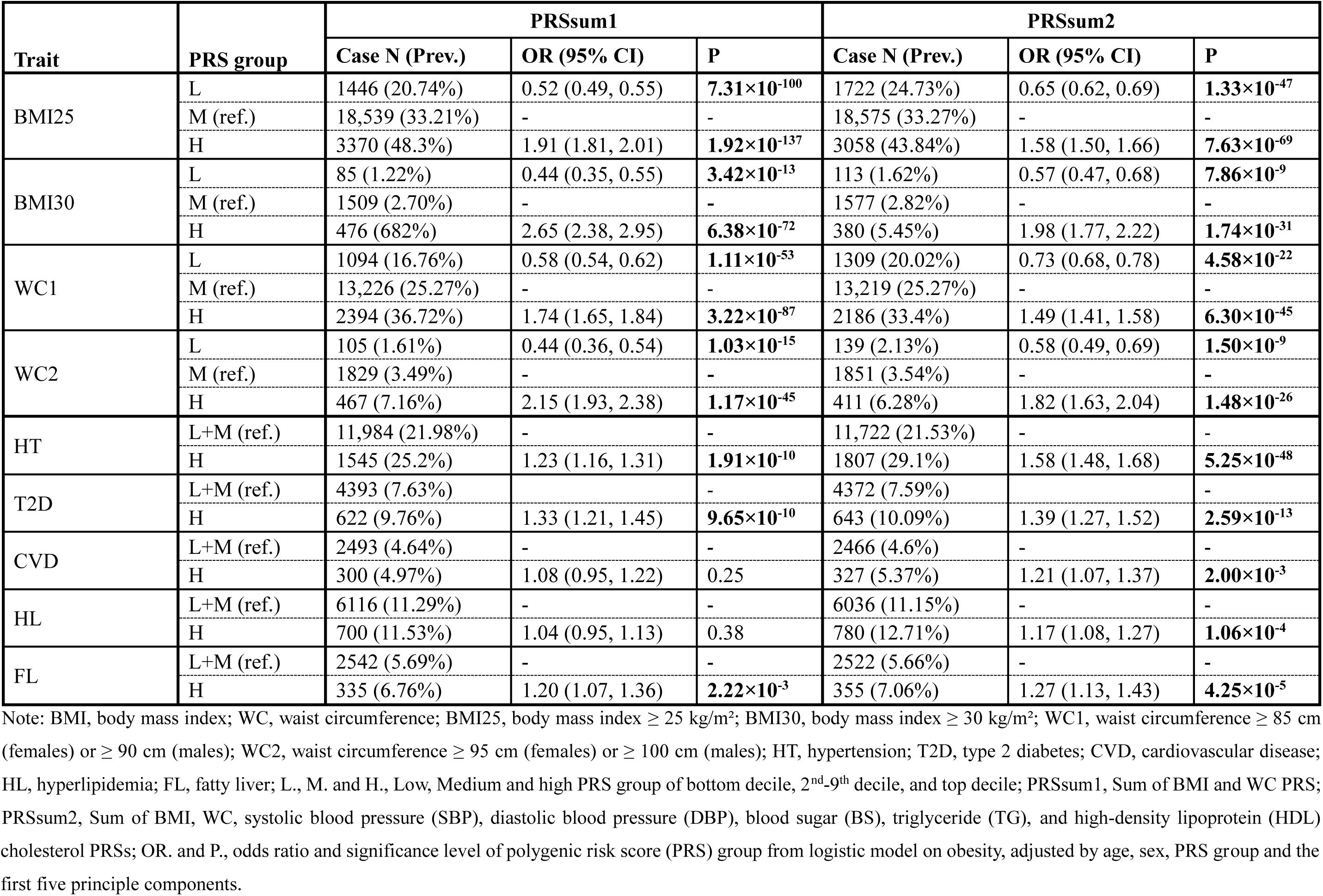
Effect of multiple trait PRSsum on obesity and obesity-related diseases.

Table 3 presents the effects of single trait PRSs for BMI and WC. Across all obesity types, individuals in the L group showed decreased risk, while those in the H group exhibited increased risk. Notably, BMI PRS more clearly differentiated risk for general obesity, while WC PRS showed similar levels of risk in abdominal obesity. Both PRSs also demonstrated significant associations with HT and T2D in the H group. For FL, only the H group of the BMI PRS showed a significant association in odds ratio (OR) (OR = 1.13 with P = 0.04). Interestingly, the WC PRS showed a stronger association with T2D (OR = 1.23 with P = 1.25 × 10^-5^) than the BMI PRS (OR = 1.10 with P = 0.05).

In Table 4, risk associations are stratified by multiple trait PRSs: PRSsum1 for obesity-related indicators only and PRSsum2 for obesity-related indicators with metabolic health indicators. Both models maintained consistent directional trends across obesity outcomes, with reduced ORs in the L group and elevated risks in the H group. PRSsum1 showed stronger associations with obesity-related diseases than single trait PRSs. PRSsum2 further amplified these effects, with significantly increased risks observed for HT (OR = 1.58 with P = 5.25 × 10^-48^), T2D (OR = 1.39 with P = 2.59 × 10^-13^), and FL (OR = 1.27 with P = 4.25 × 10^-5^). Additionally, associations with CVD (OR = 1.21 with P = 2.00 × 10^-3^) and HL (OR = 1.17 with P = 1.06 × 10^-4^), previously non-significant in single trait models, emerged as significant in PRSsum2.

These findings suggest that multiple trait PRSs more effectively capture the complex genetic architectures underlying obesity and its comorbidities. The comparative effects of both PRS types are visualized in **Figure 3**. These results reinforce the utility of composite genetic models for risk stratification in clinical settings, particularly within EAS populations.

**Figure 3.**
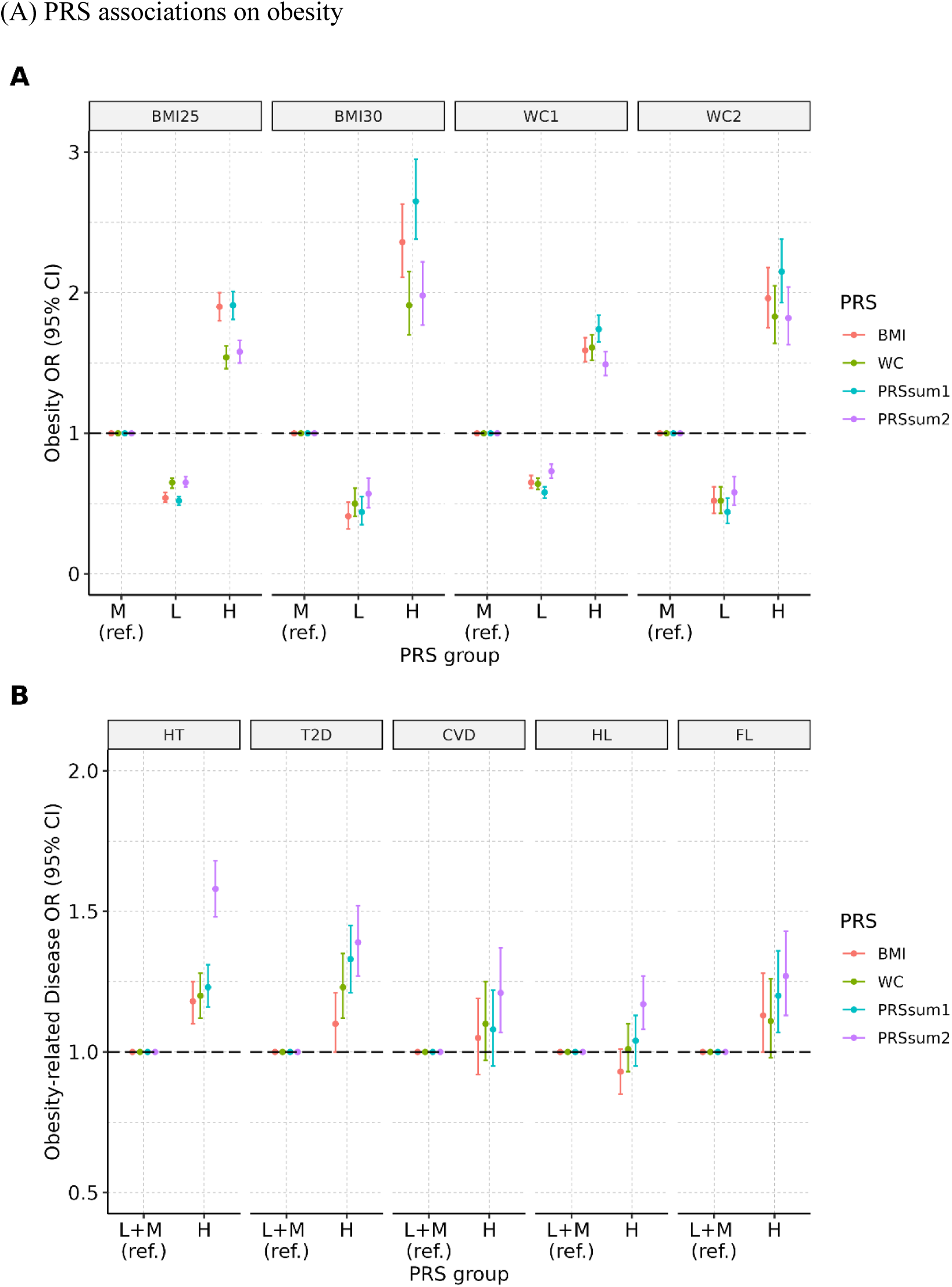
PRS associations on obesity and obesity related diseases (A) PRS associations on obesity Associations of PRS with obesity and related diseases are illustrated. PRSsum1 represents the sum of standardized PRSs for BMI and WC; PRSsum2 includes PRSs for BMI, WC, and metabolic health traits (systolic blood pressure, diastolic blood pressure, fasting plasma glucose, triglycerides, and high-density lipoprotein cholesterol).

### Interaction and mediation analyses of polygenic risk scores for obesity on obesity-related diseases

To evaluate interaction effects between obesity and related diseases, we calculated and validated PRSs for T2D and CVD, as shown in Table S16. We then assessed the interactions between obesity (defined either by phenotype or PRS) and the corresponding disease PRSs. As shown in Table S17A, both the target disease PRS and obesity classification were highly significant predictors in all regression models. In contrast, the obesity PRSs showed relatively weaker significance, with some effects being non-significant. Interestingly, we observed significant interaction effects between obesity and T2D, indicating antagonistic interactions that while both obesity and T2D PRS independently increased disease risk, their interaction term was associated with a reduction in that risk.

We further performed mediation analyses to disentangle the direct and indirect effects of obesity PRSs on T2D and CVD. Overall, the mediation effects were significant. Notably, for the BMI PRS, the direct effect was non-significant, while the indirect effect via obesity was significant. This pattern suggests a strong mediation pathway, indicating that BMI PRS likely exerts its influence on T2D and CVD primarily through general obesity. In contrast, other PRSs, except for the BMI PRS on T2D/CVD and PRSsum1 on CVD, exhibited partial mediation, with both significant direct and indirect effects. Detailed mediation results are summarized in in Table S17B. These findings highlight the role of obesity as a key intermediate phenotype linking genetic risk factors to cardiometabolic outcomes, particularly in EAS populations.

## Discussion

In this study, we investigated the genetic factors underlying obesity and their influence on obesity-related diseases in a large Korean cohort, representing EAS populations. We hypothesized that genetic predisposition to obesity is shared between general obesity and abdominal obesity, as well as across EAS and NHW populations, despite differences in genetic backgrounds. Furthermore, we compared the performance of traditional single trait PRSs with multiple trait approaches, including PRSsum method, to evaluate their predictive accuracy and potential for identifying shared genetic components. Using PRS-derived risk profiles, we further explored the mechanisms by which genetic susceptibility to obesity contributes to disease onset through interaction and mediation analyses.

We commenced out investigation by conducting GWASs on two obesity-related indicators and two definition of obesity, each with two severity levels, alongside five obesity-related diseases. Our analyses revealed 50 genome-wide significant SNPs for obesity-related indicators, of which 47 were replicated across populations. These included 34 SNPs for BMI and 13 for WC, with six SNPs overlapping, as shown in Table S4. Among these, four SNPs, rs220063 (*ZEB1*), rs7209484 (*NFE2L1*), rs11881883 (*FBXO46*), and rs4817972 (*LOC400867*), were novel. Although these genes have not directly associated with obesity in previous studies, prior literatures suggest indirect relevance. *ZEB1* transcription factor and *NFE2LI* isoforms have been linked to the regulation of adiposity in mouse models [34, 35], *FBXO46* gene has been associated with BMI [36], and CpG representing the *LOC400867* gene with birth weight [37].

In the GWAS of obesity, we identified 22 genome-wide significant SNPs (Table 2), among which 11 were replicated: 10 for general obesity and two for abdominal obesity, with one overlapping. When comparing GWAS results on continuous and dichotomized outcomes, we observed substantial overlap, with genetic correlations exceeding 0.90 and visualization in Figure S4. Notably, continuous traits exhibited a broader range of associations, while dichotomous outcomes retained unique findings, highlighting the importance of considering multiple definition of target.

In our analysis of obesity-related diseases, we identified 59 genome-wide significant SNPs (Table S5), including two novel loci (rs1268353 in *BUD13* and rs6499554 in *PKD1L3*) for HL. The overlap between obesity-associated and disease-associated SNPs was limited, with only two SNPs located in shared gene regions (*CDKAL1* and *MC4R*). Nevertheless, EAS - NHW transethnic genetic correlations were significant: 0.83 for general obesity and 0.76 for abdominal obesity, demonstrating transferability of genetic signals across EAS and NHW populations (Table S6C). Furthermore, EAS populations exhibited higher correlations between abdominal obesity and obesity-related diseases, compared to NHW counterparts. Distinct patterns of local genetic correlations were observed in Figure S6 and Table S7.

Gene-based analyses identified significant genes for both obesity and its related diseases (Table S8), including *CDKAL1* as a shared gene between general obesity and T2D. However, gene-set analyses (Table S9 and S10) did not reveal significant enrichment, suggesting limited overlap at the pathway level. These findings emphasize the need for ethnic-specific approaches to decipher the genetic architecture of obesity and related disorders.

Using PRSs derived from both BMI and WC, we confirmed that individuals in high risk groups showed increased susceptibility to obesity, HT, T2D, and FL (Table 3). WC PRS particularly associated with T2D, while BMI PRS showed stronger associations with FL. Multiple trait PRSs demonstrated improved predictive accuracy (Table S15), particularly when combining PRSs for obesity-related indicators and metabolic health traits (PRSsum2), which further enhanced prediction of disease outcomes such as HT, T2D, and CVD (Table 4). These findings suggest that multiple trait PRSs better capture the polygenic architecture of comorbidities than single trait PRSs, especially when the PRS traits and outcome are aligned.

To further understand the biological mechanisms linking genetic predisposition of obesity to disease, we performed interaction and mediation analyses on obesity-related diseases such as T2D and CVD (Table S17A and Table S17B). Although interactions between obesity PRSs and disease PRSs were not significant, we observed significant interactions between obesity phenotypes and disease PRSs. Mediation analyses revealed that the total effects of obesity PRSs on T2D and CVD were significant, primarily driven by indirect effects through obesity. In particular, the direct effects of BMI PRS on disease were non-significant, suggesting full mediation through general obesity. Other PRSs, such as WC PRS, PRSsum1 and PRSsum2, showed partial mediation effects.

While our findings provide novel insights, several limitations should be acknowledged. First, the study population was limited to Koreans. Although genetic similarity among EAS populations was supported by high cross population correlations (Table S12B), subtle population specific differences may affect generalizability. Second, while multiple trait PRSs improved performance for many outcomes, the optimal PRS composition of each phenotype requires further evaluation. Despite these limitations, our study, the largest genetic study of obesity in a Korean population to date, offers provides valuable insights into the shared genetic architectures of obesity and its related diseases. These findings support the development of population-specific polygenic models to inform precision medicine initiatives.

## Materials and Methods

### Research participants and genotype data processing

A comprehensive overview of the study population and genotype data has been described previously [33]; here, provide a brief summary tailored to the present analysis. Geneotype data were obtained from a total of 231,302 participants enrolled in four Korean cohorts, all genotyped using Korean Biobank Array (Koreanchip) [38]: These cohorts include the Korean Genome Epidemiology Study (KoGES) [39], Gene-Environment of Interaction and Phenotype (GENIE) [40], YonSei University Hospital medical center (YSUH), and the Veterans Health Service Medical Center (VHSMC). For replication purposes, we incorporated three independent datasets: 1) **REP1_Kor_**, an additional Korean subset from KoGES genotyped with Affymetrix Arrays (N = 8856); 2) **REP2_Chi_**, comprising individuals of Chinese ancestry dataset (N = 1503) from UK Biobank (UKB) [41]; and 3) **REP3_NHW_**, a large-scale European ancestry dataset (N = 459,259) also from UKB. Genotype data underwent standardized quality-control (QC) procedures, including filtering based on individual and SNP call rate, Hardy-Weinberg equilibrium, and minor allele frequency (MAF). Phasing was performed using the Eagle v2.4 [42] and imputation was carried out via the Northeast Asian Reference Database imputation server [43]. Additional genotype processing and QC were conducted using PLINK [44, 45] and GCTA [46].

After applying harmonized inclusion and exclusion criteria, the resulting datasets includes a discovery dataset of 85,947 Korean individuals with 4,736,957 SNPs, and three replication datasets of consisting of 7726 Koreans with 2,221,602 SNPs, 1496 Chinese individuals with 2,076,717 SNPs, and 392,160 NHW individuals with 4,936,389 SNPs.

### Anthropometric indicators of obesity and obesity-related diseases

Obesity-related phenotypes were evaluated based on BMI and WC, following criteria established for Korean population [12, 14]. For general obesity, individuals of EAS ancestry were classified as obese (case) if their BMI ≥ 25 kg/m^2^, whereas NHW individuals were considered obese at BMI ≥ 30 kg/m^2^. Individuals below these thresholds were categorized as controls (referred to BMI25 and BMI30 criteria, respectively). Severe obesity was further stratified as BMI ≥ 30 kg/m^2^ for EAS populations (BMI30) and BMI ≥ 40 kg/m^2^ for NHW populations (BMI40). Additionally, abdominal obesity was defined uniformly across all ancestry groups as WC ≥ 85 cm in females and ≥ 90 cm in males (WC1) for moderate level, and WC ≥ 95 cm for females or ≥ 100 cm for males (WC2) for severe level, due to the absence of ancestry-specific guidelines for this trait. While BMI-based definitions were adjusted by ancestry, WC-based definitions remained consistent across populations.

For obesity-related diseases, we assessed five conditions at baseline: HT, T2D, CVD, HL and FL. CVD encompassed both coronary heart disease and cerebrovascular accident. Disease status was determined primarily by self-reported diagnosis. In case of T2D, diagnosis was based either on self-report or on meeting one or more of the following clinical criteria: 1) FPG levels ≥ 126 mg/dL, 2) 2-hour postprandial plasma glucose ≥ 200 mg/dL, or 3) glycated hemoglobin of HbA1c ≥ 6.5 %.

### Genome-wide association studies, meta-analyses, and correlation analyses

GWAS were conducted using a modified version of our previously described analytical framework [33], tailored to the design of the present study. We aimed to identify genetic variants associated with obesity, its related indicators, and comorbid conditions using the discovery dataset. Specifically, for obesity-related indicators, we applied a rank-base inverse normal transformation. Linear and logistic regression models were applied through PLINK, with age, sex, and the top ten principal components (PCs) included as covariates. A genome-wide significance threshold was set at α = 5×10^−8^. To reduce redundancy, SNPs in high linkage disequilibrium (*r^2^* > 0.1) with top signals were excluded. The remaining lead SNPs were annotated using ANNOVAR [47]. For replication of GWAS findings related to obesity and obesity-related indicators, all three replication datasets were utilized. To evaluate generalizability of Korean-derived associations to other EAS populations, we compared results with publicly available GWAS summary statistics for BMI from BBJ [48] (http://jenger.riken.jp/en) and WC from TWB [49] (https://www.ebi.ac.uk/gwas/publications/38116116). For obesity-related diseases, SNP validation was performed primarily in the REP3_NHW_ dataset and partially in BBJ limited to T2D and CVD, focusing on large-scale data sources.

Summary level GWAS results were analyzed using LDSC [50] to assess genomic inflation and to estimate SNP-based heritability. Genetic correlations among obesity-related indicators and between these traits and obesity were quantified in the discovery dataset using LDSC. In addition, correlations between obesity and obesity-related diseases were evaluated in both the discovery and REP3_NHW_ datasets. While LDSC was used to estimate genome-wide correlations, local genetic correlations were estimated using LAVA [51]. To explore shared genetic architecture between EAS and NHW populations, we conducted meta-analyses using GWAMA [52], integrating summary statistics from the discovery and two replication datasets (REP1_Kor_ and REP2_Chi_). Transethnic genetic correlations were further examined for moderate obesity (BMI25/WC1 for EAS and BMI30/WC1 for NHW) using POPCORN [53].

### Gene-based and gene-set analyses

We performed gene-based association analyses across 18,432 genes using the discovery dataset, REP1_Kor_, and REP2_Chi_. These analyses were conducted with MAGMA [54], applying the 1000G EAS reference panel. For analyses in the UKB_NHW_ population, the 1000G EUR reference data were used instead to account for ancestry-specific genetic structures.

To further examine the biological relevance of obesity- and disease-associated genes, we conducted gene-set analyses focusing on HT, T2D, and HL. This process involved three key steps. First, we defined two separate gene sets derived from significant gene-based results: one for obesity (BMI25/WC1) and one for obesity-related diseases (HT, T2D, HL). In the second step, we extracted gene expression data from three GEO datasets (GSE24752 for HT, GSE278204 for T2D, and GSE1010 for HL) and calculated log fold changes using the DESeq2 package (version 1.46.0) [55]. Finally, we performed two types of gene-set analyses using R: 1) gene set enrichment analysis (GSEA) via the fgsea package (version 1.32.2) [56] based on the Reactome database, and 2) over-representation analysis (ORA) with the clusterProfiler package (v4.14.6) [57], referencing both GO and KEGG databases.

### Construction and evaluation of single trait polygenic risk scores for obesity

To evaluate genetic susceptibility to obesity, we constructed PRSs using genome-wide summary statistics from BBJ and the TWB. Variants with MAF greater than 0.005 were retained, resulting in 5,925,388 SNPs from BBJ and 7,614,251 SNPs from TWB for downstream analysis. These sets were used as the source for PRS construction. Multiple widely used PRS methods were applied, including clumping and thresholding (CT) [58], LDpred (infinitesimal, grid and auto models) [59, 60], lassosum [61] and PRS-CS [62]. Linkage disequilibrium was estimated using the EAS reference panel from the 1000G project. For LDpred grid models, various prior probabilities for the proportion of causal variants were tested (*ρ* = 1%, 3%, 10%, 30% and 100%).

For model evaluation, we split the discovery dataset by randomly assigning 15,000 individuals for validation and the remaining 70,947 for testing. During the validation phase, 10-fold cross-validation was conducted using linear or logistic regression models, incorporating age, sex, the corresponding PRS, and the first five PCs as covariates. The optimal PRS from each method was selected based on multiple performance criteria: 1) the strength of correlation with obesity-related traits, 2) statistical significance, 3) model fit assessed via the Akaike Information Criterion (AIC), and 4) predictive accuracy, as indicated by the area under the curve (AUC) for binary outcomes.

### Construction and evaluation of multiple trait polygenic risk score models for obesity and obesity-related diseases

To enhance the genetic prediction of obesity and its comorbid diseases, we implemented a multiple trait PRS strategy [63] that incorporates information across several obesity-related traits. Specifically, we constructed PRSs for two primary obesity-related indicators (BMI and WC), as well as five metabolic traits (SBP, DBP, BS for FPG, TG, and HDL). These trait-specific PRSs were generated using single trait PRS modeling methods, with summary statistics primarily derived from BBJ, except for WC, which utilized data from the TWB. To assess the transferability of these PRSs from the Japanese and Taiwanese training populations to the Korean target population, we estimated SNP-based heritability and pairwise genetic correlations within and across EAS populations using LDSC. PRS validation was performed using the same criteria as in the single trait models, excluding AUC during initial selection.

For comparative evaluation, four PRS models were examined: two single trait PRSs of BMI (Model 1, M1) and WC (Model 2, M2), and two multiple trait models of obesity-related indicators only (Model 3, M3) and obesity-related indicators combined with metabolic traits (Model 4, M4). Predictive performance was assessed using the AIC and AUC. Model comparisons were conducted using the DeLong test [64].

### Association analyses of polygenic risk scores on obesity and obesity-related diseases

We evaluated the associations between PRSs and both obesity and obesity-related diseases using logistic regression models. Four PRSs, derived from two distinct modeling strategies of single trait and multiple trait approaches, were analyzed. Additionally, we implemented the PRSsum method [65], in which multiple standardized PRSs are aggregated with equal weights to create composite scores. Two types of PRSsum were constructed: PRSsum1, composed solely of obesity-related indicators, and PRSsum2, which includes both obesity indicators and metabolic health traits. This approach was intended to assess whether these composite indices more effectively identify individuals at elevated risk for obesity and related conditions.

For each PRS model, individuals were stratified into three risk categories comprising of L (bottom 10%), M (middle 80%), and H (top 10%), corresponding to the low, medium, and high risk in PRS distribution. In disease risk associations, we focused on comparing the H group, presumed to carry elevated genetic risk, against the combined L and M groups to examine its discriminatory capacity.

### Interaction and mediation analyses of genetics of obesity on obesity-related diseases

To investigate how genetic predisposition to obesity influences its related comorbidities, we conducted both interaction and mediation analyses. These analyses focused on T2D and CVD, for which publicly available summary statistics from BBJ were accessible. Validation procedures for PRS construction and performance followed the same protocols described in earlier sections.

For the interaction analysis, logistic regression models were applied with the following covariates: age, sex, the top five PCs, obesity status, obesity PRSs (including single trait PRSs for BMI and WC, and multiple trait PRSsum for PRSsum1 and PRSsum2), the target disease PRS, and an interaction term, either obesity × disease PRS or obesity PRS × disease PRS. Mediation analyses [66] were carried out to quantify the indirect effects of obesity between genetic risk and disease outcomes. The model framework specified obesity PRS as the independent variable (X), obesity status as the mediator (M), and obesity-related disease as the dependent variable (Y). This framework paralleled the four PRSs used in interaction analyses. Mediation was tested in three sequential models: 1) disease outcome ∼ age + sex + obesity PRS, 2) obesity ∼ age + sex + obesity PRS, and 3) disease outcome ∼ age + sex + obesity PRS + obesity. All mediation analyses were conducted using the mediation package (version 4.5.0) [67] in R.

## Ethical statement

No animal experiments were involved in this study.

This research was designed in accordance with the principles of Declaration of Helsinki, and conducted after approval of the institutional review board of Seoul National University (IRB No. E2303/004-012).

**KoGES:** This study was conducted with bioresources from National Biobank of Korea, the Korea Disease Control and Prevention Agency, Republic of Korea (IRB No. E2308/001-020) and was approved under Accession ID: KBN-2020-101 (https://nih.go.kr/contents.es?mid=a50401010100).

**GENIE:** As a sub-cohort of H-PEACE study, this study was conducted with bioresources from the Seoul National University Hospital Gangnam Center with institutional review board (IRB No. H-2102-068-1196) and data access was granted through http://en-healthcare.snuh.org/HPEACEstudy.

**VHSMC:** This study was conducted with bioresources from the VHS Biobank with institutional review board (IRB No. BOHUN-2020-12-002) and was approved under Accession ID: VBP-2020-03.

**UK Biobank:** This research has been conducted using the UK Biobank Resource under Project Number 41056 and data access was granted through at https://www.ukbiobank.ac.uk.

**Biobank Japan** and **Taiwan biobank**: These summary statistics were obtained from publicly available sources, and no additional ethical approval was required.

## Data availability

The GWAS summary statistics generated in this study from the discovery dataset—including BMI, BMI25, BMI30, WC, WC1, and WC2—will be publicly available in the GWAS Catalog under the following accession numbers before the publication.

## CRediT author statement

**Jinyeon Jo**: Conceptualization, Data curation, Formal analysis, Investigation, Methodology, Validation, Visualization, Writing - original draft, and Writing - review & editing. **Nayoung Ha**: Writing – review & editing. **Yunmi Ji**: Writing – review & editing. **Ahra Do**: Data curation. **Je Hyun Seo**: Resources and Writing – review & editing. **Bumjo Oh**: Resources and Writing – review & editing. **Sungkyoung Choi**: Writing – review & editing. **Eun Kyung Choe**: Resources and Writing – review & editing. **Woojoo Lee**: Supervision, Project administration, and Writing – review & editing. **Jang Won Son**: Conceptualization, Funding acquisition, Investigation, Project administration, Supervision, and Writing – review & editing. **Sungho Won**: Conceptualization, Funding acquisition, Methodology, Project administration, Resources, Supervision, and Writing – review & editing.

## Acknowledgements

This study was conducted with bioresources from National Biobank of Korea, the Korea Disease Control and Prevention Agency, Republic of Korea (KBN-2020-101). This work was supported by the National Research Foundation of Korea (NRF) grant funded by the Korean government (MSIT) (No. RS-2021-NR060088 and No. RS-2024-00346850). This research was supported by a grant of ‘Korea Health Technology R&D Project’ through the Korea Health Industry Development Institute (KHIDI), funded by the Ministry of Health & Welfare, Republic of Korea (grant number: RS-2024-00403700). Statistical analyses were supported by the national supercomputing center with supercomputing resources including technical support (KSC-2022-CRE-0319 and KSC-2023-CRE-0117). This study was supported by Research Grant from Korean Society for the Study of Obesity (Grant No. KSSO-D-2021002).

