## Supplementary Tables and Figures for "Association of Multiple Trait Polygenic Risk Score with Obesity and Cardiometabolic Diseases in Korean population"

**Table S1.** Baseline characteristics according to the incident obesity on $REP1_{Kor}$

(A) Baseline characteristics according to the incident general obesity

| Variables | Total (N=7089) | BMI25 (Prevalence = 39.70%) Non-obese (N=4275) | BMI25 Obese (N=2814) | BMI25 P | BMI30 (Prevalence = 4.57%) Non-obese (N=6765) | BMI30 Obese (N=324) | BMI30 P | Missing data, N (%) |
|---|---|---|---|---|---|---|---|---|
| **Age (years)** | 54.55 ± 9.06 | 54.30 ± 9.21 | 54.92 ± 8.82 | 0.004 | 54.52 ± 9.09 | 55.21 ± 8.49 | 0.151 | 0 (0%) |
| **Sex**, n (%) | | | | <0.001 | | | <0.001 | 0 (0%) |
| Female | 3850 (54.31%) | 2279 (59.19%) | 1571 (40.81%) | | 3615 (93.90%) | 235 (6.10%) | | |
| Male | 3239 (45.69%) | 1996 (61.62%) | 1243 (38.38%) | | 3150 (97.25%) | 89 (2.75%) | | |
| **Body mass index** ($kg/m^2$) | 24.37 ± 3.13 | 22.39 ± 1.82 | 27.38 ± 2.14 | <0.001 | 24.01 ± 2.68 | 31.94 ± 1.91 | <0.001 | 0 (0%) |
| **Waist circumference** (cm) | | | | <0.001 | | | <0.001 | 29 (0.41%) |
| Female | 81.66 ± 9.34 | 76.79 ± 7.03 | 88.15 ± 7.91 | | 80.48 ± 8.46 | 96.12 ± 9.04 | | |
| Male | 85.54 ± 7.84 | 81.61 ± 6.29 | 91.86 ± 5.67 | | 85.11 ± 7.47 | 100.98 ± 4.86 | | |
| **Body fat ratio** (%) | 27.44 ± 7.30 | 24.23 ± 6.25 | 31.66 ± 6.36 | <0.001 | 26.82 ± 6.90 | 38.14 ± 5.54 | <0.001 | 4433 (62.53%) |
| **Abdominal fat ratio** (%) | 0.91 ± 0.05 | 0.88 ± 0.04 | 0.94 ± 0.04 | <0.001 | 0.90 ± 0.04 | 1.00 ± 0.05 | <0.001 | 4433 (62.53%) |
| **Systolic blood pressure** (mm Hg) | 124.64 ± 17.90 | 122.46 ± 17.74 | 127.96 ± 17.64 | <0.001 | 124.33 ± 17.78 | 131.18 ± 19.12 | <0.001 | 2 (0.03%) |
| **Diastolic blood pressure** (mm Hg) | 79.60 ± 11.25 | 78.03 ± 11.15 | 81.99 ± 10.99 | <0.001 | 79.37 ± 11.22 | 84.46 ± 10.90 | <0.001 | 2 (0.03%) |
| **Fasting plasma glucose** (mg/dL) | 97.21 ± 33.33 | 94.78 ± 29.98 | 100.91 ± 37.57 | <0.001 | 96.90 ± 33.29 | 103.56 ± 33.49 | 0.001 | 68 (0.96%) |
| **HbA1c** (%) | 5.86 ± 0.99 | 5.76 ± 0.95 | 5.98 ± 1.03 | <0.001 | 5.84 ± 0.99 | 6.16 ± 1.03 | <0.001 | 4434 (62.55%) |
| **Triglyceride** (mg/dL) | 151.38 ± 110.86 | 137.04 ± 104.65 | 173.16 ± 116.36 | <0.001 | 149.68 ± 109.35 | 186.93 ± 134.01 | <0.001 | 4 (0.06%) |
| **High density lipoprotein** (mg/dL) | | | | <0.001 | | | <0.001 | 0 (0%) |
| Female | 49.93 ± 13.01 | 52.61 ± 13.53 | 47.00 ± 11.79 | | 50.66 ± 13.17 | 45.13 ± 11.42 | | |
| Male | 46.88 ± 12.22 | 48.73 ± 12.72 | 43.90 ± 10.74 | | 47.02 ± 12.22 | 41.78 ± 10.99 | | |
| **Hypertension**, case n (%) | 1488 (21.00%) | 660 (15.44%) | 828 (29.43%) | <0.001 | 1359 (20.09%) | 129 (39.81%) | <0.001 | 2 (0.03%) |
| **Type 2 Diabetes**, case n (%) | 973 (14.59%) | 472 (11.63%) | 501 (19.18%) | <0.001 | 901 (14.15%) | 72 (23.76%) | <0.001 | 420 (5.92%) |
| **Cardiovascular Diseases**, case n (%) | 249 (3.51%) | 134 (3.14%) | 115 (4.09%) | <0.001 | 230 (3.40%) | 19 (5.86%) | <0.001 | 2 (0.03%) |
| **Hyperlipidemia**, case n (%) | 318 (4.49%) | 157 (3.67%) | 161 (5.72%) | <0.001 | 296 (4.38%) | 22 (6.79%) | <0.001 | 2 (0.03%) |
| **Fatty liver**, case n (%) | 290 (6.54%) | 114 (4.12%) | 176 (10.56%) | <0.001 | 264 (6.21%) | 26 (14.44%) | <0.001 | 2657 (37.48%) |

(B) Baseline characteristics according to the incident abdominal obesity

| Variables | Total (N=7697) | WC1 (Prevalence = 33.19%) | | | WC2 (Prevalence = 6.51%) | | | Missing data, N (%) |
|---|---|---|---|---|---|---|---|---|
| | | Non-obese (N=5142) | Obese (N=2555) | P | Non-obese (N=7196) | Obese (N=501) | P | |
| **Age (years)** | 54.42 ± 9.10 | 53.42 ± 9.10 | 57.12 ± 8.50 | <0.001 | 54.41 ± 9.06 | 58.17 ± 8.48 | <0.001 | 0 (0%) |
| **Sex**, n (%) | | | | <0.001 | | | <0.001 | 0 (0%) |
| Female | 4145 (63.55%) | 2634 (63.55%) | 1511 (36.45%) | | 3767 (90.88%) | 378 (9.12%) | | |
| Male | 3552 (46.15%) | 2508 (70.61%) | 1044 (29.39%) | | 3429 (96.54%) | 123 (3.46%) | | |
| **Body mass index** ($kg/m^2$) | 24.37 ± 3.12 | 23.05 ± 2.37 | 27.04 ± 2.73 | <0.001 | 24.01 ± 2.79 | 29.65 ± 2.98 | <0.001 | 637 (8.28%) |
| **Waist circumference** (cm) | | | | <0.001 | | | <0.001 | 0 (0%) |
| Female | 81.66 ± 9.34 | 76.03 ± 5.70 | 91.48 ± 5.58 | | 79.89 ± 7.72 | 99.34 ± 4.32 | | |
| Male | 85.39 ± 7.88 | 81.57 ± 5.59 | 94.58 ± 4.06 | | 84.77 ± 7.25 | 102.90 ± 3.06 | | |
| **Body fat ratio** (%) | 27.44 ± 7.30 | 25.25 ± 6.68 | 32.51 ± 6.04 | <0.001 | 26.78 ± 6.99 | 36.29 ± 5.36 | <0.001 | 5043 (65.52%) |
| **Abdominal fat ratio** (%) | 0.91 ± 0.05 | 0.89 ± 0.04 | 0.95 ± 0.05 | <0.001 | 0.90 ± 0.05 | 0.97 ± 0.05 | <0.001 | 5043 (65.52%) |
| **Systolic blood pressure** (mm Hg) | 124.85 ± 18.14 | 122.43 ± 17.83 | 129.73 ± 17.77 | <0.001 | 124.35 ± 18.03 | 132.04 ± 18.08 | <0.001 | 2 (0.03%) |
| **Diastolic blood pressure** (mm Hg) | 79.82 ± 11.30 | 78.42 ± 11.22 | 82.66 ± 10.91 | <0.001 | 79.53 ± 11.28 | 84.02 ± 10.64 | <0.001 | 2 (0.03%) |
| **Fasting plasma glucose** (mg/dL) | 96.39 ± 32.76 | 93.77 ± 31.62 | 101.74 ± 34.36 | <0.001 | 95.88 ± 32.19 | 103.91 ± 39.54 | <0.001 | 126 (1.64%) |
| **HbA1c** (%) | 5.87 ± 1.01 | 5.76 ±0.94 | 6.11 ± 1.11 | <0.001 | 5.85 ± 1.00 | 6.17 ± 1.08 | <0.001 | 4411 (57.31%) |
| **Triglyceride** (mg/dL) | 152.82 ± 110.57 | 139.59 ± 103.92 | 179.44 ± 118.49 | <0.001 | 150.51 ± 110.50 | 186.06 ± 106.33 | <0.001 | 6 (0.08%) |
| **High density lipoprotein** (mg/dL) | | | | <0.001 | | | <0.001 | 1 (0.01%) |
| Female | 49.93 ± 13.01 | 52.09 ± 13.51 | 46.26 ± 11.20 | | 50.53 ± 13.15 | 44.35 ± 10.13 | | |
| Male | 46.72 ± 12.18 | 48.11 ± 12.48 | 43.36 ± 10.75 | | 46.88 ± 12.21 | 41.89 ± 10.60 | | |
| **Hypertension**, case n (%) | 1613 (20.96%) | 785 (15.27%) | 828 (32.41%) | <0.001 | 1613 (20.96%) | 1402 (19.49%) | <0.001 | 1 (0.01%) |
| **Type 2 Diabetes**, case n (%) | 1067 (14.67%) | 536 (10.89%) | 531 (22.62%) | <0.001 | 1067 (14.67%) | 955 (14.00%) | <0.001 | 426 (5.53%) |
| **Cardiovascular Diseases**, case n (%) | 288 (3.74%) | 157 (3.05%) | 131 (5.13%) | <0.001 | 288 (3.74%) | 253 (3.52%) | <0.001 | 1 (0.01%) |
| **Hyperlipidemia**, case n (%) | 324 (4.21%) | 197 (3.83%) | 127 (4.97%) | <0.001 | 324 (4.21%) | 294 (4.09%) | <0.001 | 1 (0.01%) |
| **Fatty liver**, case n (%) | 289 (6.55%) | 140 (4.86%) | 149 (9.74%) | <0.001 | 289 (6.55%) | 260 (6.27%) | <0.001 | 3288 (42.72%) |

Note: BMI, body mass index; WC, waist circumference; BMI25, BMI ≥ 25 $kg/m^2$; BMI30, BMI ≥ 30 $kg/m^2$; BMI40, BMI ≥ 40 $kg/m^2$; WC1, WC ≥ 85 cm for females / 90 cm for males; WC2, WC ≥ 95 cm for females / 100 cm for males.

**Table S2.** Baseline characteristics according to the incident obesity on REP2$_{Chi}$

(A) Baseline characteristics according to the incident general obesity

| Variables | Total (N=1494) | BMI25 (Prevalence = 35.88%) Non-obese (N=958) | Obese (N=536) | P | BMI30 (Prevalence = 5.35%) Non-obese (N=1414) | Obese (N=80) | P | Missing data, N (%) |
|---|---|---|---|---|---|---|---|---|
| **Age (years)** | 52.44 ± 7.65 | 52.09 ± 7.58 | 53.05 ± 7.74 | 0.020 | 52.48 ± 7.66 | 51.73 ± 7.59 | 0.393 | 0 (0%) |
| **Sex**, n (%) | | | | <0.001 | | | <0.001 | 0 (0%) |
| Female | 933 (62.45%) | 670 (71.81%) | 263 (28.19%) | | 895 (95.93%) | 38 (4.07%) | | |
| Male | 561 (37.55%) | 288 (51.34%) | 273 (48.66%) | | 519 (92.51%) | 42 (7.48%) | | |
| **Body mass index** ($kg/m^2$) | 24.11 ± 3.44 | 22.09 ± 1.84 | 27.72 ± 2.57 | <0.001 | 23.64 ± 2.82 | 32.45 ± 2.73 | <0.001 | 0 (0%) |
| **Waist circumference** (cm) | | | | <0.001 | | | <0.001 | 0 (0%) |
| Female | 76.30 ± 8.86 | 72.60 ± 6.15 | 85.56 ± 7.69 | | 75.43 ± 7.81 | 95.74 ± 9.03 | | |
| Male | 87.46 ± 8.88 | 81.98 ± 6.25 | 93.25 ± 7.46 | | 86.12 ± 7.60 | 104.07 ± 6.38 | | |
| **Body fat ratio** (%) | 26.75 ± 7.14 | 24.83 ± 6.29 | 30.18 ± 7.28 | <0.001 | 26.29 ± 6.85 | 34.79 ± 7.37 | <0.001 | 18 (1.20%) |
| **Abdominal fat ratio** (%) | 0.46 ± 0.10 | 0.44 ± 0.09 | 0.52 ± 0.08 | <0.001 | 0.45 ± 0.09 | 0.58 ± 0.02 | <0.001 | 1365 (91.37%) |
| **Systolic blood pressure** (mm Hg) | 131.51 ± 19.06 | 128.56 ± 19.26 | 136.72 ± 17.55 | <0.001 | 131.20 ± 19.27 | 136.82 ± 14.11 | 0.001 | 60 (4.02%) |
| **Diastolic blood pressure** (mm Hg) | 81.55 ± 10.78 | 79.57 ± 10.63 | 85.05 ± 10.15 | <0.001 | 81.32 ± 10.84 | 85.47 ± 8.89 | <0.001 | 60 (4.02%) |
| **Fasting plasma glucose** (mmol/L) | 5.11 ± 0.99 | 5.06 ± 1.03 | 5.18 ± 0.92 | 0.035 | 5.09 ± 0.94 | 5.45 ± 1.64 | 0.087 | 203 (13.59%) |
| **HbA1c** (mmol/mol) | 37.15 ± 5.96 | 36.17 ± 5.50 | 38.88 ± 6.34 | <0.001 | 36.94 ± 5.83 | 40.88 ± 7.04 | <0.001 | 70 (4.69%) |
| **Triglyceride** (mmol/L) | 1.76 ± 1.11 | 1.55 ± 0.98 | 2.12 ± 1.22 | <0.001 | 1.74 ± 1.10 | 2.16 ± 1.08 | 0.001 | 78 (5.22%) |
| **High density lipoprotein** (mmol/L) | | | | <0.001 | | | <0.001 | 203 (13.59%) |
| Female | 1.59 ± 0.37 | 1.67 ± 0.38 | 1.41 ± 0.29 | | 1.60 ± 0.37 | 1.32 ± 0.22 | | |
| Male | 1.25 ± 0.29 | 1.33 ± 0.31 | 1.17 ± 0.24 | | 1.26 ± 0.29 | 1.11 ± 0.19 | | |
| **Hypertension**, case n (%) | 599 (40.31%) | 310 (32.53%) | 289 (54.22%) | <0.001 | 546 (38.83%) | 53 (66.25%) | <0.001 | 8 (0.54%) |
| **Type 2 Diabetes**, case n (%) | 115 (9.26%) | 36 (4.57%) | 79 (17.40%) | <0.001 | 97 (8.23%) | 18 (28.57%) | <0.001 | 252 (16.87%) |
| **Cardiovascular Diseases**, case n (%) | 68 (6.13%) | 29 (4.17%) | 39 (9.42%) | <0.001 | 57 (5.50%) | 11 (15.28%) | 0.003 | 385 (25.77%) |
| **Hyperlipidemia**, case n (%) | 102 (9.19%) | 39 (5.61%) | 63 (15.18%) | <0.001 | 88 (8.48%) | 14 (19.44%) | 0.004 | 384 (25.70%) |
| **Fatty liver**, case n (%) | 2 (0.18%) | 1 (0.14%) | 1 (0.24%) | 1.000 | 2 (0.19%) | 0 (0.00%) | 1.000 | 384 (25.70%) |

(B) Baseline characteristics according to the incident abdominal obesity

| Variables | Total (N=1496) | WC1 (Prevalence = 25.53%) Non-obese (N=1114) | WC1 Obese (N=382) | WC1 P | WC2 (Prevalence = 5.28%) Non-obese (N=1417) | WC2 Obese (N=79) | WC2 P | Missing data, N (%) |
|---|---|---|---|---|---|---|---|---|
| **Age (years)** | 52.45 ± 7.66 | 51.98 ± 7.54 | 53.82 ± 7.85 | <0.001 | 52.41 ± 7.62 | 53.19 ± 8.32 | 0.379 | 0 (0%) |
| **Sex**, n (%) | | | | <0.001 | | | <0.001 | 0 (0%) |
| Female | 935 (62.50%) | 774 (82.78%) | 161 (17.22%) | | 909 (92.22%) | 26 (2.78%) | | |
| Male | 561 (37.50%) | 340 (60.61%) | 221 (39.39%) | | 508 (90.55%) | 53 (9.45%) | | |
| **Body mass index** ($kg/m^2$) | 24.09 ± 3.44 | 22.77 ± 2.37 | 28.04 ± 3.07 | <0.001 | 23.70 ± 2.93 | 31.50 ± 3.59 | <0.001 | 2 (0.13%) |
| **Waist circumference** (cm) | | | | <0.001 | | | <0.001 | 0 (0%) |
| Female | 76.30 ± 8.86 | 73.33 ± 6.04 | 90.54 ± 5.91 | | 75.60 ± 7.84 | 100.77 ± 7.54 | | |
| Male | 87.46 ± 8.88 | 81.85 ± 5.26 | 96.09 ± 5.90 | | 85.65 ± 7.06 | 104.84 ± 4.79 | | |
| **Body fat ratio** (%) | 26.75 ± 7.14 | 25.62 ± 6.68 | 30.03 ± 7.42 | 0.001 | 26.45 ± 6.98 | 32.33 ± 7.78 | <0.001 | 20 (1.34%) |
| **Abdominal fat ratio** (%) | 0.46 ± 0.10 | 0.44 ± 0.09 | 0.51 ± 0.08 | <0.001 | 0.45 ± 0.09 | 0.55 ± 0.08 | 0.016 | 1367 (91.38%) |
| **Systolic blood pressure** (mm Hg) | 131.53 ± 19.06 | 129.23 ± 19.30 | 138.21 ± 16.66 | <0.001 | 131.09 ± 19.20 | 139.29 ± 4.40 | <0.001 | 60 (4.01%) |
| **Diastolic blood pressure** (mm Hg) | 81.52 ± 10.79 | 80.01 ± 10.67 | 85.91 ± 9.93 | <0.001 | 81.26 ± 10.81 | 86.22 ± 9.28 | <0.001 | 60 (4.01%) |
| **Fasting plasma glucose** (mmol/L) | 5.11 ± 1.01 | 5.06 ± 0.98 | 5.27 ± 1.07 | 0.002 | 5.08 ± 0.94 | 5.62 ± 1.72 | 0.013 | 204 (13.64%) |
| **HbA1c** (mmol/mol) | 37.18 ± 6.05 | 36.34 ± 5.74 | 39.67 ± 6.26 | <0.001 | 36.94 ± 5.81 | 41.61 ± 8.36 | <0.001 | 70 (4.68%) |
| **Triglyceride** (mmol/L) | 1.76 ± 1.11 | 1.60 ± 1.00 | 2.23 ± 1.27 | <0.001 | 1.73 ± 1.08 | 2.34 ± 1.36 | <0.001 | 79 (5.28%) |
| **High density lipoprotein** (mmol/L) | | | | <0.001 | | | <0.001 | 204 (13.64%) |
| Female | 1.59 ± 0.37 | 1.64 ± 0.38 | 1.36 ± 0.26 | | 1.60 ± 0.37 | 1.36 ± 0.26 | | |
| Male | 1.25 ± 0.29 | 1.31 ± 0.30 | 1.17 ± 0.23 | | 1.26 ± 0.29 | 1.15 ± 0.23 | | |
| **Hypertension**, case n (%) | 601 (40.39%) | 373 (33.73%) | 228 (59.69%) | <0.001 | 545 (38.68%) | 56 (70.89%) | <0.001 | 8 (0.53%) |
| **Type 2 Diabetes**, case n (%) | 116 (9.33%) | 54 (5.81%) | 62 (19.81%) | <0.001 | 98 (8.31%) | 18 (28.12%) | <0.001 | 253 (16.91%) |
| **Cardiovascular Diseases**, case n (%) | 68 (6.12%) | 39 (4.79%) | 29 (9.80%) | 0.003 | 58 (5.58%) | 10 (14.08%) | 0.009 | 385 (24.74%) |
| **Hyperlipidemia**, case n (%) | 102 (9.17%) | 53 (6.50%) | 49 (16.50%) | <0.001 | 89 (8.55%) | 13 (18.31%) | 0.011 | 384 (25.67%) |
| **Fatty liver**, case n (%) | 2 (0.18%) | 2 (0.25%) | 0 (0.00%) | 1.000 | 2 (0.19%) | 0 (0.00%) | 1.000 | 384 (25.67%) |

Note: BMI, body mass index; WC, waist circumference; BMI25, BMI ≥ 25 kg/m²; BMI30, BMI ≥ 30 kg/m²; BMI40, BMI ≥ 40 kg/m²; WC1, WC ≥ 85 cm for females / 90 cm for males; WC2, WC ≥ 95 cm for females / 100 cm for males.

**Table S3.** Baseline characteristics according to the incident obesity on $REP3_{NHW}$

(A) Baseline characteristics according to the incident general obesity

| Variables | Total (N=391,463) | BMI30 (Prevalence = 24.13%) Non-obese (N=297,015) | Obese (N=94,448) | P | BMI40 (Prevalence = 1.87%) Non-obese (N=384,147) | Obese (N=7316) | P | Missing data, N (%) |
|---|---|---|---|---|---|---|---|---|
| **Age** (years) | 56.77 ± 7.99 | 56.66 ± 8.07 | 57.14 ± 7.75 | <0.001 | 56.79 ± 8.00 | 55.67 ± 7.575 | <0.001 | 0 (0%) |
| **Sex,** n (%) | | | | <0.001 | | | <0.001 | 0 (0%) |
| Female | 211,227 (53.96%) | 162,713 (77.03%) | 48,514 (22.97%) | | 206,314 (97.67%) | 4,913 (2.33%) | | |
| Male | 180,236 (46.04%) | 134,302 (74.51%) | 45,934 (25.49%) | | 177,833 (98.67%) | 2,403 (1.33%) | | |
| **Body mass index** (kg/m$^2$) | 27.39 ± 4.77 | 25.31 ± 2.71 | 33.92 ± 3.85 | <0.001 | 27.07 ± 4.21 | 43.75 ± 3.73 | <0.001 | 0 (0%) |
| **Waist circumference** (cm) | | | | <0.001 | | | <0.001 | 82 (0.02%) |
| Female | 84.53 ± 12.50 | 79.77 ± 8.51 | 100.48 ± 10.29 | | 83.75 ± 11.48 | 117.00 ± 10.04 | | |
| Male | 97.06 ± 11.35 | 92.56 ± 7.89 | 110.16 ± 9.57 | | 96.58 ± 10.60 | 131.87 ± 9.76 | | |
| **Body fat ratio** (%) | 31.34 ± 8.52 | 29.19 ± 7.57 | 38.12 ± 7.75 | <0.001 | 31.05 ± 8.29 | 46.53 ± 6.15 | <0.001 | 5906 (1.51%) |
| **Abdominal fat ratio** (%) | 0.50 ± 0.11 | 0.47 ± 0.11 | 0.59 ± 0.08 | <0.001 | 0.49 ± 0.11 | 0.68 ± 0.06 | <0.001 | 359,556 (91.85%) |
| **Systolic blood pressure** (mm Hg) | 138.04 ± 18.62 | 136.73 ± 18.70 | 142.14 ± 17.77 | <0.001 | 137.94 ± 18.62 | 143.14 ± 17.75 | <0.001 | 22,506 (5.75%) |
| **Diastolic blood pressure** (mm Hg) | 82.22 ± 10.12 | 81.06 ± 9.95 | 85.87 ± 9.77 | <0.001 | 82.11 ± 10.09 | 87.87 ± 10.10 | <0.001 | 22,503 (5.75%) |
| **Fasting plasma glucose** (mmol/L) | 5.12 ± 1.21 | 5.02 ± 1.00 | 5.41 ± 1.67 | <0.001 | 5.10 ± 1.18 | 5.84 ± 2.25 | <0.001 | 50,117 (12.80%) |
| **HbA1c** (mmol/mol) | 35.95 ± 6.53 | 35.19 ± 5.42 | 38.33 ± 8.74 | <0.001 | 35.83 ± 6.34 | 41.81 ± 11.43 | <0.001 | 18,275 (4.67%) |
| **Triglyceride** (mmol/L) | 1.75 ± 1.02 | 1.62 ± 0.94 | 2.17 ± 1.16 | <0.001 | 1.75 ± 1.02 | 2.15 ± 1.08 | <0.001 | 18,465 (4.72%) |
| **High density lipoprotein** (mmol/L) | 1.45 ± 0.38 | | | <0.001 | | | <0.001 | 49,867 (12.74%) |
| Female | 1.60 ± 0.38 | 1.66 ± 0.38 | 1.40 ± 0.31 | | 1.61 ± 0.38 | 1.29 ± 0.27 | | |
| Male | 1.28 ± 0.31 | 1.33 ± 0.32 | 1.15 ± 0.26 | | 1.29 ± 0.31 | 1.07 ± 0.23 | | |
| **Hypertension**, case n (%) | 205,226 (52.80%) | 139,934 (47.48%) | 65,292 (69.49%) | <0.001 | 205,226 (52.80%) | 199,389 (52.28%) | <0.001 | 2792 (0.71%) |
| **Type 2 Diabetes**, case n (%) | 24,814 (7.57%) | 10,801 (4.36%) | 14,013 (17.54%) | <0.001 | 24,814 (7.57%) | 22,650 (7.05%) | <0.001 | 63,829 (16.31%) |
| **Cardiovascular Diseases**, case n (%) | 40,084 (12.46%) | 25,924 (10.80%) | 14,160 (17.29%) | <0.001 | 40,084 (12.46%) | 38,864 (12.33%) | <0.001 | 69,657 (17.79%) |
| **Hyperlipidemia**, case n (%) | 40,426 (12.53%) | 25,230 (10.49%) | 15,196 (18.51%) | <0.001 | 40,426 (12.53%) | 39,014 (12.35%) | <0.001 | 68,921 (17.61%) |
| **Fatty liver**, case n (%) | 757 (0.23%) | 509 (0.21%) | 248 (0.30%) | <0.001 | 757 (0.23%) | 738 (0.23%) | 0.460 | 68,921 (17.61%) |

(B) Baseline characteristics according to the incident abdominal obesity

| Variables | Total (N=392,065) | WC1 (Prevalence = 58.37%) Non-obese (N=163,226) | WC1 Obese (N=228,839) | WC1 P | WC2 (Prevalence = 27.95%) Non-obese (N=282,466) | WC2 Obese (N=109,599) | WC2 P | Missing data, N (%) |
|---|---|---|---|---|---|---|---|---|
| **Age** (years) | 56.774 (7.994) | 55.705 (8.135) | 57.537 (7.804) | <0.001 | 56.36 ± 8.09 | 57.84 ± 7.65 | <0.001 | 0 (0%) |
| **Sex,** n (%) | | | | <0.001 | | | <0.001 | 0 (0%) |
| Female | 211,491 (53.94%) | 117,332 (55.48%) | 94,159 (44.52%) | | 169,293 (80.05%) | 42,198 (19.95%) | | |
| Male | 180,574 (46.06%) | 45,894 (25.42%) | 134,680 (74.58%) | | 113,173 (62.67%) | 67,401 (37.33%) | | |
| **Body mass index** ($kg/m^2$) | 27.384 (4.765) | 23.856 (2.485) | 29.904 (4.390) | <0.001 | 25.38 ± 3.05 | 32.57 ± 4.48 | <0.001 | 684 (0.17%) |
| **Waist circumference** (cm) | | | | <0.001 | | | <0.001 | 0 (0%) |
| Female | 84.53 ± 12.50 | 75.61 ± 5.63 | 95.65 ± 9.42 | | 79.74 ± 7.93 | 103.77 ± 8.28 | | |
| Male | 97.06 ± 11.35 | 83.94 ± 4.50 | 101.53 ± 9.34 | | 90.23 ± 6.32 | 108.53 ± 8.27 | | |
| **Body fat ratio** (%) | 31.338 (8.515) | 28.779 (7.729) | 33.170 (8.579) | <0.001 | 29.70 ± 8.02 | 35.58 ± 8.29 | <0.001 | 6488 (1.65%) |
| **Abdominal fat ratio** (%) | 0.495 (0.112) | 0.458 (0.113) | 0.531 (0.100) | <0.001 | 0.48 ± 0.11 | 0.57 ± 0.09 | <0.001 | 360,142 (91.86%) |
| **Systolic blood pressure** (mm Hg) | 138.039 (18.624) | 133.676 (18.844) | 141.134 (17.830) | <0.001 | 136.30 ± 18.73 | 142.50 ± 17.59 | <0.001 | 22,557 (5.75%) |
| **Diastolic blood pressure** (mm Hg) | 82.218 (10.119) | 79.125 (9.700) | 84.412 (9.833) | <0.001 | 80.88 ± 9.90 | 85.66 ± 9.86 | <0.001 | 22,554 (5.75%) |
| **Fasting plasma glucose** (mmol/L) | 5.117 (1.211) | 4.945 (0.852) | 5.238 (1.398) | <0.001 | 5.00 ± 0.95 | 5.41 ± 1.67 | <0.001 | 50,199 (12.80%) |
| **HbA1c** (mmol/mol) | 35.949 (6.533) | 34.586 (4.466) | 36.921 (7.526) | <0.001 | 35.04 ± 5.16 | 38.28 ± 8.75 | <0.001 | 18,300 (4.67%) |
| **Triglyceride** (mmol/L) | 1.75 (1.023) | 1.36 (0.726) | 2.03 (1.108) | <0.001 | 1.58 ± 0.90 | 2.21 ± 1.17 | <0.001 | 18,499 (4.72%) |
| **High density lipoprotein** (mmol/L) | | | | <0.001 | | | <0.001 | 49,949 (12.74%) |
| Female | 1.60 ± 0.38 | 1.71 ± 0.38 | 1.46 ± 0.33 | | 1.65 ± 0.37 | 1.39 ± 0.31 | | |
| Male | 1.28 ± 0.31 | 1.43 ± 0.34 | 1.24 ± 0.29 | | 1.35 ± 0.32 | 1.18 ± 0.27 | | |
| **Hypertension**, case n (%) | 205,637 (52.83%) | 62,670 (38.75%) | 142,967 (62.83%) | <0.001 | 129,172 (46.10%) | 76,465 (70.10%) | <0.001 | 2,794 (0.71%) |
| **Type 2 Diabetes**, case n (%) | 24,915 (7.59%) | 2991 (2.21%) | 21,924 (11.38%) | <0.001 | 8751 (3.72%) | 16,164 (17.41%) | <0.001 | 63,925 (16.30%) |
| **Cardiovascular Diseases**, case n (%) | 40,245 (12.48%) | 9704 (7.48%) | 30,541 (15.86%) | <0.001 | 22,643 (9.94%) | 17,602 (18.62%) | <0.001 | 69,683 (17.77%) |
| **Hyperlipidemia**, case n (%) | 40,409 (12.53%) | 9083 (6.99%) | 31,326 (16.26%) | <0.001 | 21,996 (9.65%) | 18,413 (19.49%) | <0.001 | 69,601 (17.75%) |
| **Fatty liver**, case n (%) | 755 (0.23%) | 191 (0.15%) | 564 (0.29%) | <0.001 | 400 (0.18%) | 355 (0.38%) | <0.001 | 69,601 (17.75%) |

Note: BMI, body mass index; WC, waist circumference; BMI30, BMI ≥ 30 kg/m²; BMI40, BMI ≥ 40 kg/m²; WC1, WC ≥ 85 cm for females / 90 cm for males; WC2, WC ≥ 95 cm for females / 100 cm for males.

**Table S4.** Significant SNPs identified from GWASs of obesity-related indicators

| CHR | BP | SNP | REF/ ALT | MAF | GENE | TRAIT | BETA | STAT | P | Replication P | | | | |
|---|---|---|---|---|---|---|---|---|---|---|---|---|---|---|
| | | | | | | | | | | **$REP3_{EUR}$** | **$REP2_{Chi}$** | **$REP3_{EUR}$** | **BBJ** | **TWB** |
| 1 | 177873210 | rs574367 | G/T | 0.25 | *SEC16B** | BMI | 0.06 | 11.02 | $3.03\times10^{-28}$ | **$2.18\times10^{-5}$** | 0.82 | **$2.48\times10^{-70}$** | **$5.32\times10^{-29}$** | **$4.79\times10^{-18}$** |
| | | | | | | WC | 0.04 | 7.49 | $7.15\times10^{-14}$ | **$2.33\times10^{-3}$** | 0.86 | **$4.89\times10^{-50}$** | - | **$3.98\times10^{-11}$** |
| 2 | 632101 | rs11127486 | T/C | 0.09 | *TMEM18** | BMI | -0.07 | -8.45 | $2.94\times10^{-17}$ | - | 0.70 | **$1.23\times10^{-75}$** | **$1.90\times10^{-16}$** | **$1.12\times10^{-22}$** |
| | 25133148 | rs6744205 | T/C | 0.45 | *ADCY3** | BMI | 0.04 | 8.9 | $5.52\times10^{-19}$ | **$5.22\times10^{-4}$** | - | - | **$1.39\times10^{-10}$** | **$3.47\times10^{-17}$** |
| | 25400559 | rs13022041 | G/T | 0.31 | *POMC* | BMI | 0.03 | 5.66 | $1.51\times10^{-8}$ | 0.44 | - | - | **$6.09\times10^{-4}$** | **0.02** |
| | 54240105 | rs6715161 | C/T | 0.01 | *ACYP2* | BMI | -0.17 | -6.99 | $2.70\times10^{-12}$ | - | - | **$9.32\times10^{-5}$** | - | - |
| | 169129145 | rs1955337 | G/T | 0.30 | *STK39* | BMI | 0.04 | 7.04 | $1.96\times10^{-12}$ | 0.86 | 0.99 | 0.12 | **$1.16\times10^{-9}$** | **$7.41\times10^{-6}$** |
| | 181361163 | rs1453053 | A/G | 0.31 | *SCHLAP1* | BMI | -0.03 | -5.73 | $9.80\times10^{-9}$ | 0.60 | - | - | **$9.81\times10^{-6}$** | **$2.24\times10^{-4}$** |
| | 632101 | rs11127486 | T/C | 0.09 | *TMEM18** | WC | -0.05 | -6.27 | $3.74\times10^{-10}$ | - | 0.90 | **$6.57\times10^{-59}$** | - | **$1.55\times10^{-8}$** |
| | 169129145 | rs1955337 | G/T | 0.30 | *STK39* | WC | 0.03 | 6.77 | $1.30\times10^{-11}$ | 0.96 | 0.30 | **0.02** | - | **$3.39\times10^{-4}$** |
| | 227231563 | rs72617133 | C/G | 0.34 | *LOC646736* | WC | 0.03 | 5.69 | $1.29\times10^{-8}$ | 0.73 | - | - | - | 0.26 |
| 3 | 52893465 | rs6798941 | C/T | 0.37 | *TMEM110* | BMI | 0.04 | 8.38 | $5.39\times10^{-17}$ | **$5.60\times10^{-3}$** | - | - | **$8.94\times10^{-8}$** | **$8.51\times10^{-5}$** |
| 4 | 45182527 | rs10938397 | A/G | 0.28 | *GNPDA2** | BMI | 0.04 | 6.84 | $7.76\times10^{-12}$ | 0.07 | 0.19 | **$6.11\times10^{-41}$** | **$7.98\times10^{-13}$** | **$1.10\times10^{-19}$** |
| | | | | | | WC | 0.03 | 5.77 | $8.19\times10^{-9}$ | **0.04** | 0.35 | **$8.14\times10^{-29}$** | - | **$3.89\times10^{-12}$** |
| 5 | 87970352 | rs112862634 | G/C | 0.50 | *LINC00461* | BMI | 0.03 | 6.16 | $7.51\times10^{-10}$ | 0.21 | - | - | **$1.76\times10^{-13}$** | **$1.10\times10^{-7}$** |
| | 95835574 | rs7713143 | C/T | 0.44 | *LOC101929710** | BMI | 0.03 | 5.9 | $3.54\times10^{-9}$ | 0.07 | - | - | **$1.03\times10^{-21}$** | **$5.13\times10^{-14}$** |
| | 122750847 | rs6595447 | T/C | 0.32 | *CEP120* | BMI | -0.03 | -6.1 | $1.05\times10^{-9}$ | **$4.78\times10^{-3}$** | - | - | **$2.99\times10^{-6}$** | **$9.77\times10^{-3}$** |
| 6 | 20686878 | rs67131976 | C/T | 0.46 | *CDKAL1** | BMI | -0.03 | -6.76 | $1.35\times10^{-11}$ | **0.05** | 0.20 | **$1.06\times10^{-6}$** | $1.32\times10^{-28}$ | **$2.82\times10^{-5}$** |
| | | | | | | WC | -0.02 | -5.45 | $4.95\times10^{-8}$ | 0.32 | 0.10 | **$1.06\times10^{-4}$** | - | 0.14 |
| | 28782998 | rs16897420 | C/T | 0.38 | *LINC01623* | BMI | -0.03 | -5.72 | $1.09\times10^{-8}$ | - | 0.79 | 0.07 | - | **$2.09\times10^{-3}$** |
| 7 | 138817193 | rs11525873 | T/C | 0.29 | *TTC26* | BMI | -0.03 | -5.84 | $5.40\times10^{-9}$ | - | 0.51 | **$9.05\times10^{-8}$** | **$6.26\times10^{-4}$** | **$5.25\times10^{-3}$** |
| 10 | 31800583 | **rs220063** | A/G | 0.19 | ***ZEB1*** | BMI | -0.03 | -5.49 | $3.95\times10^{-8}$ | - | - | - | **$9.30\times10^{-4}$** | **$1.55\times10^{-4}$** |
| | 77437044 | rs2454812 | T/C | 0.48 | *LRMDA* | BMI | -0.03 | -5.83 | $5.47\times10^{-9}$ | 0.62 | - | - | **0.01** | 0.93 |
| | 104222963 | rs12570201 | T/C | 0.25 | *MFSD13A* | BMI | -0.03 | -5.89 | $3.85\times10^{-9}$ | 0.28 | 0.66 | **$5.25\times10^{-4}$** | **0.02** | 0.12 |
| 11 | 8612000 | rs4418812 | A/G | 0.41 | *STK33* | BMI | 0.03 | 5.68 | $1.32\times10^{-8}$ | 0.97 | 0.47 | **$3.32\times10^{-8}$** | **$1.53\times10^{-6}$** | **$8.32\times10^{-5}$** |
| | 27704209 | rs34379767 | G/A | 0.46 | *BDNF-AS** | BMI | -0.05 | -9.84 | $8.08\times10^{-23}$ | **$2.75\times10^{-3}$** | 0.52 | **$4.13\times10^{-45}$** | **$5.97\times10^{-25}$** | **$4.90\times10^{-14}$** |
| | 27650524 | rs10742179 | A/G | 0.50 | *BDNF-AS* | WC | 0.03 | 6.95 | $3.58\times10^{-12}$ | - | - | - | - | **$3.80\times10^{-5}$** |

| | | | | | | | | | | | | | | |
|---|---|---|---|---|---|---|---|---|---|---|---|---|---|---|
| 12 | 111684461 | rs59214806 | A/G | 0.49 | *CUX2* | WC | 0.03 | 6.42 | $1.38\times10^{-10}$ | 0.37 | - | - | - | 0.14 |
| | 111963570 | rs11065939 | T/C | 0.47 | *ATXN2* | WC | 0.03 | 5.79 | $6.92\times10^{-9}$ | 0.37 | - | - | - | 0.18 |
| 13 | 54084032 | rs4883723 | G/A | 0.28 | *LINC00558* | BMI | 0.04 | 6.85 | $7.45\times10^{-12}$ | - | - | - | **$1.89\times10^{-7}$** | **$3.55\times10^{-7}$** |
| | | | | | | WC | 0.03 | 5.53 | $3.21\times10^{-8}$ | - | - | - | - | **$3.80\times10^{-3}$** |
| 16 | 20037123 | rs57705530 | G/A | 0.39 | *GPR139* | BMI | 0.03 | 5.46 | $4.66\times10^{-8}$ | **0.02** | 0.28 | **$2.44\times10^{-8}$** | **$8.30\times10^{-7}$** | **$1.17\times10^{-5}$** |
| | 20257039 | rs4420537 | C/T | 0.22 | *GP2** | BMI | -0.04 | -6.45 | $1.11\times10^{-10}$ | 0.08 | - | - | **$7.87\times10^{-11}$** | **$7.24\times10^{-18}$** |
| | 30093779 | rs2278557 | C/G | 0.32 | *PPP4C* | BMI | 0.03 | 6.68 | $2.39\times10^{-11}$ | **$7.86\times10^{-3}$** | 0.53 | **$4.53\times10^{-25}$** | **$1.98\times10^{-5}$** | **$9.55\times10^{-11}$** |
| | 53839135 | rs8044769 | C/T | 0.39 | *FTO** | BMI | -0.03 | -5.59 | $2.30\times10^{-8}$ | - | - | - | **$3.21\times10^{-11}$** | **$2.14\times10^{-8}$** |
| | 53845487 | rs11642841 | C/A | 0.03 | *FTO** | BMI | 0.09 | 7.07 | $1.57\times10^{-12}$ | - | - | - | **$7.69\times10^{-32}$** | **$7.76\times10^{-13}$** |
| | | | | | | WC | 0.08 | 6.1 | $1.05\times10^{-9}$ | - | - | - | - | **$1.74\times10^{-11}$** |
| | 30068354 | rs9939774 | C/T | 0.32 | *ALDOA* | WC | 0.03 | 6.48 | $8.96\times10^{-11}$ | **0.04** | 0.88 | **$2.60\times10^{-30}$** | - | **$1.35\times10^{-9}$** |
| 17 | 79047278 | rs35867081 | G/A | 0.36 | *BAIAP2* | BMI | 0.03 | 5.66 | $1.50\times10^{-8}$ | - | - | - | **$1.16\times10^{-3}$** | **$2.63\times10^{-7}$** |
| | 46143042 | **rs7209484** | T/C | 0.26 | ***NFE2L1*** | WC | -0.03 | -5.8 | $6.77\times10^{-9}$ | - | - | - | - | **$1.17\times10^{-4}$** |
| 18 | 57852587 | rs476828 | T/C | 0.27 | *MC4R** | BMI | 0.07 | 12.26 | $1.65\times10^{-34}$ | **0.01** | 0.75 | **$5.14\times10^{-85}$** | **$2.05\times10^{-29}$** | **$1.91\times10^{-16}$** |
| | | | | | | WC | 0.05 | 9.67 | $4.07\times10^{-22}$ | **0.03** | 0.54 | **$4.54\times10^{-78}$** | **$3.97\times10^{-5}$** | **$5.62\times10^{-5}$** |
| | 63441752 | rs8096722 | C/G | 0.31 | *CDH7* | BMI | 0.03 | 5.96 | $2.57\times10^{-9}$ | 0.93 | - | - | - | **$9.77\times10^{-12}$** |
| 19 | 46177235 | rs55669001 | C/T | 0.47 | *GIPR** | BMI | 0.04 | 8.25 | $1.60\times10^{-16}$ | - | - | - | **$5.56\times10^{-39}$** | **$9.12\times10^{-7}$** |
| | 46267453 | rs16980013 | G/T | 0.20 | *BHMG1* | BMI | 0.03 | 5.48 | $4.38\times10^{-8}$ | 0.50 | 0.70 | 0.72 | **$3.24\times10^{-12}$** | **0.01** |
| | 47588325 | rs2287842 | G/A | 0.21 | *ZC3H4* | BMI | 0.04 | 6.34 | $2.34\times10^{-10}$ | **$9.58\times10^{-3}$** | 0.06 | **$2.38\times10^{-8}$** | - | **$6.31\times10^{-8}$** |
| | 46221726 | **rs11881883** | G/A | 0.40 | ***FBXO46**** | WC | -0.03 | -6.82 | $9.10\times10^{-12}$ | - | - | - | - | **$4.17\times10^{-7}$** |
| 20 | 60574216 | rs2296083 | T/C | 0.31 | *TAF4* | BMI | 0.03 | 5.69 | $1.26\times10^{-8}$ | **0.05** | 0.28 | 0.23 | **$7.15\times10^{-8}$** | **$8.51\times10^{-5}$** |
| | 57469073 | rs6026584 | C/T | 0.35 | *GNAS* | WC | -0.03 | -5.6 | $2.18\times10^{-8}$ | 0.89 | 0.40 | **$8.89\times10^{-4}$** | - | **$8.91\times10^{-3}$** |
| 21 | 40309591 | **rs4817972** | C/G | 0.32 | ***LOC400867*** | BMI | 0.03 | 5.66 | $1.48\times10^{-8}$ | 0.79 | - | - | **$2.47\times10^{-4}$** | **$1.20\times10^{-8}$** |

Note: BMI, body mass index; WC, waist circumference; BBJ, Biobank Japan; TWB, Taiwan Biobank. * indicates gene regions that do not contain genome-wide significant SNPs associated with obesity, as defined by the obesity-related indicators listed in the TRAIT column.

**Table S5.** Significant SNPs identified from GWAS of obesity-related diseases

| CHR | BP | SNP | REF/ ALT | MAF | GENE | TRAIT | BETA | STAT | P | Replication P | | | | |
|---|---|---|---|---|---|---|---|---|---|---|---|---|---|---|
| | | | | | | | | | | REP1$_{Kor}$ | REP2$_{Chi}$ | REP3$_{EUR}$ | BBJ | TWB |
| 1 | 10799577 | rs12046278 | C/T | 0.24 | *CASZ1* | HT | 0.92 | -5.47 | $4.51\times10^{-8}$ | - | - | - | - | - |
| | 113046879 | rs3790604 | C/A | 0.30 | *WNT2B* | HT | 1.1 | 6.7 | $2.07\times10^{-11}$ | - | - | **$2.18\times10^{-16}$** | - | - |
| 2 | 26929282 | rs13394970 | T/G | 0.22 | *KCNK3* | HT | 0.86 | -9.12 | $7.45\times10^{-20}$ | - | - | - | - | - |
| | 54774112 | rs886213800 | C/A | 0.04 | *SPTBN1* | HT | 1.21 | 5.49 | $4.06\times10^{-8}$ | - | - | - | - | - |
| | 54924901 | rs138938305 | C/T | 0.03 | *SPTBN1* | HT | 1.35 | 7.93 | $2.14\times10^{-15}$ | - | - | - | - | - |
| | 165132714 | rs1561467 | T/A | 0.33 | *GRB14* | HT | 0.92 | -5.56 | $2.63\times10^{-8}$ | - | - | - | - | - |
| 3 | 53548031 | rs59968724 | C/G | 0.11 | *CACNA1D* | HT | 1.13 | 5.78 | $7.35\times10^{-9}$ | - | - | - | - | - |
| | 53597491 | rs807191 | A/C | 0.47 | *CACNA1D* | HT | 1.09 | 6.5 | $7.82\times10^{-11}$ | - | - | - | - | - |
| 4 | 81155937 | rs79620921 | A/T | 0.26 | *PRDM8* | HT | 1.12 | 7.81 | $5.76\times10^{-15}$ | - | - | - | - | - |
| | 81166686 | rs140801183 | A/G | 0.02 | *FGF5* | HT | 1.29 | 5.8 | $6.71\times10^{-9}$ | - | - | - | - | - |
| | 81207963 | rs3733336 | A/G | 0.21 | *FGF5* | HT | 1.17 | 9.86 | $6.04\times10^{-23}$ | - | - | - | - | - |
| 5 | 32829929 | rs1177765 | C/T | 0.39 | *NPR3* | HT | 0.92 | -5.89 | $3.92\times10^{-9}$ | - | - | **$3.51\times10^{-27}$** | - | - |
| | 114409933 | rs4435857 | A/G | 0.44 | *TRIM36* | HT | 1.08 | 5.78 | $7.31\times10^{-9}$ | - | - | - | - | - |
| | 122460332 | rs2287696 | A/G | 0.43 | *PRDM6* | HT | 0.92 | -6.15 | $7.66\times10^{-10}$ | - | - | - | - | - |
| 8 | 143988986 | rs62525983 | C/G | 0.32 | *CYP11B2* | HT | 0.9 | -7.26 | $3.93\times10^{-13}$ | - | - | - | - | - |
| 10 | 104386309 | rs4285804 | T/A | 0.26 | *SUFU* | HT | 1.09 | 5.92 | $3.20\times10^{-9}$ | - | - | 0.09 | - | - |
| | 104928914 | rs12416331 | T/A | 0.24 | *NT5C2* | HT | 0.88 | -8.13 | $4.15\times10^{-16}$ | - | - | - | - | - |
| 11 | 116882638 | rs4245168 | C/A | 0.5 | *SIK3* | HT | 0.93 | -5.78 | $7.61\times10^{-9}$ | - | - | - | - | - |
| 12 | 90058842 | rs111478946 | G/A | 0.37 | *ATP2B1* | HT | 0.87 | -9.92 | $3.53\times10^{-23}$ | - | - | **$6.22\times10^{-29}$** | - | - |
| | 111465104 | rs7980382 | C/T | 0.22 | *CUX2* | HT | 0.9 | -6.27 | $3.56\times10^{-10}$ | - | - | - | - | - |
| | 113164710 | rs34560882 | C/A | 0.39 | *RPH3A* | HT | 0.93 | -5.76 | $8.43\times10^{-9}$ | - | - | - | - | - |
| | 113713062 | rs11066525 | A/C | 0.07 | *TPCN1* | HT | 0.87 | -5.58 | $2.46\times10^{-8}$ | - | - | - | - | - |
| | 115553034 | rs35442 | G/A | 0.24 | *TBX3* | HT | 0.89 | -7.54 | $4.76\times10^{-14}$ | - | - | - | - | - |
| 13 | 30146201 | rs9508495 | C/T | 0.35 | *SLC7A1* | HT | 0.91 | -6.48 | $9.19\times10^{-11}$ | - | - | - | - | - |
| 17 | 78358945 | rs112735431 | G/A | 0.01 | *RNF213* | HT | 1.77 | 10.05 | $8.92\times10^{-24}$ | - | - | - | - | - |
| 19 | 11526765 | rs167479 | G/T | 0.49 | *RGL3* | HT | 0.91 | -7.6 | $3.06\times10^{-14}$ | - | - | - | - | - |
| 20 | 10965998 | rs1887320 | A/G | 0.48 | *C20orf187* | HT | 0.9 | -7.93 | $2.18\times10^{-15}$ | - | - | **$1.98\times10^{-26}$** | - | - |

| | | | | | | | | | | | | | | |
|---|---|---|---|---|---|---|---|---|---|---|---|---|---|---|
| 2 | 234346270 | rs117447187 | T/C | 0.03 | *DGKD* | T2D | 1.39 | 6.05 | $1.46\times10^{-9}$ | - | - | - | - | - |
| 3 | 23298683 | rs9866739 | C/T | 0.19 | *UBE2E2* | T2D | 0.86 | -5.89 | $3.80\times10^{-9}$ | - | - | **$1.97\times10^{-4}$** | **$6.82\times10^{-20}$** | - |
| | 63826364 | rs73120894 | T/C | 0.26 | *C3orf49* | T2D | 0.88 | -5.52 | $3.39\times10^{-8}$ | - | - | **0.02** | **$1.39\times10^{-12}$** | - |
| | 187121145 | rs79282614 | A/G | 0.01 | *RTP4* | T2D | 1.53 | 5.69 | $1.24\times10^{-8}$ | - | - | - | - | - |
| 6 | 20682622 | rs35612982 | T/C | 0.46 | *CDKAL1** | T2D | 1.32 | 14.35 | $1.10\times10^{-46}$ | - | - | - | **$1.93\times10^{-91}$** | - |
| 7 | 127253550 | rs2233580 | C/T | 0.07 | *PAX4* | T2D | 1.42 | 10.41 | $2.28\times10^{-25}$ | - | - | - | **$6.30\times10^{-90}$** | - |
| 8 | 118184783 | rs13266634 | C/T | 0.40 | *SLC30A8* | T2D | 0.87 | -7.12 | $1.07\times10^{-12}$ | - | - | **$4.74\times10^{-27}$** | **$1.22\times10^{-37}$** | - |
| 9 | 22134068 | rs10811660 | G/A | 0.44 | *CDKN2B-AS1* | T2D | 0.79 | -11.59 | $4.59\times10^{-31}$ | - | - | **$3.23\times10^{-26}$** | **$5.37\times10^{-98}$** | - |
| 10 | 12309139 | rs11257657 | C/G | 0.45 | *CDC123* | T2D | 1.14 | 6.87 | $6.59\times10^{-12}$ | - | - | - | **$2.71\times10^{-32}$** | - |
| | 94150879 | rs12260406 | G/A | 0.23 | *MARK2P9* | T2D | 1.14 | 5.82 | $5.84\times10^{-9}$ | - | - | - | **$1.82\times10^{-9}$** | - |
| | 94466439 | rs12219514 | G/A | 0.15 | *HHEX* | T2D | 1.23 | 8.1 | $5.28\times10^{-16}$ | - | - | **$9.99\times10^{-23}$** | **$1.05\times10^{-41}$** | - |
| | 94735056 | rs549156247 | G/G | 0.04 | *EXOC6* | T2D | 1.32 | 6.03 | $1.59\times10^{-9}$ | - | - | - | **$4.76\times10^{-21}$** | - |
| 18 | 57808978 | rs66723169 | C/A | 0.23 | *MC4R** | T2D | 1.14 | 5.9 | $3.73\times10^{-9}$ | - | - | - | **$8.76\times10^{-13}$** | - |
| 20 | 42972856 | rs8115481 | G/T | 0.50 | *R3HDML* | T2D | 0.89 | -5.88 | $4.17\times10^{-9}$ | - | - | - | **$8.80\times10^{-6}$** | - |
| 17 | 78358945 | rs112735431 | G/A | 0.01 | *RNF213* | CVD | 2.04 | 7.5 | $6.42\times10^{-14}$ | - | - | - | - | - |
| 1 | 109822166 | rs599839 | A/G | 0.06 | *PSRC1* | HL | 0.78 | -6.78 | $1.19\times10^{-11}$ | - | - | - | - | - |
| 2 | 20849856 | rs116948525 | G/C | 0.04 | *HS1BP3* | HL | 1.24 | 5.74 | $9.74\times10^{-9}$ | - | - | - | - | - |
| | 21195522 | rs72788559 | G/T | 0.13 | *APOB* | HL | 0.84 | -6.67 | $2.61\times10^{-11}$ | - | - | **$6.38\times10^{-3}$** | - | - |
| | 21242731 | rs13306206 | G/A | 0.01 | *APOB* | HL | 1.87 | 9.48 | $2.47\times10^{-21}$ | - | - | - | - | - |
| | 27742603 | rs780093 | T/C | 0.46 | *GCKR* | HL | 0.91 | -5.89 | $3.84\times10^{-9}$ | - | - | - | - | - |
| 5 | 74617262 | rs2878417 | A/G | 0.48 | *HMGCR* | HL | 0.9 | -6.68 | $2.45\times10^{-11}$ | - | - | **$5.26\times10^{-16}$** | - | - |
| 8 | 126482621 | rs2954022 | A/C | 0.44 | *TRIB1* | HL | 1.15 | 8.2 | $2.33\times10^{-16}$ | - | - | **$4.28\times10^{-30}$** | - | - |
| 11 | 116639692 | **rs1268353** | C/T | 0.22 | *BUD13* | HL | 0.88 | -6.17 | $6.75\times10^{-10}$ | - | - | **$1.09\times10^{-5}$** | - | - |
| | 116707684 | rs2070665 | G/A | 0.36 | *APOA1-AS* | HL | 1.11 | 6.03 | $1.68\times10^{-9}$ | - | - | - | - | - |
| | 116879519 | rs12284696 | A/G | 0.17 | *SIK3* | HL | 0.88 | -5.72 | $1.08\times10^{-8}$ | - | - | 0.46 | - | - |
| 16 | 72009439 | **rs6499554** | G/A | 0.30 | *PKD1L3* | HL | 0.9 | -5.79 | $7.03\times10^{-9}$ | - | - | **$2.62\times10^{-4}$** | - | - |
| | 72214276 | rs11643192 | C/A | 0.30 | *PMFBP1* | HL | 1.1 | 5.55 | $2.88\times10^{-8}$ | - | - | - | - | - |
| 18 | 47142243 | rs17726130 | G/A | 0.28 | *LIPG* | HL | 1.12 | 6.19 | $6.19\times10^{-10}$ | - | - | 0.50 | - | - |
| 19 | 11242307 | rs2738464 | C/G | 0.29 | *LDLR* | HL | 0.88 | -6.99 | $2.68\times10^{-12}$ | - | - | - | - | - |
| | 11346155 | rs56322906 | G/A | 0.27 | *DOCK6* | HL | 0.89 | -6.1 | $1.05\times10^{-9}$ | - | - | - | - | - |

| | | | | | | | | | | | | | | |
|---|---|---|---|---|---|---|---|---|---|---|---|---|---|---|
| | 45412079 | rs7412 | C/T | 0.06 | *APOE* | HL | 0.69 | -9.74 | $2.04\times10^{-22}$ | **-** | **-** | **$1.29\times10^{-86}$** | - | **-** |
| 22 | 44324727 | rs738409 | C/G | 0.42 | *PNPLA3* | FL | 1.22 | 8.13 | $4.47\times10^{-16}$ | **-** | **-** | **$5.18\times10^{-10}$** | - | **-** |

Note: HT, hypertension; T2D, type 2 diabetes; CVD, cardiovascular disease; HL, hyperlipidemia; FL, fatty liver; BBJ, Biobank Japan; TWB, Taiwan Biobank. * indicates gene regions that overlap with genome-wide significant SNPs associated with obesity.

**Table S6.** Genomic inflation, SNP heritability, and genetic correlations for obesity-related indicators and obesity-related diseases

(A) Genomic inflation factors and SNP heritability estimates for obesity-related indicators

| Dataset | Estimator | General obesity | | | Abdominal obesity | | |
|---|---|---|---|---|---|---|---|
| | | **BMI** | **BMI25/BMI30** | **BMI30/BMI40** | **WC** | **WC1** | **WC2** |
| Discovery | $\lambda_{GC}$ | 1.26 | 1.16 | 1.06 | 1.17 | 1.11 | 1.05 |
| | $\lambda_{LDSC}$ | 1.04 | 1.03 | 1.00 | 1.03 | 1.05 | 1.01 |
| | $h^2$ (P) | **0.21** ($1.07 \times 10^{-62}$) | **0.13** ($9.99 \times 10^{-38}$) | **0.04** ($2.17 \times 10^{-10}$) | **0.15** ($3.66 \times 10^{-46}$) | **0.09** ($3.27 \times 10^{-24}$) | **0.04** ($2.11 \times 10^{-8}$) |
| $REP1_{Kor}$ | $\lambda_{GC}$ | 1.02 | 1.03 | 1.01 | 1.02 | 1.01 | 1.01 |
| | $\lambda_{LDSC}$ | 1.01 | 1.00 | 0.98 | 1.01 | 1.00 | 1.00 |
| | $h^2$ (P) | **0.19** (0.04) | 0.14 (0.09) | 0.10 (0.16) | **0.15** (0.04) | 0.07 (0.31) | 0 (0.97) |
| $REP2_{Chi}$ | $\lambda_{GC}$ | 0.99 | 0.99 | 1.01 | 0.98 | 0.99 | 1.02 |
| | $\lambda_{LDSC}$ | 1.01 | 1.01 | 1.03 | 1.01 | 1.02 | 1.01 |
| | $h^2$ (P) | -0.37 (0.27) | -0.64 (0.04) | -0.44 (0.22) | -0.48 (0.21) | -0.66 (0.05) | 0 (0.99) |
| $REP3_{NHW}$ | $\lambda_{GC}$ | 1.91 | 1.61 | 1.18 | 1.76 | 1.50 | 1.49 |
| | $\lambda_{LDSC}$ | 1.55 | 1.10 | 1.01 | 1.47 | 1.09 | 1.09 |
| | $h^2$ (P) | **0.20** ($2.44 \times 10^{-92}$) | **0.14** ($1.67 \times 10^{-150}$) | **0.04** ($1.34 \times 10^{-43}$) | **0.16** ($2.55 \times 10^{-89}$) | **0.12** ($2.44 \times 10^{-146}$) | **0.12** ($8.25 \times 10^{-137}$) |

(B) Genomic inflation factors and SNP heritability estimates for obesity-related diseases

| Dataset | Estimator | HT | T2D | CVD | HL | FL |
|---|---|---|---|---|---|---|
| Discovery | $\lambda_{GC}$ | 1.12 | 1.08 | 1.01 | 1.05 | 1.01 |
| | $\lambda_{LDSC}$ | 1.02 | 1.02 | 0.99 | 1.02 | 0.99 |
| | $h^2$ (P) | **0.11** ($9.55 \times 10^{-26}$) | **0.06** ($3.06 \times 10^{-11}$) | **0.01** (0.02) | **0.04** ($2.87 \times 10^{-7}$) | 0.01 (0.09) |
| $REP3_{NHW}$ | $\lambda_{GC}$ | 1.49 | 1.25 | 1.17 | 1.19 | 0.99 |

| | | | | | | |
|---|---|---|---|---|---|---|
| | $\lambda_{LDSC}$ | 1.55 | 1.10 | 1.01 | 1.47 | 1.09 |
| | $h^2$ (P) | **0.20** ($2.44 \times 10^{-92}$) | **0.14** ($1.67 \times 10^{-150}$) | **0.04** ($1.34 \times 10^{-43}$) | **0.16** ($2.55 \times 10^{-89}$) | **-**$8.75\times10^{-5}$ (1.00) |

(C) Pairwise genetic correlations among obesity-related indicators and obesity-related diseases

| **Dataset** | **Estimator** | **HT** | **T2D** | **CVD** | **HL** | **FL** |
|---|---|---|---|---|---|---|
| Discovery | $GC_{BMI25}$ (P) | **0.26** ($1.38 \times 10^{-5}$) | **0.33** ($5.80 \times 10^{-5}$) | **0.30** (0.03) | 0.12 (0.15) | **0.33** (0.02) |
| | $GC_{WC1}$ (P) | **0.30** ($8.10 \times 10^{-6}$) | **0.42** ($2.86 \times 10^{-6}$) | **0.38** (0.03) | **0.19** (0.05) | 0.33 (0.11) |
| $REP3_{NHW}$ | $GC_{BMI30}$ (P) | **0.36** ($5.31 \times 10^{-58}$) | **0.65** ($4.71 \times 10^{-72}$) | **0.45** ($6.98 \times 10^{-47}$) | **0.47** ($2.29 \times 10^{-20}$) | nan (nan) |
| | $GC_{WC1}$ (P) | **0.34** ($2.45 \times 10^{-39}$) | **0.65** ($1.78 \times 10^{-72}$) | **0.42** ($2.25 \times 10^{-34}$) | **0.44** ($2.65 \times 10^{-18}$) | nan (nan) |

Note: BMI, body mass index; BMI25, BMI ≥ 25 kg/m²; BMI30, BMI ≥ 30 kg/m²; BMI40, BMI ≥ 40 kg/m²; WC, waist circumference; WC1, WC ≥ 85 cm for females / 90 cm for males; WC2, WC ≥ 95 cm for females / 100 cm for males; GC, genetic correlation; $\lambda_{GC}$, genomic inflation factor; $\lambda_{LDSC}$., LD score intercept; $h^2$ (P), SNP heritability estimate with P value; $GC_{BMI25/BMI30}$ (P) and $GC_{WC1}$ (P), genetic correlations between obesity and obesity related diseases, with corresponding P values in parentheses; nan, not available due to out-of-bound heritability estimates.

**Table S7.** Significant genetic regions identified by local genetic correlations between obesity and obesity-related diseases

(A) Significant local genetic correlations between general obesity and obesity-related diseases

| **Disease** | **Discovery dataset** | | | | **Replication dataset of REP3$_{NHW}$** | | | |
|---|---|---|---|---|---|---|---|---|
| | **REGION** | **GC (P)** | **BMI25 $h^2$(P)** | **Disease $h^2$(P)** | **REGION** | **GC (P)** | **BMI30 $h^2$(P)** | **Disease $h^2$(P)** |
| HT | 1:200135728-201173714 | 0.75 (3.07×10$^{-6)}$ | 1.91×10$^{-3}$ (8.57×10$^{-9}$) | 1.98×10$^{-3}$ (1.78×10$^{-8}$) | 1:200134006-201067952 | 0.01 (0.97) | 1.39×10$^{-4}$ (5.80×10$^{-3}$) | 1.73×10$^{-4}$ (1.70×10$^{-3}$) |
| HT | 2:215569986-217243305 | 0.95 (6.51×10$^{-7)}$ | 1.30×10$^{-3}$ (3.10×10$^{-5}$) | 1.65×10$^{-3}$ (2.26×10$^{-5}$) | 2:215899571-217566011 | -0.08 (0.65) | 2.52×10$^{-4}$ (2.83×10$^{-4}$) | 4.85×10$^{-4}$ (3.56×10$^{-10}$) |
| **HT** | **3:8327718-9760484** | 1.00 (6.23×10$^{-7)}$ | 1.20×10$^{-3}$ (1.08×10$^{-4}$) | 1.60×10$^{-3}$ (6.61×10$^{-6}$) | **3:8664893-9970731** | 0.61 (**9.46×10$^{-5}$**) | 5.35×10$^{-4}$ (1.03×10$^{-11}$) | 3.71×10$^{-4}$ (6.84×10$^{-7}$) |
| **HT** | **3:51817883-53541920** | 0.82 (4.94×10$^{-6)}$ | 1.46×10$^{-3}$ (8.07×10$^{-7}$) | 1.74×10$^{-3}$ (1.87×10$^{-7}$) | **3:51953969-54074844** | 0.24 (**0.04**) | 5.05×10$^{-4}$ (5.29×10$^{-11}$) | 7.79×10$^{-4}$ (9.40×10$^{-22}$) |
| **HT** | **7:1355243-2477957** | 0.65 (4.32×10$^{-6)}$ | 1.62×10$^{-3}$ (1.26×10$^{-7}$) | 3.32×10$^{-3}$ (8.44×10$^{-17}$) | **7:1366973-2473749** | 0.34 (**0.01**) | 5.84×10$^{-4}$ (1.82×10$^{-14}$) | 4.57×10$^{-4}$ (6.88×10$^{-10}$) |
| HT | 11:6882946-7618719 | 1.00 (5.30×10$^{-6}$) | 5.87×10$^{-4}$ (0.01) | 1.69×10$^{-3}$ (4.98×10$^{-7}$) | 11:7191231-7915071 | 0.15 (0.56) | 3.29×10$^{-4}$ (2.41×10$^{-7}$) | 1.39×10$^{-4}$ (0.01) |
| HT | 12:109020532-110116508 | 0.86 (4.18×10$^{-7}$) | 1.41×10$^{-3}$ (7.09×10$^{-7}$) | 2.02×10$^{-3}$ (6.60×10$^{-10}$) | 12:108183189-109031819 | 0.02 (0.92) | 2.93×10$^{-4}$ (2.86×10$^{-6}$) | 3.22×10$^{-4}$ (6.26×10$^{-7}$) |
| HT | 12:110116509-112206126 | 0.73 (2.33×10$^{-6}$) | 1.16×10$^{-3}$ (3.12×10$^{-5}$) | 3.50×10$^{-3}$ (2.49×10$^{-21}$) | 12:110111515-111592381 | 0.18 (0.30) | 2.36×10$^{-4}$ (1.11×10$^{-5}$) | 4.12×10$^{-4}$ (7.26×10$^{-12}$) |
| HT | 13:27952091-29255849 | 1.00 (5.40×10$^{-7}$) | 1.74×10$^{-3}$ (4.94×10$^{-8}$) | 1.18×10$^{-3}$ (5.38×10$^{-4}$) | 13:27392006-28587125 | 0.18 (0.24) | 5.30×10$^{-4}$ (8.82×10$^{-13}$) | 3.66×10$^{-4}$ (2.41×10$^{-7}$) |
| HT | 15:25803835-26656298 | 0.47 (2.60×10$^{-6}$) | 2.91×10$^{-3}$ (1.02×10$^{-12}$) | 4.85×10$^{-3}$ (3.65×10$^{-31}$) | 15:25384328-26392947 | 0.33 (0.13) | 3.59×10$^{-4}$ (6.13×10$^{-7}$) | 2.15×10$^{-4}$ (1.20×10$^{-3}$) |
| **HT** | **16:1036951-1994688** | 0.63 (4.75×10$^{-5}$) | 2.11×10$^{-3}$ (1.31×10$^{-10}$) | 2.12 ×10$^{-3}$ (4.26×10$^{-8}$) | **16:1049080-1996492** | 0.46 (**3.63×10$^{-3}$**) | 3.72×10$^{-4}$ (3.43×10$^{-8}$) | 4.70×10$^{-4}$ (1.01×10$^{-10}$) |
| HT | 16:5782847-6446996 | 0.82 (1.72×10$^{-6}$) | 2.16×10$^{-3}$ (4.20×10$^{-10}$) | 1.63×10$^{-3}$ (8.01×10$^{-6}$) | 16:5782969-6446081 | 0.14 (0.24) | 6.33×10$^{-4}$ (2.39×10$^{-17}$) | 5.87×10$^{-4}$ (1.32×10$^{-14}$) |
| HT | 22:48274208-48979272 | 0.96 (3.99×10$^{-6}$) | 1.60×10$^{-3}$ (7.14×10$^{-7}$) | 1.27×10$^{-3}$ (2.17×10$^{-4}$) | 22:48269178-48977538 | 0.24 (0.07) | 5.23×10$^{-4}$ (6.94×10$^{-13}$) | 4.67×10$^{-4}$ (1.26×10$^{-10}$) |
| **T2D** | **1:183930245-184982129** | 1.00 (3.71×10$^{-6}$) | 1.74×10$^{-3}$ (7.10×10$^{-9}$) | 5.69×10$^{-4}$ (0.02) | **1:183920617-184953089** | 0.75 (**1.05×10$^{-3}$**) | 4.12×10$^{-4}$ (4.02×10$^{-10}$) | 1.78×10$^{-4}$ (7.89×10$^{-3}$) |
| T2D | 3:76161795-77157986 | 0.89 (5.21×10$^{-6}$) | 1.21×10$^{-3}$ (1.59×10$^{-5}$) | 9.54×10$^{-4}$ (1.40×10$^{-6}$) | 3:76157471-77507832 | 0.14 (0.67) | 1.97×10$^{-4}$ (8.90×10$^{-4}$) | 1.36×10$^{-4}$ (0.03) |

| | | | | | | | | |
|---|---|---|---|---|---|---|---|---|
| T2D | 10:68374233-69909446 | 0.94 ($1.58\times10^{-5}$) | $1.04\times10^{-3}$ ($2.09\times10^{-4}$) | $1.15\times10^{-3}$ ($5.68\times10^{-6}$) | 10:69904171-71050504 | 0.28 (0.34) | $1.30\times10^{-4}$ (0.02) | $2.78\times10^{-4}$ ($2.31\times10^{-4}$) |
| T2D | 12:110116509-112206126 | 0.85 ($9.57\times10^{-8}$) | $1.16\times10^{-3}$ ($3.12\times10^{-5}$) | $2.74\times10^{-3}$ ($1.24\times10^{-18}$) | 12:110111515-111592381 | 0.29 (0.23) | $2.36\times10^{-4}$ ($1.11\times10^{-5}$) | $1.95\times10^{-4}$ ($1.02\times10^{-3}$) |
| **T2D** | **14:98987578-100039414** | 0.89 ($2.41\times10^{-5}$) | $1.27\times10^{-3}$ ($2.66\times10^{-5}$) | $1.28\times10^{-3}$ ($3.53\times10^{-5}$) | **14:98885323-99474533** | 0.44 (**0.05**) | $2.29\times10^{-4}$ ($8.32\times10^{-5}$) | $2.67\times10^{-4}$ ($3.57\times10^{-5}$) |
| T2D | 15:25803835-26656298 | 0.44 ($3.80\times10^{-5}$) | $2.91\times10^{-3}$ ($1.02\times10^{-17}$) | $1.67\times10^{-3}$ ($3.58\times10^{-22}$) | 15:25384328-26392947 | -0.05 (0.78) | $3.59\times10^{-4}$ ($6.13\times10^{-7}$) | $3.24\times10^{-4}$ $1.58\times10^{-5}$) |
| **T2D** | **18:30265237-32456385** | 1.00 ($2.26\times10^{-6}$) | $1.80\times10^{-3}$ ($2.97\times10^{-8}$) | $4.09\times10^{-3}$ ($7.41\times10^{-3}$) | **18:30401129-32458907** | 0.93 (**$1.24\times10^{-5}$**) | $4.91\times10^{-4}$ ($2.51\times10^{-11}$) | $1.81\times10^{-4}$ ($5.11\times10^{-3}$) |
| T2D | 20:1769210-2663437 | 0.46 ($6.76\times10^{-5}$) | $2.44\times10^{-3}$ ($3.94\times10^{-13}$) | $2.47\times10^{-3}$ ($1.30\times10^{-20}$) | 20:1771286-2668108 | 0.35 (0.14) | $4.66\times10^{-4}$ ($5.15\times10^{-11}$) | $1.44\times10^{-4}$ (0.02) |
| T2D | 22:26790795-27794871 | 0.99 ($2.04\times10^{-6}$) | $7.60\times10^{-4}$ ($5.25\times10^{-3}$) | $2.32\times10^{-3}$ ($3.80\times10^{-12}$) | 22:27192924-27952441 | 0.32 (**0.02**) | $5.32\times10^{-4}$ ($1.97\times10^{-12}$) | $6.03\times10^{-4}$ ($1.34\times10^{-11}$) |
| **CVD** | **2:49913792-50821547** | 0.94 ($3.75\times10^{-6}$) | $1.01\times10^{-3}$ ($1.80\times10^{-4}$) | $1.48\times10^{-3}$ ($1.63\times10^{-7}$) | **2:49849186-50756240** | 0.67 (**$4.03\times10^{-3}$**) | $2.51\times10^{-4}$ ($2.13\times10^{-5}$) | $2.20\times10^{-4}$ ($4.94\times10^{-4}$) |
| CVD | 5:10043-1207104 | -0.61 ($5.91\times10^{-5}$) | $2.15\times10^{-3}$ ($1.35\times10^{-10}$) | $9.75\times10^{-4}$ ($2.73\times10^{-8}$) | 5:1206610-2106885 | 0.23 (0.12) | $4.10\times10^{-4}$ ($1.59\times10^{-8}$) | $5.49\times10^{-4}$ ($8.62\times10^{-11}$) |
| CVD | 6:11228239-12372255 | 0.76 ($1.52\times10^{-5}$) | $1.47\times10^{-3}$ ($1.87\times10^{-6}$) | $1.25\times10^{-3}$ ($3.94\times10^{-8}$) | 6:10416551-11790671 | -0.14 (0.64) | $1.50\times10^{-4}$ (0.01) | $2.68\times10^{-4}$ ($6.53\times10^{-4}$) |
| CVD | 7:36359074-37353248 | 1.00 ($2.23\times10^{-5}$) | $8.33\times10^{-4}$ ($1.60\times10^{-3}$) | $6.73\times10^{-4}$ ($1.33\times10^{-3}$) | 7:36507691-37981936 | 0.08 (0.74) | $3.68\times10^{-4}$ ($1.87\times10^{-7}$) | $2.29\times10^{-4}$ ($3.64\times10^{-3}$) |
| CVD | 13:53609234-55588449 | 0.93 ($1.18\times10^{-6}$) | $1.69\times10^{-3}$ ($9.35\times10^{-8}$) | $9.05\times10^{-4}$ ($6.94\times10^{-5}$) | 13:54684857-55576912 | 0.45 (0.06) | $4.18\times10^{-4}$ ($5.09\times10^{-12}$) | $1.28\times10^{-4}$ (0.01) |
| HP | 3:2147501-3242009 | -0.46 ($3.59\times10^{-5}$) | $3.44\times10^{-3}$ ($1.12\times10^{-20}$) | $2.68\times10^{-3}$ ($8.00\times10^{-15}$) | 3:2140012-3128152 | 0.07 (0.73) | $2.61\times10^{-4}$ ($7.70\times10^{-5}$) | $4.06\times10^{-4}$ ($1.01\times10^{-6}$) |
| HP | 3:72012791-73244159 | 0.98 ($3.60\times10^{-7}$) | $1.58\times10^{-3}$ ($3.58\times10^{-6}$) | $1.48\times10^{-3}$ ($5.29\times10^{-6}$) | 3:72334705-73606612 | 0.24 (0.22) | $2.24\times10^{-4}$ ($7.28\times10^{-4}$) | $5.38\times10^{-4}$ ($1.44\times10^{-9}$) |
| **HP** | **6:105630354-106629704** | 0.94 ($3.45\times10^{-8}$) | $1.63\times10^{-3}$ ($8.01\times10^{-8}$) | $1.79\times10^{-3}$ ($8.61\times10^{-8}$) | **6:104951345-106053915** | 0.58 (**0.05**) | $3.13\times10^{-4}$ ($1.14\times10^{-6}$) | $1.27\times10^{-4}$ (0.04) |
| **HP** | **15:73262649-74454643** | 0.86 ($8.28\times10^{-7}$) | $1.23\times10^{-3}$ ($1.80\times10^{-5}$) | $2.16\times10^{-3}$ ($4.57\times10^{-11}$) | **15:7337571974458113** | 0.36 (**0.05**) | $6.92\times10^{-4}$ ($3.34\times10^{-21}$) | $2.05\times10^{-4}$ ($2.47\times10^{-3}$) |
| FL | 3:186602334-188069213 | 0.91 ($2.85\times10^{-7}$) | $1.77\times10^{-3}$ ($1.58\times10^{-7}$) | $1.29\times10^{-3}$ ($1.53\times10^{-6}$) | - | - | - | - |
| FL | 6:2898448-4319834 | 0.69 ($9.17\times10^{-10}$) | $2.18\times10^{-3}$ ($4.17\times10^{-11}$) | $4.01\times10^{-3}$ ($7.95\times10^{-30}$) | - | - | - | - |
| FL | 6:155739846-157043139 | -1.00 ($7.85\times10^{-6}$) | $7.31\times10^{-4}$ ($7.16\times10^{-3}$) | $9.42\times10^{-4}$ ($7.84\times10^{-6}$) | - | - | - | - |

| | | | | | | | | |
|---|---|---|---|---|---|---|---|---|
| FL | 12:27797119-28936164 | 0.80 ($1.19\times10^{-5}$) | $1.05\times10^{-3}$ ($6.60\times10^{-5}$) | $2.39\times10^{-3}$ ($1.74\times10^{-10}$) | - | - | - | - |

(B) Significant local genetic correlation between abdominal obesity and obesity-related diseases

| Disease | Discovery dataset | | | | Replication dataset of $REP3_{NHW}$ | | | |
|---|---|---|---|---|---|---|---|---|
| | **REGION** | **GC (P)** | **WC1 $h^2$(P)** | **Disease $h^2$(P)** | **REGION** | **GC (P)** | **WC1 $h^2$(P)** | **Disease $h^2$(P)** |
| HT | 2:1376568-2648919 | 0.44 ($4.04\times10^{-5}$) | $3.26\times10^{-3}$ ($5.07\times10^{-17}$) | $4.25\times10^{-3}$ ($3.40\times10^{-22}$) | 2:1875061- 2680063 | 0.13 (0.44) | $3.57\times10^{-4}$ ($4.62\times10^{-8}$) | $3.27\times10^{-4}$ ($4.41\times10^{-7}$) |
| **HT** | **3:76161795-77157986** | 0.94 ($8.74\times10^{-6}$) | $1.75\times10^{-3}$ ($1.17\times10^{-7}$) | $1.16\times10^{-3}$ ($5.42\times10^{-4}$) | **3:76157471- 77507832** | 0.67 (**$3.60\times10^{-3}$**) | $3.97\times10^{-4}$ ($8.18\times10^{-9}$) | $1.60\times10^{-4}$ ($6.62\times10^{-3}$) |
| **HT** | **7:38970303-40921737** | 0.98 ($1.74\times10^{-5}$) | $4.74\times10^{-4}$ (0.03) | $3.00\times10^{-3}$ ($1.15\times10^{-15}$) | **7:40261241-41415925** | 0.66 (**$1.62\times10^{-7}$**) | $5.54\times10^{-4}$ ($5.69\times10^{-15}$) | $5.51\times10^{-4}$ ($1.05\times10^{-14}$) |
| **HT** | **8:123448394-124868507** | 0.83 ($1.71\times10^{-5}$) | $1.54\times10^{-3}$ ($3.29\times10^{-6}$) | $1.67\times10^{-3}$ ($5.53\times10^{-6}$) | **8:124001020- 125453322** | 0.34 (**0.04**) | $2.45\times10^{-4}$ ($2.20\times10^{-4}$) | $6.16\times10^{-4}$ ($2.86\times10^{-15}$) |
| HT | 10:133514844-134600671 | 1.00 ($1.29\times10^{-6}$) | $8.33\times10^{-4}$ ($5.98\times10^{-3}$) | $1.72\times10^{-3}$ ($4.41\times10^{-6}$) | 10:133818037-134856054 | 0.26 (0.20) | $3.08\times10^{-4}$ ($9.20\times10^{-6}$) | $3.08\times10^{-4}$ ($9.75\times10^{-6}$) |
| HT | 12:110116509-112206126 | 0.68 ($8.97\times10^{-8}$) | 1.94 ($7.05\times10^{-10}$) | $3.50\times10^{-3}$ ($2.49\times10^{-21}$) | 12:110111515- 111592381 | 0.19 (0.34) | $1.64\times10^{-4}$ ($1.05\times10^{-3}$) | $4.12\times10^{-4}$ ($7.26\times10^{-12}$) |
| HT | 18:72211138-73568040 | 1.00 ($1.15\times10^{-6}$) | $9.27\times10^{-4}$ ($2.08\times10^{-3}$) | $1.52\times10^{-3}$ ($1.01\times10^{-5}$) | 18:72785207-73568260 | 0.24 (0.16) | $2.45\times10^{-4}$ ($7.20\times10^{-5}$) | $5.63\times10^{-4}$ ($5.58\times10^{-15}$) |
| HT | 19:40168770-41325021 | 1.00 ($1.08\times10^{-5}$) | $1.08\times10^{-3}$ ($7.40\times10^{-5}$) | $7.63\times10^{-4}$ ($3.00\times10^{-3}$) | 19:41004106-41733463 | -0.05 (0.87) | $1.14\times10^{-4}$ (0.02) | $2.50\times10^{-4}$ ($1.66\times10^{-5}$) |
| **HT** | **21:16262048-17832028** | 0.50 ($1.79\times10^{-5}$) | 2.94 ($1.17\times10^{-15}$) | 3.42 ($6.12\times10^{-17}$) | **21:16873444-18222037** | 0.37 (**$6.81\times10^{-3}$**) | $4.85\times10^{-4}$ ($9.60\times10^{-12}$) | $4.77\times10^{-4}$ ($2.71\times10^{-11}$) |
| T2D | 12:107850176-109020531 | 0.86 ($4.81\times10^{-6}$) | $1.84\times10^{-3}$ ($1.26\times10^{-8}$) | $1.09\times10^{-3}$ ($4.64\times10^{-5}$) | 12:108183189-109031819 | 0.28 (0.06) | $4.92\times10^{-4}$ ($1.25\times10^{-12}$) | $4.25\times10^{-4}$ ($5.73\times10^{-9}$) |
| **T2D** | **12:110116509-112206126** | 0.78 ($3.17\times10^{-9}$) | $1.94\times10^{-3}$ ($7.05\times10^{-10}$) | $2.74\times10^{-3}$ ($1.24\times10^{-18}$) | **12:110111515-111592381** | 0.75 (**$6.36\times10^{-3}$**) | $1.64\times10^{-4}$ ($1.05\times10^{-3}$) | $1.95\times10^{-4}$ ($1.02\times10^{-3}$) |
| **T2D** | **12:112206127-113925391** | 0.96 ($1.17\times10^{-5}$) | $8.04\times10^{-4}$ ($3.39\times10^{-3}$) | $1.88\times10^{-3}$ ($1.26\times10^{-9}$) | **12:111592382-113947983** | 0.39 (**0.02**) | $4.60\times10^{-4}$ ($1.11\times10^{-10}$) | $3.32\times10^{-4}$ ($6.42\times10^{-6}$) |
| T2D | 13:79597789-80800514 | 1.00 ($8.64\times10^{-6}$) | $1.22\times10^{-3}$ ($5.01\times10^{-5}$) | $7.46\times10^{-4}$ ($3.88\times10^{-3}$) | 13:79489046-80778339 | -0.16 (0.25) | $5.24\times10^{-4}$ ($5.07\times10^{-14}$) | $4.59\times10^{-4}$ ($1.30\times10^{-9}$) |
| T2D | 13:113803026-115109851 | 0.91 ($1.29\times10^{-6}$) | $1.39\times10^{-3}$ ($2.24\times10^{-5}$) | $1.49\times10^{-3}$ ($3.72\times10^{-8}$) | 13:113573119-114302561 | 0.27 (0.18) | $1.93\times10^{-4}$ ($8.07\times10^{-4}$) | $4.48\times10^{-4}$ ($3.25\times10^{-9}$) |

| | | | | | | | | |
|---|---|---|---|---|---|---|---|---|
| **T2D** | **16:52662976-54022423** | 0.74 ($1.97\times10^{-5}$) | $1.66\times10^{-3}$ ($2.91\times10^{-7}$) | $1.79\times10^{-3}$ ($1.86\times10^{-7}$) | **16:52041337-53393882** | 0.66 (**$1.15\times10^{-7}$**) | $5.97\times10^{-4}$ ($2.83\times10^{-15}$) | $6.45\times10^{-4}$ ($9.81\times10^{-14}$) |
| **T2D** | **16:65539267-67045298** | 1.00 ($1.21\times10^{-5}$) | 1.58 ($7.22\times10^{-7}$) | $6.73\times10^{-4}$ ($3.57\times10^{-3}$) | **16:65882821-66738843** | 0.58 (**$8.08\times10^{-4}$**) | $3.28\times10^{-4}$ ($5.78\times10^{-7}$) | $4.16\times10^{-4}$ ($4.22\times10^{-8}$) |
| **T2D** | **17:1388659-2627122** | 0.80 ($1.50\times10^{-5}$) | $2.72\times10^{-3}$ ($3.03\times10^{-12}$) | $8.88\times10^{-4}$ ($7.00\times10^{-4}$) | **17:1480057-2699135** | 0.60 (**$5.38\times10^{-5}$**) | $5.46\times10^{-4}$ ($7.84\times10^{-13}$) | $4.21\times10^{-4}$ ($8.11\times10^{-8}$) |
| **CVD** | **7:80834729-82021734** | 1.00 ($1.62\times10^{-5}$) | $6.97\times10^{-4}$ (0.01) | $9.68\times10^{-4}$ ($2.47\times10^{-6}$) | **7:80055722-81533566** | 0.67 (**$2.61\times10^{-3}$**) | $2.26\times10^{-4}$ ($5.62\times10^{-4}$) | $3.57\times10^{-4}$ ($7.49\times10^{-6}$) |
| CVD | 9:7373645-8011590 | -0.64 ($3.11\times10^{-5}$) | $1.56\times10^{-3}$ ($1.34\times10^{-6}$) | $1.75\times10^{-3}$ ($2.74\times10^{-13}$) | 9:7388203-8262303 | -0.11 (0.66) | $3.62\times10^{-4}$ ($1.53\times10^{-7}$) | $1.46\times10^{-4}$ (0.02) |
| **CVD** | **13:53609234-55588449** | 1.00 ($7.46\times10^{-6}$) | $7.70\times10^{-4}$ ($3.66\times10^{-3}$) | $9.05\times10^{-4}$ ($6.94\times10^{-5}$) | **13:54684857-55576912** | 0.64 (**0.03**) | $2.17\times10^{-4}$ ($5.39\times10^{-5}$) | $1.28\times10^{-4}$ (0.02) |
| CVD | 17:9751976-10575270 | 0.71 ($2.74\times10^{-5}$) | $2.87\times10^{-3}$ ($4.23\times10^{-16}$) | $9.61\times10^{-4}$ ($2.01\times10^{-4}$) | 17:9619357-10572617 | -0.27 (0.20) | $3.22\times10^{-4}$ ($1.77\times10^{-6}$) | $2.69\times10^{-4}$ ($2.84\times10^{-4}$) |
| **HP** | **3:67856978-69346534** | 1.00 ($3.30\times10^{-6}$) | $1.16\times10^{-3}$ ($5.51\times10^{-4}$) | $8.92\times10^{-4}$ ($2.16\times10^{-3}$) | **3:68319771-69784969** | -0.44 (**0.04**) | $1.93\times10^{-4}$ ($2.02\times10^{-3}$) | $4.03\times10^{-4}$ ($1.02\times10^{-7}$) |
| HP | 6:14325796-15374604 | 0.92 ($9.40\times10^{-6}$) | $1.02\times10^{-3}$ ($4.58\times10^{-4}$) | $1.85\times10^{-3}$ ($8.02\times10^{-8}$) | 6:13352327-15069421 | 0.28 (0.11) | $4.08\times10^{-4}$ ($4.75\times10^{-8}$) | $4.16\times10^{-4}$ ($7.27\times10^{-7}$) |
| **HP** | **8:1957826-2571479** | 0.58 ($1.13\times10^{-4}$) | $1.82\times10^{-3}$ ($1.57\times10^{-9}$) | $2.05\times10^{-3}$ ($3.37\times10^{-10}$) | **8:2396123-2970491** | 0.56 (**0.04**) | $1.65\times10^{-4}$ ($4.57\times10^{-3}$) | $2.59\times10^{-4}$ ($3.42\times10^{-4}$) |
| HP | 10:12988489-13717645 | 1.00 ($6.01\times10^{-8}$) | $1.69\times10^{-3}$ ($1.15\times10^{-7}$) | $1.19\times10^{-3}$ ($2.18\times10^{-4}$) | 10:13208062-14008135 | -0.03 (0.90) | $2.27\times10^{-4}$ ($4.47\times10^{-4}$) | $4.09\times10^{-4}$ ($5.31\times10^{-7}$) |
| HP | 11:114864027-116311324 | 0.76 ($1.31\times10^{-6}$) | 1.47 ($7.07\times10^{-6}$) | $2.47\times10^{-3}$ ($4.48\times10^{-14}$) | 11:114742318-116247377 | 0.13 (0.41) | 4.67 ($6.85\times10^{-10}$) | $4.18\times10^{-4}$ ($1.36\times10^{-6}$) |
| HP | 12:95717323-96555768 | -0.75 ($2.45\times10^{-5}$) | 1.65 ($1.15\times10^{-7}$) | $1.39\times10^{-3}$ ($1.38\times10^{-6}$) | 12:96017783-96828314 | 0.05 (0.87) | $2.84\times10^{-4}$ ($5.23\times10^{-6}$) | $1.55\times10^{-4}$ (0.02) |
| HP | 15:92881034-93723088 | 0.81 ($2.55\times10^{-6}$) | $1.63\times10^{-3}$ ($1.37\times10^{-6}$) | $1.85\times10^{-3}$ ($1.86\times10^{-8}$) | 15:92675406-93707010 | 0.17 (0.28) | $4.30\times10^{-4}$ ($4.17\times10^{-9}$) | $5.00\times10^{-4}$ ($6.05\times10^{-9}$) |
| FL | 6:2898448-4319834 | 0.67 ($1.80\times10^{-6}$) | $1.35\times10^{-3}$ ($2.35\times10^{-5}$) | $4.01\times10^{-3}$ ($7.95\times10^{-30}$) | - | - | - | - |
| FL | 6:155739846-157043139 | -1.00 ($1.10\times10^{-10}$) | $1.96\times10^{-3}$ ($7.57\times10^{-8}$) | $9.42\times10^{-4}$ ($7.84\times10^{-6}$) | - | - | - | - |
| FL | 15:40554834-42332112 | 1.00 ($7.61\times10^{-6}$) | $6.81\times10^{-4}$ (0.01) | $1.28\times10^{-3}$ ($7.55\times10^{-6}$) | - | - | - | - |

Note: HT, hypertension; T2D, type 2 diabetes; CVD, cardiovascular disease; HL, hyperlipidemia; FL, fatty liver.

**Table S8.** Significant genes identified through gene-based analyses of obesity-related indicators and obesity-related diseases

(A) Significant genes identified from gene-based analyses of obesity-related indicators

| CHR | GENE | P for general obesity by BMI | | | | P for abdominal obesity by WC | | | |
|---|---|---|---|---|---|---|---|---|---|
| | | Discovery | REP1$_{Kor}$ | REP2$_{Chi}$ | REP3$_{NHW}$ | Discovery | REP1$_{Kor}$ | REP2$_{Chi}$ | REP3$_{NHW}$ |
| 1 | *HIVEP3* | $2.37\times10^{-6}$ | 0.36 | 0.30 | **$2.15\times10^{-3}$** | | - | - | - |
| | *AGBL4* | $1.31\times10^{-6}$ | 0.49 | 0.08 | **$4.57\times10^{-4}$** | | - | - | - |
| 2 | *ADCY3* | $4.96\times10^{-13}$ | **$4.49\times10^{-4}$** | 0.40 | **$4.21\times10^{-14}$** | | - | - | - |
| | *EFR3B* | $2.98\times10^{-8}$ | 0.11 | 0.44 | **$2.64\times10^{-10}$** | | - | - | - |
| | *ACYP2* | $2.48\times10^{-8}$ | 0.43 | 0.08 | **$2.11\times10^{-4}$** | | - | - | - |
| 3 | *STAB1* | $2.99\times10^{-7}$ | 0.16 | 0.20 | **$2.46\times10^{-4}$** | | - | - | - |
| | *NT5DC2* | $3.61\times10^{-8}$ | 0.13 | 0.17 | **$3.25\times10^{-7}$** | | - | - | - |
| | *SMIM4* | $2.03\times10^{-8}$ | 0.13 | 0.33 | **$1.94\times10^{-6}$** | | - | - | - |
| | *PBRM1* | $2.00\times10^{-8}$ | 0.13 | 0.36 | **$1.51\times10^{-6}$** | | - | - | - |
| | *GNL3* | $2.59\times10^{-8}$ | 0.15 | 0.34 | **$2.93\times10^{-7}$** | | - | - | - |
| | *GLT8D1* | $2.30\times10^{-8}$ | 0.17 | 0.39 | **$2.01\times10^{-6}$** | | - | - | - |
| | *SPCS1* | $2.64\times10^{-8}$ | 0.15 | 0.36 | **$1.08\times10^{-5}$** | | - | - | - |
| | *NEK4* | $1.80\times10^{-8}$ | 0.16 | 0.33 | **$3.01\times10^{-6}$** | | - | - | - |
| | *ITIH1* | $7.56\times10^{-8}$ | 0.16 | 0.80 | **$1.17\times10^{-7}$** | | - | - | - |
| | *MUSTN1* | $1.91\times10^{-6}$ | 0.19 | 0.62 | **$5.51\times10^{-6}$** | | - | - | - |
| | *TMEM110-MUSTN1* | $5.24\times10^{-7}$ | 0.15 | 0.78 | **$1.39\times10^{-11}$** | | - | - | - |
| | *TMEM110* | $5.61\times10^{-7}$ | 0.15 | 0.78 | **$1.62\times10^{-11}$** | | - | - | - |
| | *SFMBT1* | $1.28\times10^{-7}$ | 0.46 | 0.96 | **$7.71\times10^{-10}$** | | - | - | - |
| 6 | *CDKAL1* | $2.19\times10^{-8}$ | **0.02** | 0.76 | **$1.01\times10^{-3}$** | | - | - | - |
| | *C6orf1* | - | - | - | - | $5.11\times10^{-8}$ | 1.00 | 0.12 | **$5.48\times10^{-11}$** |
| 7 | *AUTS2* | $9.88\times10^{-7}$ | 0.60 | 0.06 | **$3.30\times10^{-7}$** | | - | - | - |
| 10 | *INPP5A* | $2.64\times10^{-6}$ | 0.32 | 0.38 | **$2.66\times10^{-3}$** | | - | - | - |
| 11 | *BDNF* | $2.50\times10^{-12}$ | **0.04** | 0.41 | **$5.00\times10^{-10}$** | | - | - | - |
| 18 | *GAREM* | $1.31\times10^{-7}$ | 0.90 | 0.03 | **0.05** | | - | - | - |
| 19 | *GIPR* | $1.24\times10^{-10}$ | - | 0.28 | **$6.11\times10^{-16}$** | | - | - | - |
| | *SNRPD2* | $1.95\times10^{-6}$ | - | 0.23 | **$4.22\times10^{-10}$** | | - | - | - |
| | *QPCTL* | $1.26\times10^{-8}$ | - | 0.32 | **$4.37\times10^{-10}$** | | - | - | - |
| | *FBXO46* | $4.79\times10^{-8}$ | - | 0.49 | **$3.47\times10^{-5}$** | $1.02\times10^{-6}$ | - | 0.69 | **$3.41\times10^{-4}$** |
| | *ZC3H4* | $3.32\times10^{-7}$ | 0.24 | 0.70 | **$2.02\times10^{-11}$** | | - | - | - |
| | *U2AF2* | - | - | - | - | $1.65\times10^{-6}$ | 0.82 | 0.26 | **0.03** |

(B) Significant genes identified from gene-based analyses of obesity-related diseases

| CHR | GENE | P for obesity-related diseases in discovery/$REP3_{NHW}$ | | | | | | | |
|---|---|---|---|---|---|---|---|---|---|
| | | HT | | T2D | | HL | | FL | |
| | | Discovery | $REP3_{NHW}$ | Discovery | $REP3_{NHW}$ | Discovery | $REP3_{NHW}$ | Discovery | $REP3_{NHW}$ |
| 1 | *TCEA3* | $1.12 \times 10^{-6}$ | 0.10 | - | - | - | - | - | - |
| | *CELSR2* | - | - | - | - | $7.91 \times 10^{-10}$ | **$5.00 \times 10^{-10}$** | - | - |
| | *WNT2B* | $2.89 \times 10^{-8}$ | **$1.28 \times 10^{-15}$** | - | - | - | - | - | - |
| 2 | *TDRD15* | - | - | - | - | $2.33 \times 10^{-6}$ | **$3.01 \times 10^{-11}$** | - | - |
| | *KCNK3* | $1.55 \times 10^{-11}$ | **$4.16 \times 10^{-15}$** | - | - | - | - | - | - |
| | *AC109829.1* | - | - | - | - | $1.67 \times 10^{-6}$ | **$1.01 \times 10^{-12}$** | - | - |
| | ***SPTBN1*** | $3.52 \times 10^{-8}$ | **$4.12 \times 10^{-3}$** | - | - | - | - | - | - |
| | *EML6* | $3.19 \times 10^{-9}$ | **$3.04 \times 10^{-3}$** | - | - | - | - | - | - |
| 3 | *FGD5* | $8.05 \times 10^{-7}$ | **$1.10 \times 10^{-9}$** | - | - | - | - | - | - |
| | *UBE2E2* | - | - | $2.96 \times 10^{-7}$ | **$7.86 \times 10^{-10}$** | - | - | - | - |
| | *CACNA1D* | $3.04 \times 10^{-9}$ | **$1.37 \times 10^{-8}$** | - | - | - | - | - | - |
| | ***LRRIQ4*** | $4.23 \times 10^{-7}$ | **$5.10 \times 10^{-3}$** | - | - | - | - | - | - |
| | *IGF2BP2* | - | - | $3.27 \times 10^{-7}$ | **$5.00 \times 10^{-10}$** | - | - | - | - |
| 4 | *CRIPAK* | - | - | $1.06 \times 10^{-6}$ | 0.36 | - | - | - | - |
| 5 | *HMGCR* | - | - | - | - | $1.48 \times 10^{-7}$ | **$1.01 \times 10^{-14}$** | - | - |
| | *PRDM6* | $2.61 \times 10^{-10}$ | **$6.61 \times 10^{-4}$** | - | - | - | - | - | - |
| 6 | *CDKAL1* | - | - | $3.19 \times 10^{-18}$ | **$8.09 \times 10^{-14}$** | - | - | - | - |
| 7 | *HOXA13* | $1.96 \times 10^{-6}$ | 0.36 | - | - | - | - | - | - |
| 8 | *GML* | $1.43 \times 10^{-9}$ | **$2.53 \times 10^{-5}$** | - | - | - | - | - | - |
| | *CYP11B2* | $1.04 \times 10^{-13}$ | **$2.81 \times 10^{-4}$** | - | - | - | - | - | - |
| 9 | ***STXBP1*** | $1.72 \times 10^{-7}$ | **$5.50 \times 10^{-3}$** | - | - | - | - | - | - |
| | ***PTRH1*** | $4.27 \times 10^{-7}$ | **$2.32 \times 10^{-3}$** | - | - | - | - | - | - |
| | *C9orf117* | $9.30 \times 10^{-7}$ | **0.02** | - | - | - | - | - | - |
| | ***TTC16*** | $4.95 \times 10^{-7}$ | **$4.21 \times 10^{-3}$** | - | - | - | - | - | - |
| 10 | *CDC123* | - | - | $1.52 \times 10^{-6}$ | **$7.82 \times 10^{-10}$** | - | - | - | - |
| | *IDE* | - | - | $3.74 \times 10^{-9}$ | **$4.46 \times 10^{-6}$** | - | - | - | - |
| | *TMEM180* | $2.67 \times 10^{-9}$ | 0.26 | - | - | - | - | - | - |
| | *SUFU* | $7.94 \times 10^{-10}$ | 0.20 | - | - | - | - | - | - |
| | *TRIM8* | $6.11 \times 10^{-7}$ | 0.14 | - | - | - | - | - | - |
| | ***ARL3*** | $2.53 \times 10^{-11}$ | **$5.65 \times 10^{-4}$** | - | - | - | - | - | - |
| | ***SFXN2*** | $1.68 \times 10^{-6}$ | **$1.63 \times 10^{-3}$** | - | - | - | - | - | - |
| | *WBP1L* | $1.02 \times 10^{-7}$ | **$6.68 \times 10^{-3}$** | - | - | - | - | - | - |

| | | | | | | | | | |
|---|---|---|---|---|---|---|---|---|---|
| | *CYP17A1* | $2.81 \times 10^{-10}$ | **$1.34 \times 10^{-3}$** | - | - | - | - | - | - |
| | *C10orf32-ASMT* | $4.57 \times 10^{-12}$ | **$2.74 \times 10^{-5}$** | - | - | - | - | - | - |
| | *AS3MT* | $5.33 \times 10^{-12}$ | **$2.46 \times 10^{-5}$** | - | - | - | - | - | - |
| | *CNNM2* | $1.84 \times 10^{-10}$ | **$5.09 \times 10^{-4}$** | - | - | - | - | - | - |
| | *NT5C2* | $5.10 \times 10^{-9}$ | **$3.90 \times 10^{-4}$** | - | - | - | - | - | - |
| | ***INA*** | $1.44 \times 10^{-6}$ | **$7.88 \times 10^{-9}$** | - | - | - | - | - | - |
| | ***PDCD11*** | $5.49 \times 10^{-8}$ | **$2.95 \times 10^{-4}$** | - | - | - | - | - | - |
| | *CALHM1* | $3.99 \times 10^{-7}$ | 0.30 | - | - | - | - | - | - |
| 11 | *KCNJ11* | - | - | $1.14 \times 10^{-6}$ | **$2.46 \times 10^{-8}$** | - | - | - | - |
| | *LRRC10B* | $1.49 \times 10^{-6}$ | **0.02** | - | - | - | - | - | - |
| | *BUD13* | - | - | - | - | $1.46 \times 10^{-9}$ | **$2.33 \times 10^{-13}$** | - | - |
| | *ZNF259* | - | - | - | - | $5.64 \times 10^{-12}$ | **$1.50 \times 10^{-14}$** | - | - |
| | ***APOA4*** | - | - | - | - | $3.65 \times 10^{-7}$ | **$6.16 \times 10^{-7}$** | - | - |
| | *APOA1* | - | - | - | - | $4.50 \times 10^{-10}$ | 0.18 | - | - |
| | *SIK3* | - | - | - | - | $3.07 \times 10^{-9}$ | **0.03** | - | - |
| | *PAFAH1B2* | - | - | - | - | $1.36 \times 10^{-7}$ | **0.01** | - | - |
| | *PCSK7* | - | - | - | - | $2.16 \times 10^{-6}$ | **$4.60 \times 10^{-3}$** | - | - |
| 12 | *ATP2B1* | 5.00E-10 | **$5.00 \times 10^{-10}$** | - | - | - | - | - | - |
| | *C12orf42* | - | - | $1.51 \times 10^{-7}$ | **0.02** | - | - | - | - |
| 13 | *SLC7A1* | 2.93E-09 | **$4.11 \times 10^{-5}$** | - | - | - | - | - | - |
| 15 | *FURIN* | 8.60E-07 | **$9.35 \times 10^{-6}$** | - | - | - | - | - | - |
| 16 | ***TXNL4B*** | - | - | - | - | $2.52 \times 10^{-7}$ | **$1.13 \times 10^{-6}$** | - | - |
| | ***HP*** | - | - | - | - | $1.14 \times 10^{-7}$ | **$4.57 \times 10^{-6}$** | - | - |
| | *HPR* | - | - | - | - | $4.95 \times 10^{-8}$ | **$7.77 \times 10^{-6}$** | - | - |
| | *DHX38* | - | - | - | - | $2.25 \times 10^{-6}$ | **$4.63 \times 10^{-7}$** | - | - |
| | *PMFBP1* | - | - | - | - | $1.39 \times 10^{-7}$ | **$8.75 \times 10^{-7}$** | - | - |
| 18 | *LIPG* | - | - | - | - | $1.57 \times 10^{-7}$ | **$3.58 \times 10^{-4}$** | - | - |
| 19 | *DOCK6* | - | - | - | - | $4.21 \times 10^{-7}$ | **$2.47 \times 10^{-3}$** | - | - |
| | *TOMM40* | - | - | - | - | $2.39 \times 10^{-12}$ | **$5.00 \times 10^{-10}$** | - | - |
| | ***APOE*** | - | - | - | - | $1.25 \times 10^{-10}$ | **$5.00 \times 10^{-10}$** | - | - |
| 20 | *GDAP1L1* | - | - | $8.94 \times 10^{-8}$ | **$1.31 \times 10^{-3}$** | - | - | - | - |
| | *FITM2* | - | - | $4.68 \times 10^{-9}$ | **$1.04 \times 10^{-4}$** | - | - | - | - |
| | *R3HDML* | - | - | $1.00 \times 10^{-9}$ | **0.01** | - | - | - | - |
| | *HNF4A* | - | - | $3.51 \times 10^{-8}$ | **$2.37 \times 10^{-4}$** | - | - | - | - |
| 22 | *PNPLA3* | - | - | - | - | - | - | $5.00 \times 10^{-10}$ | - |

Note: HT, hypertension; T2D, type 2 diabetes; CVD, cardiovascular disease; HL, hyperlipidemia; FL, fatty liver.

**Table S9.** Summary for gene-set analyses results for obesity-related diseases

| Disease | Method: Database | Gene-set 1: obesity | | | Gene-set 2: obesity-related diseases | | |
|---|---|---|---|---|---|---|---|
| | | **Pathway counts (Significant / Total)** | **Minimum P value** | **Minimum FDR** | **Pathway counts (Significant / Total)** | **Minimum P value** | **Minimum FDR** |
| HT | GSEA: Reactome | 0 / 139 | 0.06 | 0.46 | 0 / 95 | 0.06 | 0.44 |
| | ORA1: GO | 0 / 124 | $1.53 \times 10^{-3}$ | 0.23 | 0 / 0 | - | - |
| | ORA2: KEGG | 0 / 0 | - | - | **3 / 5** | $1.49 \times 10^{-5}$ | **$9.56 \times 10^{-4}$** |
| T2D | GSEA: Reactome | 0 / 143 | 0.08 | 0.43 | 0 / 10 | 0.65 | 1.00 |
| | ORA1: GO | 0 / 149 | $9.57 \times 10^{-4}$ | 0.17 | 0 / 128 | $2.63 \times 10^{-3}$ | 0.06 |
| | ORA2: KEGG | 0 / 0 | - | - | 0 / 2 | 0.03 | 0.07- |
| HP | GSEA: Reactome | 0/ 144 | 0.15 | 0.43- | 0 / 41 | 0.35 | 0.98- |
| | ORA1: GO | 0 / 110 | $2.63 \times 10^{-3}$ | 0.22- | **172 / 572** | $5.97 \times 10^{-10}$ | **$3.41 \times 10^{-7}$** |
| | ORA2: KEGG | 0 / 0 | - | - | **4 / 13** | $1.39 \times 10^{-7}$ | **$2.22 \times 10^{-6}$** |
| FL | GSEA: Reactome | 0 / 139 | 0.22 | 0.45 | 0 / 41 | 0.35 | 0.98 |
| | ORA1: GO | 0 / 124 | $1.53 \times 10^{-3}$ | 0.23 | **50 / 50** | $1.47 \times 10^{-3}$ | **0.02** |
| | ORA2: KEGG | 0 / 0 | **-** | **-** | **2 / 2** | 0.02 | **0.02** |

Note: HT, hypertension; T2D, type 2 diabetes; HL, hyperlipidemia; FL, fatty liver.

**Table S10.** Significant gene-sets identified through gene-set analyses of obesity-related diseases

(A) Significant gene-sets associated with HT from of ORA2: KEGG

| ID | Category: Subcategory: Description | Gene-set | GeneRatio | P value | FDR |
|---|---|---|---|---|---|
| hsa04925 | Organismal Systems: Endocrine system: Aldosterone synthesis and secretion | *CACNA1D, CYP11B2, KCNK3, ATP2B1* | 4/14 | $1.49\times10^{-5}$ | $9.56\times 10^{-4}$ |
| hsa04934 | Human Diseases: Endocrine and metabolic disease: Cushing syndrome | *CYP17A1, CACNA1D, WNT2B, KCNK3* | 4/14 | $9.06\times 10^{-5}$ | $2.90\times 10^{-3}$ |
| hsa04927 | Organismal Systems: Endocrine system: Cortisol synthesis and secretion | *CYP17A1, CACNA1D, KCNK3* | 4/14 | $1.44\times 10^{-4}$ | $3.08\times 10^{-3}$ |

(B) Significant gene-sets associated with HL from ORA1: GO

| ID | Description | Gene-set | GeneRatio | P value | FDR |
|---|---|---|---|---|---|
| GO:0034375 | high-density lipoprotein particle remodeling | *LIPG, APOE, APOA4, APOA1* | 4/15 | $5.97\times10^{-10}$ | $3.41\times10^{-7}$ |
| GO:0043691 | reverse cholesterol transport | *LIPG, APOE, APOA4, APOA1* | 4/15 | $1.21\times10^{-9}$ | $3.47\times10^{-7}$ |
| GO:0034368 | protein-lipid complex remodeling | *LIPG, APOE, APOA4, APOA1* | 4/15 | $1.02\times10^{-8}$ | $1.46\times10^{-6}$ |
| GO:0034369 | plasma lipoprotein particle remodeling | *LIPG, APOE, APOA4, APOA1* | 4/15 | $1.02\times10^{-8}$ | $1.46\times10^{-6}$ |
| GO:0034367 | protein-containing complex remodeling | *LIPG, APOE, APOA4, APOA1* | 4/15 | $1.30\times10^{-8}$ | $1.49\times10^{-6}$ |
| GO:1905918 | regulation of CoA-transferase activity | *APOE, APOA4, APOA1* | 3/15 | $4.77\times10^{-8}$ | $4.55\times10^{-6}$ |
| GO:0071827 | plasma lipoprotein particle organization | *LIPG, APOE, APOA4, APOA1* | 4/15 | $5.69\times10^{-8}$ | $4.65\times10^{-6}$ |
| GO:0071825 | protein-lipid complex organization | *LIPG, APOE, APOA4, APOA1* | 4/15 | $7.22\times10^{-8}$ | $5.00\times10^{-6}$ |
| GO:0034370 | triglyceride-rich lipoprotein particle remodeling | *APOE, APOA4, APOA1* | 3/15 | $8.74\times10^{-8}$ | $5.00\times10^{-6}$ |
| GO:0034372 | very-low-density lipoprotein particle remodeling | *APOE, APOA4, APOA1* | 3/15 | $8.74\times10^{-8}$ | $5.00\times10^{-6}$ |
| GO:0033700 | phospholipid efflux | *APOE, APOA4, APOA1* | 3/15 | $1.44\times10^{-7}$ | $7.51\times10^{-6}$ |
| GO:0034384 | high-density lipoprotein particle clearance | *LIPG, APOE, APOA1* | 3/15 | $3.23\times10^{-7}$ | $1.35\times10^{-5}$ |
| GO:0032371 | regulation of sterol transport | *LIPG, APOE, APOA4, APOA1* | 4/15 | $3.30\times10^{-7}$ | $1.35\times10^{-5}$ |
| GO:0032374 | regulation of cholesterol transport | *LIPG, APOE, APOA4, APOA1* | 4/15 | $3.30\times10^{-7}$ | $1.35\times10^{-5}$ |
| GO:0097006 | regulation of plasma lipoprotein particle levels | *LIPG, APOE, APOA4, APOA1* | 4/15 | $6.47\times10^{-7}$ | $2.47\times10^{-5}$ |
| GO:0042157 | lipoprotein metabolic process | *LIPG, APOE, APOA4, APOA1* | 4/15 | $1.02\times10^{-6}$ | $3.66\times10^{-5}$ |
| GO:0034377 | plasma lipoprotein particle assembly | *APOE, APOA4, APOA1* | 3/15 | $1.15\times10^{-6}$ | $3.74\times10^{-5}$ |
| GO:0042632 | cholesterol homeostasis | *LIPG, APOE, APOA4, APOA1* | 4/15 | $1.19\times10^{-6}$ | $3.74\times10^{-5}$ |
| GO:0055092 | sterol homeostasis | *LIPG, APOE, APOA4, APOA1* | 4/15 | $1.24\times10^{-6}$ | $3.74\times10^{-5}$ |

| | | | | | |
|---|---|---|---|---|---|
| GO:0065005 | protein-lipid complex assembly | *APOE, APOA4, APOA1* | 3/15 | $1.60\times10^{-6}$ | $4.57\times10^{-5}$ |
| GO:0030301 | cholesterol transport | *LIPG, APOE, APOA4, APOA1* | 4/15 | $2.17\times10^{-6}$ | $5.90\times10^{-5}$ |
| GO:0032373 | positive regulation of sterol transport | *LIPG, APOE, APOA1* | 3/15 | $2.80\times10^{-6}$ | $6.84\times10^{-5}$ |
| GO:0032376 | positive regulation of cholesterol transport | *LIPG, APOE, APOA1* | 3/15 | $2.80\times10^{-6}$ | $6.84\times10^{-5}$ |
| GO:0015918 | sterol transport | *LIPG, APOE, APOA4, APOA1* | 4/15 | $2.87\times10^{-6}$ | $6.84\times10^{-5}$ |
| GO:0008203 | cholesterol metabolic process | *APOE, APOA4, HMGCR, APOA1* | 4/15 | $4.53\times10^{-6}$ | $1.04\times10^{-4}$ |
| GO:0055090 | acylglycerol homeostasis | *APOE, APOA4, APOA1* | 3/15 | $5.55\times10^{-6}$ | $1.18\times10^{-4}$ |
| GO:0032368 | regulation of lipid transport | *LIPG, APOE, APOA4, APOA1* | 4/15 | $5.58\times10^{-6}$ | $1.18\times10^{-4}$ |
| GO:0016125 | sterol metabolic process | *APOE, APOA4, HMGCR, APOA1* | 4/15 | $6.02\times10^{-6}$ | $1.19\times10^{-4}$ |
| GO:1902652 | secondary alcohol metabolic process | *APOE, APOA4, HMGCR, APOA1* | 4/15 | $6.02\times10^{-6}$ | $1.19\times10^{-4}$ |
| GO:0055088 | lipid homeostasis | *LIPG, APOE, APOA4, APOA1* | 4/15 | $9.83\times10^{-6}$ | $1.87\times10^{-4}$ |
| GO:0034381 | plasma lipoprotein particle clearance | *LIPG, APOE, APOA1* | 3/15 | $1.02\times10^{-5}$ | $1.88\times10^{-4}$ |
| GO:1905952 | regulation of lipid localization | *LIPG, APOE, APOA4, APOA1* | 4/15 | $1.17\times10^{-5}$ | $2.09\times10^{-4}$ |
| GO:0006695 | cholesterol biosynthetic process | *APOE, HMGCR, APOA1* | 3/15 | $1.69\times10^{-5}$ | $2.85\times10^{-4}$ |
| GO:1902653 | secondary alcohol biosynthetic process | *APOE, HMGCR, APOA1* | 3/15 | $1.69\times10^{-5}$ | $2.85\times10^{-4}$ |
| GO:0016126 | sterol biosynthetic process | *APOE, HMGCR, APOA1* | 3/15 | $2.30\times10^{-5}$ | $3.76\times10^{-4}$ |
| GO:0033344 | cholesterol efflux | *APOE, APOA4, APOA1* | 3/15 | $2.50\times10^{-5}$ | $3.97\times10^{-4}$ |
| GO:0035023 | regulation of Rho protein signal transduction | *APOE, DOCK6, APOA1* | 3/15 | $3.66\times10^{-5}$ | $5.66\times10^{-4}$ |
| GO:1904729 | regulation of intestinal lipid absorption | *APOA4, APOA1* | 2/15 | $3.83\times10^{-5}$ | $5.76\times10^{-4}$ |
| GO:0032370 | positive regulation of lipid transport | *LIPG, APOE, APOA1* | 3/15 | $4.21\times10^{-5}$ | $6.17\times10^{-4}$ |
| GO:0034380 | high-density lipoprotein particle assembly | *APOE, APOA1* | 2/15 | $4.52\times10^{-5}$ | $6.47\times10^{-4}$ |
| GO:1904478 | regulation of intestinal absorption | *APOA4, APOA1* | 2/15 | $5.27\times10^{-5}$ | $7.36\times10^{-4}$ |
| GO:0098869 | cellular oxidant detoxification | *APOE, HP, APOA4* | 3/15 | $5.98\times10^{-5}$ | $8.09\times10^{-4}$ |
| GO:2001140 | positive regulation of phospholipid transport | *APOE, APOA1* | 2/15 | $6.08\times10^{-5}$ | $8.09\times10^{-4}$ |
| GO:0015850 | organic hydroxy compound transport | *LIPG, APOE, APOA4, APOA1* | 4/15 | $6.54\times10^{-5}$ | $8.51\times10^{-4}$ |
| GO:2001138 | regulation of phospholipid transport | *APOE, APOA1* | 2/15 | $6.95\times10^{-5}$ | $8.83\times10^{-4}$ |
| GO:0015914 | phospholipid transport | *APOE, APOA4, APOA1* | 3/15 | $7.13\times10^{-5}$ | $8.86\times10^{-4}$ |
| GO:0006641 | triglyceride metabolic process | *LIPG, APOE, APOA4* | 3/15 | $7.75\times10^{-5}$ | $9.19\times10^{-4}$ |
| GO:0032488 | Cdc42 protein signal transduction | *APOE, APOA1* | 2/15 | $7.87\times10^{-5}$ | $9.19\times10^{-4}$ |
| GO:0055091 | phospholipid homeostasis | *LIPG, APOA1* | 2/15 | $7.87\times10^{-5}$ | $9.19\times10^{-4}$ |
| GO:0030299 | intestinal cholesterol absorption | *APOA4, APOA1* | 2/15 | $8.85\times10^{-5}$ | $9.94\times10^{-4}$ |
| GO:1905954 | positive regulation of lipid localization | *LIPG, APOE, APOA1* | 3/15 | $8.87\times10^{-5}$ | $9.94\times10^{-4}$ |
| GO:0090205 | positive regulation of cholesterol metabolic process | *APOE, APOA1* | 2/15 | $9.89\times10^{-5}$ | $1.09\times10^{-3}$ |
| GO:1900221 | regulation of amyloid-beta clearance | *APOE, HMGCR* | 2/15 | $1.10\times10^{-4}$ | $1.17\times10^{-3}$ |

| | | | | | |
|---|---|---|---|---|---|
| GO:0008202 | steroid metabolic process | *APOE, APOA4, HMGCR, APOA1* | 4/15 | $1.10\times10^{-4}$ | $1.17\times10^{-3}$ |
| GO:1990748 | cellular detoxification | *APOE, HP, APOA4* | 3/15 | $1.20\times10^{-4}$ | $1.24\times10^{-3}$ |
| GO:0044241 | lipid digestion | *APOA4, APOA1* | 2/15 | $1.33\times10^{-4}$ | $1.34\times10^{-3}$ |
| GO:0016042 | lipid catabolic process | *LIPG, APOE, APOA4, PAFAH1B2* | 4/15 | $1.33\times10^{-4}$ | $1.34\times10^{-3}$ |
| GO:0045834 | positive regulation of lipid metabolic process | *APOE, APOA4, APOA1* | 3/15 | $1.41\times10^{-4}$ | $1.39\times10^{-3}$ |
| GO:0006639 | acylglycerol metabolic process | *LIPG, APOE, APOA4* | 3/15 | $1.50\times10^{-4}$ | $1.42\times10^{-3}$ |
| GO:0097237 | cellular response to toxic substance | *APOE, HP, APOA4* | 3/15 | $1.50\times10^{-4}$ | $1.42\times10^{-3}$ |
| GO:0006066 | alcohol metabolic process | *APOE, APOA4, HMGCR, APOA1* | 4/15 | $1.52\times10^{-4}$ | $1.42\times10^{-3}$ |
| GO:0006638 | neutral lipid metabolic process | *LIPG, APOE, APOA4* | 3/15 | $1.54\times10^{-4}$ | $1.42\times10^{-3}$ |
| GO:0098856 | intestinal lipid absorption | *APOA4, APOA1* | 2/15 | $1.59\times10^{-4}$ | $1.45\times10^{-3}$ |
| GO:0007266 | Rho protein signal transduction | *APOE, DOCK6, APOA1* | 3/15 | $1.64\times10^{-4}$ | $1.46\times10^{-3}$ |
| GO:0046486 | glycerolipid metabolic process | *LIPG, APOE, APOA4, APOA1* | 4/15 | $1.90\times10^{-4}$ | $1.68\times10^{-3}$ |
| GO:0046165 | alcohol biosynthetic process | *APOE, HMGCR, APOA1* | 3/15 | $1.97\times10^{-4}$ | $1.71\times10^{-3}$ |
| GO:0010875 | positive regulation of cholesterol efflux | *APOE, APOA1* | 2/15 | $2.02\times10^{-4}$ | $1.73\times10^{-3}$ |
| GO:0062013 | positive regulation of small molecule metabolic process | *APOE, APOA4, APOA1* | 3/15 | $2.17\times10^{-4}$ | $1.80\times10^{-3}$ |
| GO:0019433 | triglyceride catabolic process | *LIPG, APOA4* | 2/15 | $2.18\times10^{-4}$ | $1.80\times10^{-3}$ |
| GO:0042744 | hydrogen peroxide catabolic process | *HP, APOA4* | 2/15 | $2.67\times10^{-4}$ | $2.19\times10^{-3}$ |
| GO:0098754 | detoxification | *APOE, HP, APOA4* | 3/15 | $2.81\times10^{-4}$ | $2.27\times10^{-3}$ |
| GO:0045940 | positive regulation of steroid metabolic process | *APOE, APOA1* | 2/15 | $2.85\times10^{-4}$ | $2.27\times10^{-3}$ |
| GO:0015748 | organophosphate ester transport | *APOE, APOA4, APOA1* | 3/15 | $3.12\times10^{-4}$ | $2.45\times10^{-3}$ |
| GO:0006869 | lipid transport | *LIPG, APOE, APOA4, APOA1* | 4/15 | $3.72\times10^{-4}$ | $2.88\times10^{-3}$ |
| GO:0031329 | regulation of cellular catabolic process | *APOE, HP, APOA4, PAFAH1B2* | 4/15 | $4.14\times10^{-4}$ | $3.11\times10^{-3}$ |
| GO:0006694 | steroid biosynthetic process | *APOE, MGCR, APOA1* | 3/15 | $4.17\times10^{-4}$ | $3.11\times10^{-3}$ |
| GO:0046461 | neutral lipid catabolic process | *LIPG, APOA4* | 2/15 | $4.24\times10^{-4}$ | $3.11\times10^{-3}$ |
| GO:0046464 | acylglycerol catabolic process | *LIPG, APOA4* | 2/15 | $4.24\times10^{-4}$ | $3.11\times10^{-3}$ |
| GO:0097242 | amyloid-beta clearance | *APOE, HMGCR* | 2/15 | $4.69\times10^{-4}$ | $3.40\times10^{-3}$ |
| GO:0000302 | response to reactive oxygen species | *APOE, HP, APOA4* | 3/15 | $4.77\times10^{-4}$ | $3.41\times10^{-3}$ |
| GO:0044058 | regulation of digestive system process | *APOA4, APOA1* | 2/15 | $5.16\times10^{-4}$ | $3.65\times10^{-3}$ |
| GO:0050892 | intestinal absorption | *APOA4, APOA1* | 2/15 | $5.41\times10^{-4}$ | $3.73\times10^{-3}$ |
| GO:0070328 | triglyceride homeostasis | *APOE, APOA1* | 2/15 | $5.41\times10^{-4}$ | $3.73\times10^{-3}$ |
| GO:0071402 | cellular response to lipoprotein particle stimulus | *APOE, APOA1* | 2/15 | $5.66\times10^{-4}$ | $3.85\times10^{-3}$ |
| GO:0090181 | regulation of cholesterol metabolic process | *APOE, APOA1* | 2/15 | $5.91\times10^{-4}$ | $3.93\times10^{-3}$ |
| GO:0090207 | regulation of triglyceride metabolic process | *APOE, APOA4* | 2/15 | $5.91\times10^{-4}$ | $3.93\times10^{-3}$ |

| | | | | | |
|---|---|---|---|---|---|
| GO:0031331 | positive regulation of cellular catabolic process | *APOE, APOA4, PAFAH1B2* | 3/15 | $7.63\times10^{-4}$ | $5.02\times10^{-3}$ |
| GO:0010874 | regulation of cholesterol efflux | *APOE, APOA1* | 2/15 | $8.14\times10^{-4}$ | $5.29\times10^{-3}$ |
| GO:0042743 | hydrogen peroxide metabolic process | *HP, APOA4* | 2/15 | $8.44\times10^{-4}$ | $5.43\times10^{-3}$ |
| GO:1901617 | organic hydroxy compound biosynthetic process | *APOE, HMGCR, APOA1* | 3/15 | $9.67\times10^{-4}$ | $6.14\times10^{-3}$ |
| GO:0042158 | lipoprotein biosynthetic process | *APOE, APOA1* | 2/15 | $1.00\times10^{-3}$ | $6.31\times10^{-3}$ |
| GO:0009636 | response to toxic substance | *APOE, HP, APOA4* | 3/15 | $1.14\times10^{-3}$ | $7.09\times10^{-3}$ |
| GO:0046503 | glycerolipid catabolic process | *LIPG, APOA4* | 2/15 | $1.21\times10^{-3}$ | $7.47\times10^{-3}$ |
| GO:0051056 | regulation of small GTPase mediated signal transduction | *APOE, DOCK6, APOA1* | 3/15 | $1.54\times10^{-3}$ | $9.39\times10^{-3}$ |
| GO:0051347 | positive regulation of transferase activity | *APOE, APOA4, APOA1* | 3/15 | $1.60\times10^{-3}$ | $9.57\times10^{-3}$ |
| GO:0050709 | negative regulation of protein secretion | *APOE, HMGCR* | 2/15 | $1.61\times10^{-3}$ | $9.57\times10^{-3}$ |
| GO:0046470 | phosphatidylcholine metabolic process | *APOA4, APOA1* | 2/15 | $1.65\times10^{-3}$ | $9.71\times10^{-3}$ |
| GO:0019216 | regulation of lipid metabolic process | *APOE, APOA4, APOA1* | 3/15 | $2.28\times10^{-3}$ | 0.01 |
| GO:0062012 | regulation of small molecule metabolic process | *APOE, APOA4, APOA1* | 3/15 | $2.36\times10^{-3}$ | 0.01 |
| GO:0046889 | positive regulation of lipid biosynthetic process | *APOE, APOA4* | 2/15 | $2.44\times10^{-3}$ | 0.01 |
| GO:0019915 | lipid storage | *APOE, APOA1* | 2/15 | $2.60\times10^{-3}$ | 0.01 |
| GO:0006644 | phospholipid metabolic process | *LIPG, APOA4, APOA1* | 3/15 | $2.89\times10^{-3}$ | 0.02 |
| GO:0019218 | regulation of steroid metabolic process | *APOE, APOA1* | 2/15 | $3.33\times10^{-3}$ | 0.02 |
| GO:0060291 | long-term synaptic potentiation | *APOE, HMGCR* | 2/15 | $3.33\times10^{-3}$ | 0.02 |
| GO:0022600 | digestive system process | *APOA4, APOA1* | 2/15 | $3.44\times10^{-3}$ | 0.02 |
| GO:0006979 | response to oxidative stress | *APOE, HP, APOA4* | 3/15 | $3.62\times10^{-3}$ | 0.02 |
| GO:0051224 | negative regulation of protein transport | *APOE, HMGCR* | 2/15 | $4.01\times10^{-3}$ | 0.02 |
| GO:1904950 | negative regulation of establishment of protein localization | *APOE, HMGCR* | 2/15 | $5.03\times10^{-3}$ | 0.03 |
| GO:0043603 | amide metabolic process | *APOE, PCSK7, HMGCR* | 3/15 | $5.56\times10^{-3}$ | 0.03 |
| GO:0007586 | digestion | *APOA4, APOA1* | 2/15 | $5.62\times10^{-3}$ | 0.03 |
| GO:0007264 | small GTPase-mediated signal transduction | *APOE, DOCK6, APOA1* | 3/15 | $6.20\times10^{-3}$ | 0.03 |
| GO:1903531 | negative regulation of secretion by cell | *APOE, HMGCR* | 2/15 | $6.40\times10^{-3}$ | 0.03 |
| GO:0045807 | positive regulation of endocytosis | *APOE, APOA1* | 2/15 | $6.81\times10^{-3}$ | 0.03 |
| GO:0006633 | fatty acid biosynthetic process | *LIPG, APOA4* | 2/15 | $7.32\times10^{-3}$ | 0.04 |
| GO:0014012 | peripheral nervous system axon regeneration | *APOA4* | 1/15 | $7.87\times10^{-3}$ | 0.04 |
| GO:0018158 | protein oxidation | *APOA1* | 1/15 | $7.87\times10^{-3}$ | 0.04 |
| GO:0038060 | nitric oxide-cGMP-mediated signaling | *APOE* | 1/15 | $7.87\times10^{-3}$ | 0.04 |
| GO:0045541 | negative regulation of cholesterol biosynthetic process | *APOE* | 1/15 | $7.87\times10^{-3}$ | 0.04 |
| GO:0097113 | AMPA glutamate receptor clustering | *APOE* | 1/15 | $7.87\times10^{-3}$ | 0.04 |
| GO:0097688 | glutamate receptor clustering | *APOE* | 1/15 | $7.87\times10^{-3}$ | 0.04 |

| | | | | | |
|---|---|---|---|---|---|
| GO:0106119 | negative regulation of sterol biosynthetic process | *APOE* | 1/15 | $7.87\times10^{-3}$ | 0.04 |
| GO:2000822 | regulation of behavioral fear response | *APOE* | 1/15 | $7.87\times10^{-3}$ | 0.04 |
| GO:0051048 | negative regulation of secretion | *APOE, HMGCR* | 2/15 | $8.20\times10^{-3}$ | 0.04 |
| GO:0050806 | positive regulation of synaptic transmission | *APOE, HMGCR* | 2/15 | $8.29\times10^{-3}$ | 0.04 |
| GO:1902905 | positive regulation of supramolecular fiber organization | *APOE, APOA1* | 2/15 | $8.38\times10^{-3}$ | 0.04 |
| GO:0010982 | regulation of high-density lipoprotein particle clearance | *LIPG* | 1/15 | $8.66\times10^{-3}$ | 0.04 |
| GO:0010986 | positive regulation of lipoprotein particle clearance | *LIPG* | 1/15 | $8.66\times10^{-3}$ | 0.04 |
| GO:0034115 | negative regulation of heterotypic cell-cell adhesion | *APOA1* | 1/15 | $8.66\times10^{-3}$ | 0.04 |
| GO:0070587 | regulation of cell-cell adhesion involved in gastrulation | *APOA1* | 1/15 | $8.66\times10^{-3}$ | 0.04 |
| GO:0090206 | negative regulation of cholesterol metabolic process | *APOE* | 1/15 | $8.66\times10^{-3}$ | 0.04 |
| GO:1900222 | negative regulation of amyloid-beta clearance | *HMGCR* | 1/15 | $8.66\times10^{-3}$ | 0.04 |
| GO:1903365 | regulation of fear response | *APOE* | 1/15 | $8.66\times10^{-3}$ | 0.04 |
| GO:0043534 | blood vessel endothelial cell migration | *APOE, APOA1* | 2/15 | $8.84\times10^{-3}$ | 0.04 |
| GO:0017038 | protein import | *APOE* | 1/15 | $9.44\times10^{-3}$ | 0.04 |
| GO:0070586 | cell-cell adhesion involved in gastrulation | *APOA1* | 1/15 | $9.44\times10^{-3}$ | 0.04 |
| GO:0097090 | presynaptic membrane organization | *APOE* | 1/15 | $9.44\times10^{-3}$ | 0.04 |
| GO:2000644 | regulation of receptor catabolic process | *APOE* | 1/15 | $9.44\times10^{-3}$ | 0.04 |
| GO:0046890 | regulation of lipid biosynthetic process | *APOE, APOA4* | 2/15 | 0.01 | 0.04 |
| GO:0010896 | regulation of triglyceride catabolic process | *APOA4* | 1/15 | 0.01 | 0.04 |
| GO:0038203 | TORC2 signaling | *SIK3* | 1/15 | 0.01 | 0.04 |
| GO:0097475 | motor neuron migration | *CELSR2* | 1/15 | 0.01 | 0.04 |
| GO:0001935 | endothelial cell proliferation | *APOE, APOA1* | 2/15 | 0.01 | 0.04 |
| GO:0050728 | negative regulation of inflammatory response | *APOE, APOA1* | 2/15 | 0.01 | 0.04 |
| GO:0007008 | outer mitochondrial membrane organization | *TOMM40* | 1/15 | 0.01 | 0.04 |
| GO:0045040 | protein insertion into mitochondrial outer membrane | *TOMM40* | 1/15 | 0.01 | 0.04 |
| GO:0051004 | regulation of lipoprotein lipase activity | *APOA4* | 1/15 | 0.01 | 0.04 |
| GO:0060354 | negative regulation of cell adhesion molecule production | *APOA1* | 1/15 | 0.01 | 0.04 |
| GO:0061000 | negative regulation of dendritic spine development | *APOE* | 1/15 | 0.01 | 0.04 |
| GO:1900272 | negative regulation of long-term synaptic potentiation | *APOE* | 1/15 | 0.01 | 0.04 |
| GO:1902950 | regulation of dendritic spine maintenance | *APOE* | 1/15 | 0.01 | 0.04 |
| GO:1905809 | negative regulation of synapse organization | *APOE* | 1/15 | 0.01 | 0.04 |
| GO:1905907 | negative regulation of amyloid fibril formation | *APOE* | 1/15 | 0.01 | 0.04 |
| GO:0034374 | low-density lipoprotein particle remodeling | *APOE* | 1/15 | 0.01 | 0.05 |
| GO:0035641 | locomotory exploration behavior | *APOE* | 1/15 | 0.01 | 0.05 |

| | | | | | |
|---|---|---|---|---|---|
| GO:0048167 | regulation of synaptic plasticity | *APOE, HMGCR* | 2/15 | 0.01 | 0.05 |
| GO:1903828 | negative regulation of protein localization | *APOE, HMGCR* | 2/15 | 0.01 | 0.05 |
| GO:0072330 | monocarboxylic acid biosynthetic process | *LIPG, APOA4* | 2/15 | 0.01 | 0.05 |
| GO:0043117 | positive regulation of vascular permeability | *APOE* | 1/15 | 0.01 | 0.05 |
| GO:0050746 | regulation of lipoprotein metabolic process | *LIPG* | 1/15 | 0.01 | 0.05 |
| GO:0051044 | positive regulation of membrane protein ectodomain proteolysis | *APOE* | 1/15 | 0.01 | 0.05 |
| GO:0055089 | fatty acid homeostasis | *APOE* | 1/15 | 0.01 | 0.05 |
| GO:0090209 | negative regulation of triglyceride metabolic process | *APOE* | 1/15 | 0.01 | 0.05 |
| GO:0010310 | regulation of hydrogen peroxide metabolic process | *HP* | 1/15 | 0.01 | 0.05 |
| GO:0010544 | negative regulation of platelet activation | *APOE* | 1/15 | 0.01 | 0.05 |
| GO:0015936 | coenzyme A metabolic process | *HMGCR* | 1/15 | 0.01 | 0.05 |
| GO:0042159 | lipoprotein catabolic process | *APOE* | 1/15 | 0.01 | 0.05 |
| GO:0072578 | neurotransmitter-gated ion channel clustering | *APOE* | 1/15 | 0.01 | 0.05 |
| GO:0090083 | regulation of inclusion body assembly | *APOE* | 1/15 | 0.01 | 0.05 |
| GO:1905906 | regulation of amyloid fibril formation | *APOE* | 1/15 | 0.01 | 0.05 |
| GO:0044242 | cellular lipid catabolic process | *LIPG, APOA4* | 2/15 | 0.01 | 0.05 |
| GO:0016358 | dendrite development | *CELSR2, APOE* | 2/15 | 0.01 | 0.05 |
| GO:1902931 | negative regulation of alcohol biosynthetic process | *APOE* | 1/15 | 0.01 | 0.05 |

(C) Significant gene-sets associated with HL from ORA2: KEGG

| **ID** | **Category: Subcategory: Description** | **Gene-set** | **GeneRatio** | **P value** | **FDR** |
|---|---|---|---|---|---|
| hsa04979 | Organismal Systems: Digestive system: Cholesterol metabolism | *LIPG, APOE, APOA4, APOA1* | 4/9 | $1.39\times10^{-7}$ | $2.22\times10^{-6}$ |
| hsa04977 | Organismal Systems: Digestive system: Vitamin digestion and absorption | *LIPG, APOE, APOA4, APOA1* | 2/9 | $3.17\times10^{-4}$ | $2.53\times10^{-3}$ |
| hsa05143 | Human Diseases: Infectious disease: parasitic: African trypanosomiasis | *LIPG, APOE, APOA4, APOA1* | 2/9 | $6.45\times10^{-4}$ | $3.44\times10^{-3}$ |
| hsa04975 | Organismal Systems: Digestive system: Fat digestion and absorption | LIPG/APOE/APOA4/APOA1 | 2/9 | $8.71\times10^{-4}$ | $3.49\times10^{-3}$ |

(D) Significant gene-sets associated with FL from ORA1: GO

| ID | Description | Gene-set | GeneRatio | P value | FDR |
|---|---|---|---|---|---|
| GO:0007008 | outer mitochondrial membrane organization | *SAMM50* | 1/2 | $1.47\times10^{-3}$ | 0.02 |
| GO:0045040 | protein insertion into mitochondrial outer membrane | *SAMM50* | 1/2 | $1.47\times10^{-3}$ | 0.02 |
| GO:0050872 | white fat cell differentiation | *PNPLA3* | 1/2 | $1.79\times10^{-3}$ | 0.02 |
| GO:0042407 | cristae formation | *SAMM50* | 1/2 | $2.11\times10^{-3}$ | 0.02 |
| GO:0019433 | triglyceride catabolic process | *PNPLA3* | 1/2 | $2.95\times10^{-3}$ | 0.02 |
| GO:0006654 | phosphatidic acid biosynthetic process | *PNPLA3* | 1/2 | $3.58\times10^{-3}$ | 0.02 |
| GO:0090151 | establishment of protein localization to mitochondrial membrane | *SAMM50* | 1/2 | $3.58\times10^{-3}$ | 0.02 |
| GO:0046473 | phosphatidic acid metabolic process | *PNPLA3* | 1/2 | $4.00\times10^{-3}$ | 0.02 |
| GO:0034389 | lipid droplet organization | *PNPLA3* | 1/2 | $4.10\times10^{-3}$ | 0.02 |
| GO:0046461 | neutral lipid catabolic process | *PNPLA3* | 1/2 | $4.10\times10^{-3}$ | 0.02 |
| GO:0046464 | acylglycerol catabolic process | *PNPLA3* | 1/2 | $4.10\times10^{-3}$ | 0.02 |
| GO:0019432 | triglyceride biosynthetic process | *PNPLA3* | 1/2 | $4.63\times10^{-3}$ | 0.02 |
| GO:0007007 | inner mitochondrial membrane organization | *SAMM50* | 1/2 | $4.74\times10^{-3}$ | 0.02 |
| GO:0046460 | neutral lipid biosynthetic process | *PNPLA3* | 1/2 | $5.58\times10^{-3}$ | 0.02 |
| GO:0046463 | acylglycerol biosynthetic process | *PNPLA3* | 1/2 | $5.58\times10^{-3}$ | 0.02 |
| GO:0046503 | glycerolipid catabolic process | *PNPLA3* | 1/2 | $6.94\times10^{-3}$ | 0.02 |
| GO:0071230 | cellular response to amino acid stimulus | *PNPLA3* | 1/2 | $9.56\times10^{-3}$ | 0.03 |
| GO:0071229 | cellular response to acid chemical | *PNPLA3* | 1/2 | 0.01 | 0.03 |
| GO:0033108 | mitochondrial respiratory chain complex assembly | *SAMM50* | 1/2 | 0.01 | 0.03 |
| GO:0006626 | protein targeting to mitochondrion | *SAMM50* | 1/2 | 0.01 | 0.03 |
| GO:0006641 | triglyceride metabolic process | *PNPLA3* | 1/2 | 0.01 | 0.03 |
| GO:0001676 | long-chain fatty acid metabolic process | *PNPLA3* | 1/2 | 0.01 | 0.03 |
| GO:0007006 | mitochondrial membrane organization | *SAMM50* | 1/2 | 0.01 | 0.03 |
| GO:0072655 | establishment of protein localization to mitochondrion | *SAMM50* | 1/2 | 0.01 | 0.03 |
| GO:0043200 | response to amino acid | *PNPLA3* | 1/2 | 0.01 | 0.03 |
| GO:0070585 | protein localization to mitochondrion | *SAMM50* | 1/2 | 0.01 | 0.03 |
| GO:0006639 | acylglycerol metabolic process | *PNPLA3* | 1/2 | 0.01 | 0.03 |
| GO:0006638 | neutral lipid metabolic process | *PNPLA3* | 1/2 | 0.01 | 0.03 |
| GO:0001101 | response to acid chemical | *PNPLA3* | 1/2 | 0.02 | 0.03 |
| GO:0055088 | lipid homeostasis | *PNPLA3* | 1/2 | 0.02 | 0.03 |
| GO:0006839 | mitochondrial transport | *SAMM50* | 1/2 | 0.02 | 0.03 |

| | | | | | |
|---|---|---|---|---|---|
| GO:0046474 | glycerophospholipid biosynthetic process | *PNPLA3* | 1/2 | 0.02 | 0.03 |
| GO:0032869 | cellular response to insulin stimulus | *PNPLA3* | 1/2 | 0.02 | 0.03 |
| GO:0044242 | cellular lipid catabolic process | *PNPLA3* | 1/2 | 0.03 | 0.03 |
| GO:0045017 | glycerolipid biosynthetic process | *PNPLA3* | 1/2 | 0.03 | 0.03 |
| GO:0008654 | phospholipid biosynthetic process | *PNPLA3* | 1/2 | 0.03 | 0.03 |
| GO:0009743 | response to carbohydrate | *PNPLA3* | 1/2 | 0.03 | 0.03 |
| GO:0045444 | fat cell differentiation | *PNPLA3* | 1/2 | 0.03 | 0.03 |
| GO:0032868 | response to insulin | *PNPLA3* | 1/2 | 0.03 | 0.04 |
| GO:0090150 | establishment of protein localization to membrane | *SAMM50* | 1/2 | 0.03 | 0.04 |
| GO:0006650 | glycerophospholipid metabolic process | *PNPLA3* | 1/2 | 0.03 | 0.04 |
| GO:0071375 | cellular response to peptide hormone stimulus | *PNPLA3* | 1/2 | 0.03 | 0.04 |
| GO:0006605 | protein targeting | *SAMM50* | 1/2 | 0.03 | 0.04 |
| GO:0016042 | lipid catabolic process | *PNPLA3* | 1/2 | 0.04 | 0.04 |
| GO:0006644 | phospholipid metabolic process | *PNPLA3* | 1/2 | 0.04 | 0.04 |
| GO:0046486 | glycerolipid metabolic process | *PNPLA3* | 1/2 | 0.04 | 0.04 |
| GO:0006631 | fatty acid metabolic process | *PNPLA3* | 1/2 | 0.04 | 0.04 |
| GO:0043434 | response to peptide hormone | *PNPLA3* | 1/2 | 0.05 | 0.05 |
| GO:0007005 | mitochondrion organization | *SAMM50* | 1/2 | 0.05 | 0.05 |
| GO:0072594 | establishment of protein localization to organelle | *SAMM50* | 1/2 | 0.05 | 0.05 |

(E) Significant gene-sets associated with FL from ORA2: KEGG

| **ID** | **Category: Subcategory: Description** | **Gene-set** | **GeneRatio** | **P value** | **FDR** |
|---|---|---|---|---|---|
| hsa00561 | Metabolism: Lipid metabolism: Glycerolipid metabolism | *PNPLA3* | 1/2 | 0.02 | 0.02 |
| hsa04137 | Cellular Processes: Transport and catabolism: Mitophagy | *SAMM50* | 1/2 | 0.02 | 0.02 |

Note: HT, hypertension; HL, hyperlipidemia

**Table S11.** Distribution of obesity and obesity-related diseases in samples used for PRS analyses

| TRAIT | Validation dataset | | | Test dataset | | |
|---|---|---|---|---|---|---|
| | **Prev. (%)** | **Control** | **Case** | **Prev. (%)** | **Control** | **Case** |
| BMI25 | 32.44 % | 10,132 | 4864 | 33.48 % | 46,411 | 23,355 |
| BMI30 | 2.94 % | 14,555 | 441 | 2.97 % | 67,696 | 2070 |
| WC1 | 25.15 % | 10,274 | 3453 | 25.56 % | 48,672 | 16,714 |
| WC2 | 3.72 % | 13,217 | 510 | 3.67 % | 62,985 | 2401 |
| HT | 22.65 % | 9769 | 2860 | 22.31 % | 47,122 | 13,529 |
| T2D | 7.3 % | 12,454 | 981 | 7.84 % | 58,953 | 5015 |
| CVD | 4.31 % | 11,878 | 535 | 4.68 % | 56,935 | 2793 |
| HP | 12.36 % | 10,992 | 1550 | 11.31 % | 53,432 | 6816 |
| FL | 5.7 % | 10,711 | 648 | 5.8 % | 46,742 | 2877 |

Note: BMI25, BMI ≥ 25 kg/m²; BMI30, BMI ≥ 30 kg/m²; WC1, WC ≥ 85 cm for females / 90 cm for males; WC2, WC ≥ 95 cm for females / 100 cm for males; HT, hypertension; T2D, type 2 diabetes; CVD, cardiovascular disease; HL, hyperlipidemia; FL, fatty liver.

**Table S12. Heritability analyses and genetic correlations for PRS target traits in EAS populations**

(A) SNP heritability estimates and genetic correlations in EAS populations

| | SBP | DBP | BS | TG | HDL | BMI | WC |
|---|---|---|---|---|---|---|---|
| | **0.10** ($1.88\times10^{-23}$) | **0.93** (0) | **0.26** ($1.12\times10^{-5}$) | **0.25** ($1.11\times10^{-18}$) | **-0.09** ($1.11\times10^{-7}$) | 0.29 (0.12) | **0.33** ($1.80\times10^{-8}$) |
| **SBP** | **0.08** ($2.03\times10^{-30}$) | **0.86** (0) | **0.21** ($9.21\times10^{-4}$) | **0.15** ($2.00\times10^{-3}$) | -0.11 (0.29) | **0.28** ($1.24\times10^{-14}$) | **-** |
| | **0.14** ($1.90\times10^{-46}$) | **0.86** (0) | **0.18** ($1.33\times10^{-5}$) | **0.23** ($9.69\times10^{-13}$) | **-0.20** ($1.37\times10^{-7}$) | **0.28** ($1.21\times10^{-12}$) | **0.32** ($3.96\times10^{-12}$) |
| | | **0.09** ($1.04\times10^{-25}$) | 0.12 (0.06) | **0.20** ($1.64\times10^{-5}$) | -0.09 (0.10) | **0.23** ($4.24\times10^{-7}$) | **0.28** ($8.39\times10^{-8}$) |
| **DBP** | | **0.06** ($1.24\times10^{-22}$) | 0.13 (0.09) | 0.07 (0.14) | -0.01 (0.94) | **0.28** ($2.21\times10^{-14}$) | **-** |
| | | **0.14** ($7.65\times10^{-53}$) | **0.10** (0.02) | **0.21** ($7.66\times10^{-9}$) | **-0.19** ($1.06\times10^{-6}$) | **0.26** ($2.30\times10^{-10}$) | **0.29** ($1.10\times10^{-11}$) |
| | | | **0.12** ($1.11\times10^{-18}$) | 0.16 (0.06) | **-0.20** ($4.00\times10^{-3}$) | **0.20** ($3.57\times10^{-5}$) | **0.31** ($7.72\times10^{-9}$) |
| **BS** | | | **0.05** ($1.56\times10^{-12}$) | 0.07 (0.31) | -0.08 (0.44) | **0.09** (0.08) | **-** |
| | | | **0.15** ($8.81\times10^{-18}$) | **0.24** ($4.00\times10^{-4}$) | **-0.20** ($1.00\times10^{-4}$) | **0.22** ($3.39\times10^{-6}$) | **0.26** ($7.26\times10^{-7}$) |
| | | | | **0.16** ($1.91\times10^{-7}$) | **-0.61** ($2.10\times10^{-16}$) | **0.21** ($4.31\times10^{-6}$) | **0.27** ($4.87\times10^{-8}$) |
| **TG** | | | | **0.12** ($4.16\times10^{-5}$) | **-0.72** ($1.10\times10^{-8}$) | **0.20** ($2.75\times10^{-4}$) | **-** |
| | | | | **0.20** ($3.50\times10^{-7}$) | **-0.61** ($9.00\times10^{-18}$) | **0.26** ($2.76\times10^{-6}$) | **0.27** ($1.21\times10^{-6}$) |
| | | | | | **0.17** ($8.70\times10^{-15}$) | **-0.23** ($1.22\times10^{-7}$) | **-0.27** ($6.70\times10^{-7}$) |
| **HDL** | | | | | **0.16** ($1.28\times10^{-8}$) | **-0.27** ($1.88\times10^{-3}$) | **-** |
| | | | | | **0.25** ($6.04\times10^{-19}$) | **-0.40** ($1.66\times10^{-12}$) | **-0.39** ($1.09\times10^{-10}$) |
| | | | | | | **0.21** ($1.07\times10^{-62}$) | **0.86** (0) |
| **BMI** | | | | | | **0.17** ($1.07\times10^{-101}$) | **-** |
| | | | | | | **0.22** ($1.69\times10^{-99}$) | **0.88** (0) |
| | | | | | | | **0.15** ($3.66\times10^{-46}$) |
| **WC** | | | | | | | **-** |
| | | | | | | | **0.16** ($1.12\times10^{-69}$) |

(B) Estimated genetic correlations between Japanese/Taiwanese and Korean populations

| Trait | SBP | DBP | BS | TG | HDL | BMI | WC |
|---|---|---|---|---|---|---|---|

| | | | | | | | |
|---|---|---|---|---|---|---|---|
| $GC_{BBJ\text{-}KOR}$ | **0.99** (P = $3.37 \times 10^{-68}$) | **1.02** (P = $2.77 \times 10^{-48}$) | **0.89** (P = $2.88 \times 10^{-30}$) | **0.90** (P = $1.69 \times 10^{-109}$) | **0.95** (P = $5.25 \times 10^{-56}$) | **0.90** (P = 0) | - |
| $GC_{TWB\text{-}KOR}$ | **0.99** (P = $3.35 \times 10^{-65}$) | **0.95** (P = $1.94 \times 10^{-66}$) | **1.00** (P = $1.52 \times 10^{-90}$) | **0.91** (P = $7.76 \times 10^{-115}$) | **0.89** (P = $3.37 \times 10^{-85}$) | **0.92** (P = $1.23 \times 10^{-243}$) | **0.94** (P = $3.59 \times 10^{-128}$) |

Note: SBP, systolic blood pressure; DBP, diastolic blood pressure; BS, blood sugar; TG, triglycerides; HDL, high-density lipoprotein cholesterol; BMI, body mass index; WC, waist circumference; BBJ, Biobank Japan; TWB, Taiwan Biobank. In table (A), diagonal elements denote SNP heritability estimates ($h^2_{KOR}$, $h^2_{BBJ}$ and $h^2_{TWB}$) and upper diagonal elements denote genetic correlations ($GC_{KOR}$, $GC_{BBJ}$, and $GC_{TWB}$) estimated from Korean, Japanese (BBJ), and Taiwanese (TWB) populations. In table (B), $GC_{BBJ\text{-}KOR}$ and $GC_{TWB\text{-}KOR}$ represent the genetic correlations estimated between BBJ/TWB and Korean population.

**Table S13.** Validation results of single trait PRS analyses for obesity

| Methods | General obesity | | | | | Abdominal obesity | | | | |
|---|---|---|---|---|---|---|---|---|---|---|
| | COR | P | AIC | AUC | | COR | P | AIC | AUC | |
| | | | | BMI25 | BMI30 | | | | WC1 | WC2 |
| CT | 0.138 | $1.91{\times}10^{-67}$ | 31434.34 | 0.620 | 0.605 | 0.076 | $5.47{\times}10^{-24}$ | 56565.70 | 0.614 | 0.572 |
| LDpred: infinitesimal | 0.221 | $5.52{\times}10^{-173}$ | 30948.80 | 0.645 | 0.672 | 0.128 | $8.93{\times}10^{-75}$ | 56332.75 | 0.631 | 0.619 |
| LDpred: grid1 ($\rho$ = 1%) | 0.227 | $3.25{\times}10^{-185}$ | 30892.49 | 0.648 | 0.683 | 0.132 | $1.75{\times}10^{-77}$ | 56320.30 | 0.632 | 0.629 |
| LDpred: grid2 ($\rho$ = 3%) | 0.238 | $2.10{\times}10^{-202}$ | 30813.35 | 0.652 | 0.687 | 0.140 | $1.24{\times}10^{-86}$ | 56278.24 | 0.634 | 0.630 |
| LDpred: grid3 ($\rho$ = 10%) | 0.235 | $2.17{\times}10^{-196}$ | 30841.04 | 0.650 | 0.683 | 0.135 | $1.99{\times}10^{-82}$ | 56297.57 | 0.633 | 0.625 |
| LDpred: grid4 ($\rho$ = 30%) | 0.226 | $1.94{\times}10^{-181}$ | 30909.87 | 0.646 | 0.676 | 0.130 | $6.78{\times}10^{-77}$ | 56323.00 | 0.631 | 0.621 |
| LDpred: grid5 ($\rho$ = 100%) | 0.220 | $6.84{\times}10^{-172}$ | 30953.83 | 0.644 | 0.671 | 0.128 | $9.38{\times}10^{-75}$ | 56332.85 | 0.631 | 0.619 |
| **LDpred: auto** | **0.241** | $\mathbf{4.43{\times}10^{-207}}$ | **30791.81** | **0.653** | **0.687** | **0.140** | $\mathbf{2.04{\times}10^{-87}}$ | **56274.63** | **0.634** | **0.633** |
| lassosum | 0.236 | $8.82{\times}10^{-197}$ | 30839.23 | 0.650 | 0.682 | 0.137 | $3.41{\times}10^{-85}$ | 56284.85 | 0.633 | 0.626 |
| PRS-CS | 0.225 | $5.66{\times}10^{-181}$ | 30912.02 | 0.648 | 0.675 | 0.128 | $2.62{\times}10^{-76}$ | 56325.70 | 0.630 | 0.622 |

Note: BMI25, BMI ≥ 25 kg/m²; BMI30, BMI ≥ 30 kg/m²; BMI40, BMI ≥ 40 kg/m²; WC1, WC ≥ 85 cm for females / 90 cm for males; WC2, WC ≥ 95 cm for females / 100 cm for males; CT, clumping and thresholding; COR, Pearson correlation between BMI and the PRS; P, P value for PRS association; AIC, Akaike Information Criterion from linear models for BMI, adjusted for age, sex, PRS, and the first five principal components (PC1–5); AUC, mean area under the receiver operating characteristic curve obtained from 10-fold cross-validated logistic regression models for obesity outcomes, adjusted for age, sex, PRS, and PC1–5.

**Table S14. Validation results of each single trait PRS analyses**

(A) PRS validation results for SBP

| Methods | COR | P | AIC |
|---|---|---|---|
| CT | 0.086 | $3.51\times10^{-26}$ | 74058.11 |
| LDpred: infinitesimal | 0.112 | $8.40\times10^{-43}$ | 73982.00 |
| LDpred: grid1 ($\rho$ = 1%) | 0.128 | $2.04\times10^{-56}$ | 73919.52 |
| LDpred: grid2 ($\rho$ = 3%) | 0.128 | $4.69\times10^{-56}$ | 73921.18 |
| LDpred: grid4 ($\rho$ = 10%) | 0.119 | $9.70\times10^{-49}$ | 73954.76 |
| LDpred: grid3 ($\rho$ = 30%) | 0.115 | $4.40\times10^{-45}$ | 73971.54 |
| LDpred: grid5 ($\rho$ = 100%) | 0.112 | $8.63\times10^{-43}$ | 73982.05 |
| **LDpred: auto** | **0.132** | $\mathbf{1.31\times10^{-59}}$ | **73904.86** |
| lassosum | 0.129 | $1.04\times10^{-56}$ | 73918.17 |
| PRS-CS | 0.113 | $5.83\times10^{-44}$ | 73976.68 |

(B) PRS validation results for DBP

| Methods | COR | P | AIC |
|---|---|---|---|
| CT | 0.074 | $8.87\times10^{-20}$ | 62949.91 |
| LDpred: infinitesimal | 0.107 | $5.15\times10^{-39}$ | 62861.94 |
| LDpred: grid1 ($\rho$ = 1%) | 0.123 | $7.49\times10^{-51}$ | 62807.64 |
| LDpred: grid2 ($\rho$ = 3%) | 0.119 | $3.31\times10^{-48}$ | 62819.78 |
| LDpred: grid3 ($\rho$ = 10%) | 0.111 | $3.16\times10^{-42}$ | 62847.21 |
| LDpred: grid4 ($\rho$ = 30%) | 0.108 | $1.81\times10^{-39}$ | 62859.86 |
| LDpred: grid5 ($\rho$ = 100%) | 0.107 | $4.58\times10^{-39}$ | 62861.70 |
| **LDpred: auto** | **0.125** | $\mathbf{7.27\times10^{-53}}$ | **62798.40** |
| lassosum | 0.114 | $3.75\times10^{-44}$ | 62838.38 |
| PRS-CS | 0.104 | $1.97\times10^{-36}$ | 62873.78 |

(C) PRS validation results for BS

| Methods | COR | P | AIC |
|---|---|---|---|
| CT | 0.105 | $6.53\times10^{-37}$ | 79360.22 |
| LDpred: infinitesimal | 0.055 | $2.53\times10^{-13}$ | 79467.89 |
| LDpred: grid1 ($\rho$ = 1%) | 0.103 | $2.93\times10^{-38}$ | 79354.05 |
| LDpred: grid2 ($\rho$ = 3%) | 0.085 | $2.06\times10^{-27}$ | 79403.70 |
| LDpred: grid3 ($\rho$ = 10%) | 0.065 | $2.00\times10^{-17}$ | 79449.26 |
| LDpred: grid4 ($\rho$ = 30%) | 0.058 | $2.43\times10^{-14}$ | 79463.28 |
| LDpred: grid5 ($\rho$ = 100%) | 0.055 | $2.46\times10^{-13}$ | 79467.83 |
| LDpred: auto | 0.112 | $2.20\times10^{-44}$ | 79325.97 |
| **lassosum** | **0.124** | $\mathbf{2.81\times10^{-51}}$ | **79294.33** |
| PRS-CS | 0.061 | $2.36\times10^{-15}$ | 79458.68 |

(D) PRS validation results for TG

| Methods | COR | P | AIC |
|---|---|---|---|

| | | | |
|---|---|---|---|
| CT | 0.184 | $4.07\times10^{-109}$ | 122473.10 |
| LDpred: infinitesimal | 0.131 | $7.46\times10^{-58}$ | 122708.70 |
| LDpred: grid1 ($\rho$ = 1%) | 0.238 | $3.84\times10^{-186}$ | 122118.50 |
| LDpred: grid2 ($\rho$ = 3%) | 0.226 | $2.30\times10^{-168}$ | 122200.40 |
| LDpred: grid3 ($\rho$ = 10%) | 0.199 | $1.81\times10^{-130}$ | 122374.80 |
| LDpred: grid4 ($\rho$ = 30%) | 0.166 | $6.28\times10^{-92}$ | 122552.10 |
| LDpred: grid5 ($\rho$ = 100%) | 0.130 | $4.57\times10^{-57}$ | 122712.40 |
| LDpred: auto | 0.239 | $1.01\times10^{-188}$ | 122106.70 |
| **lassosum** | **0.246** | $\mathbf{6.07\times10^{-199}}$ | **122059.60** |
| PRS-CS | 0.168 | $1.44\times10^{-92}$ | 122549.20 |

(E) PRS validation results for HDL

| **Methods** | **COR** | **P** | **AIC** |
|---|---|---|---|
| CT | 0.255 | $2.57\times10^{-221}$ | 69083.89 |
| LDpred: infinitesimal | 0.163 | $3.98\times10^{-96}$ | 69660.17 |
| LDpred: grid1 ($\rho$ = 1%) | 0.301 | $2.77\times10^{-322}$ | 68618.75 |
| LDpred: grid2 ($\rho$ = 3%) | 0.287 | $2.12\times10^{-292}$ | 68756.47 |
| LDpred: grid3 ($\rho$ = 10%) | 0.242 | $1.23\times10^{-208}$ | 69142.28 |
| LDpred: grid4 ($\rho$ = 30%) | 0.195 | $1.01\times10^{-135}$ | 69477.98 |
| LDpred: grid5 ($\rho$ = 100%) | 0.163 | $1.06\times10^{-96}$ | 69657.53 |
| LDpred: auto | 0.278 | $5.22\times10^{-296}$ | 68864.22 |
| **lassosum** | **0.312** | **0** | **68545.46** |
| PRS-CS | 0.220 | $2.07\times10^{-171}$ | 69313.70 |

Note: SBP, systolic blood pressure; DBP, diastolic blood pressure; BS, blood sugar; TG, triglycerides; HDL, high-density lipoprotein cholesterol; CT, clumping and thresholding; COR, Pearson correlation between the target traits and the its PRS; P, P value for PRS association; AIC, Akaike Information Criterion from linear models for BMI, adjusted for age, sex, PRS, and the first five principal components.

**Table S15. Model performance improvement using single trait and multiple trait PRS analyses**

| Trait | AIC | | | | AUC | | | | | | |
|---|---|---|---|---|---|---|---|---|---|---|---|
| | **M1** | **M2** | **M3** | **M4** | **M1** | **M2** | **P: M1vs.M2** | **M3** | **P: M1/2vs.M3** | **M4** | **P: M3vs.M4** |
| BMI25 | 85801.97 | 86697.21 | 85454.59 | 85453.69 | 0.628 | 0.611 | **$1.64\times10^{-18}$** | 0.635 | **$1.22\times10^{-59}$** | 0.635 | 0.13 |
| BMI30 | 18106.06 | 18346.71 | 17995.36 | 17998.04 | 0.645 | 0.610 | **$7.76\times10^{-7}$** | 0.659 | **$5.42\times10^{-23}$** | 0.659 | 0.53 |
| WC1 | 72070.99 | 71991.22 | 71604.23 | 71603.43 | 0.623 | 0.626 | 0.23 | 0.634 | **$1.03\times10^{-17}$** | 0.635 | 0.08 |
| WC2 | 20031.98 | 20081.08 | 19893.26 | 19896.41 | 0.638 | 0.631 | 0.19 | 0.653 | **$2.50\times10^{-12}$** | 0.654 | 0.18 |
| HT | 59319.81 | 59319.34 | 59284.60 | 58473.65 | 0.698 | 0.698 | 0.77 | 0.698 | **0.02** | 0.712 | **$1.36\times10^{-44}$** |
| T2D | 33204.80 | 33136.06 | 33138.05 | 32831.37 | 0.679 | 0.682 | **$2.40\times10^{-4}$** | 0.682 | 0.92 | 0.697 | **$8.31\times10^{-22}$** |
| CVD | 20426.18 | 20419.98 | 20420.34 | 20416.28 | 0.742 | 0.742 | 0.49 | 0.742 | 0.41 | 0.743 | 0.06 |
| HL | 42038.24 | 42034.94 | 42036.52 | 41814.87 | 0.592 | 0.592 | 0.65 | 0.592 | 0.78 | 0.606 | **$7.11\times10^{-11}$** |
| FL | 21239.98 | 21229.98 | 21228.91 | 21216.84 | 0.653 | 0.653 | 0.69 | 0.654 | 0.55 | 0.657 | **0.03** |

Note: P, DeLong test P value for comparing AUC between models. Model descriptions — M1: adjusted for age, sex, body mass index (BMI) PRS, and the first five principle components (PC1–5); M2: adjusted for age, sex, waist circumference (WC) PRS, and PC1–5; M3: adjusted for age, sex, BMI PRS, WC PRS, and PC1–5; M4: adjusted for age, sex, PRSs for BMI, WC, systolic blood pressure (SBP), diastolic blood pressure (DBP), blood sugar (BS), triglycerides (TG), high-density lipoprotein cholesterol (HDL), and PC1–5.

**Table S16.** Validation results for obesity-related diseases: T2D and CVD

| Methods | T2D | | | CVD | | |
|---|---|---|---|---|---|---|
| | **P** | **AIC** | **AUC** | **P** | **AIC** | **AUC** |
| CT | $2.01\times10^{-38}$ | 6454.28 | 0.716 | $1.86\times10^{-3}$ | 4000.05 | 0.746 |
| LDpred: infinitesimal | $5.68\times10^{-36}$ | 6462.21 | 0.716 | $1.99\times10^{-6}$ | 3987.01 | 0.747 |
| LDpred: grid1 ($\rho$ = 1%) | $1.69\times10^{-57}$ | 6355.18 | 0.734 | $3.93\times10^{-7}$ | 3983.89 | 0.750 |
| LDpred: grid2 ($\rho$ = 3%) | $3.50\times10^{-55}$ | 6366.96 | 0.732 | $\mathbf{2.56\times10^{-7}}$ | **3983.04** | **0.749** |
| LDpred: grid4 ($\rho$ = 10%) | $5.67\times10^{-48}$ | 6403.10 | 0.726 | $5.37\times10^{-7}$ | 3984.45 | 0.748 |
| LDpred: grid3 ($\rho$ = 30%) | $1.00\times10^{-40}$ | 6439.01 | 0.719 | $1.82\times10^{-6}$ | 3986.82 | 0.747 |
| LDpred: grid5 ($\rho$ = 100%) | $6.04\times10^{-36}$ | 6462.34 | 0.715 | $2.29\times10^{-6}$ | 3987.28 | 0.747 |
| **LDpred: auto** | $\mathbf{1.05\times10^{-57}}$ | **6354.16** | **0.734** | $2.37\times10^{-6}$ | 3987.38 | 0.749 |
| lassosum | $6.73\times10^{-30}$ | 6491.62 | 0.709 | $4.27\times10^{-7}$ | 3983.99 | 0.749 |
| PRS-CS | $5.07\times10^{-46}$ | 6413.33 | 0.724 | $4.43\times10^{-6}$ | 3988.55 | 0.747 |

Note: T2D, type 2 diabetes; CVD, cardiovascular disease.

**Table S17.** Interaction and mediation effects of obesity on obesity-related diseases: T2D and CVD

(A) Interaction effect of obesity on obesity-related diseases

| **Outcome (Disease) PRS** | **Obesity PRS** | **Obesity** | **Interaction with disease PRS** | **Disease PRS** | | **Obesity PRS** | | **Obesity** | | **Interaction** | |
|---|---|---|---|---|---|---|---|---|---|---|---|
| | | | | **OR (CI)** | **P** | **OR (CI)** | **P** | **OR (CI)** | **P** | **OR (CI)** | **P** |
| T2D | BMI | BMI25 | BMI PRS | 1.57 (1.52,1.62) | **$3.12\times10^{-174}$** | 0.92 (0.89,0.96) | **$2.39\times10^{-6}$** | 2.04 (1.92,2.17) | **$5.63\times10^{-114}$** | 1.00 (0.97,1.04) | 0.79 |
| | | | BMI25 | 1.67 (1.60,1.74) | **$4.58\times10^{-116}$** | 0.93 (0.90,0.96) | **$1.21\times10^{-6}$** | 2.12 (1.99,2.26) | **$7.88\times10^{-115}$** | 0.88 (0.83,0.94) | **$9.90\times10^{-5}$** |
| | WC | WC1 | WC PRS | 1.56 (1.51,1.61) | **$5.27\times10^{-162}$** | 1.00 (0.97,1.03) | 0.93 | 2.21 (2.07,2.35) | **$1.32\times10^{-135}$** | 1.00 (0.97,1.03) | 0.95 |
| | | | WC1 | 1.63 (1.57,1.70) | **$9.83\times10^{-120}$** | 1.00 (0.97,0.13) | 0.93 | 2.29 (2.15,2.45) | **$6.29\times10^{-135}$** | 0.89 (0.83,0.95) | **$2.22\times10^{-4}$** |
| | PRSsum1 | BMI25 | PRSsum1 | 1.56 (1.52,1.62) | **$1.59\times10^{-167}$** | 0.96 (0.93,0.99) | **0.02** | 2.01 (1.89,2.14) | **$4.24\times10^{-110}$** | 1.00 (0.97,1.03) | 0.93 |
| | | | BMI25 | 1.66 (1.59,1.74) | **$1.16\times10^{-113}$** | 0.96 (0.93,0.99) | **0.01** | 2.10 (1.96,2.24) | **$3.56\times10^{-111}$** | 0.88 (0.83,0.94) | **$9.31\times10^{-5}$** |
| | PRSsum2 | WC1 | PRSsum2 | 1.56 (1.51,1.61) | **$3.85\times10^{-165}$** | 0.98 (0.95,1.02) | 0.33 | 2.21 (2.08,2.36) | **$6.09\times10^{-137}$** | 1.00 (0.97,1.03) | 0.80 |
| | | | WC1 | 1.64 (1.57,1.71) | **$2.82\times10^{-121}$** | 0.98 (0.95,1.01) | 0.26 | 2.30 (2.16,2.46) | **$3.73\times10^{-136}$** | 0.89 (0.83,0.95) | **$2.20\times10^{-4}$** |
| CVD | BMI | BMI25 | BMI PRS | 1.23 (1.18,1.28) | **$1.30\times10^{-24}$** | 1.01 (0.97,1.05) | 0.78 | 1.45 (1.34,1.57) | **$1.22\times10^{-19}$** | 1.02 (0.98,1.06) | 0.33 |
| | | | BMI25 | 1.24 (1.17,1.30) | **$3.71\times10^{-15}$** | 1.01 (0.97,1.05) | 0.66 | 1.45 (1.34,1.58) | **$2.42\times10^{-19}$** | 0.99 (0.91,1.07) | 0.79 |
| | WC | WC1 | WC PRS | 1.24 (1.19,1.29) | **$1.70\times10^{-24}$** | 1.03 (0.99,1.07) | 0.16 | 1.51 (1.38,1.64) | **$1.15\times10^{-21}$** | 1.02 (0.98,1.06) | 0.29 |
| | | | WC1 | 1.25 (1.19,1.32) | **$6.47\times10^{-18}$** | 1.03 (0.99,1.08) | 0.10 | 1.52 (1.39,1.65) | **$1.11\times10^{-21}$** | 0.97 (0.89,1.05) | 0.44 |
| | PRSsum1 | BMI25 | PRSsum1 | 1.23 (1.18,1.28) | **$3.69\times10^{-24}$** | 1.02 (0.98,1.06) | 0.28 | 1.44 (1.33,1.56) | **$5.68\times10^{-19}$** | 1.03 (0.99,1.07) | 0.14 |
| | | | BMI25 | 1.24 (1.17,1.30) | **$5.01\times10^{-15}$** | 1.03 (0.99,1.07) | 0.18 | 1.44 (1.33,1.57) | **$1.09\times10^{-18}$** | 0.99 (0.91,1.07) | 0.79 |
| | PRSsum2 | WC1 | PRSsum2 | 1.23 (1.18,1.28) | **$7.51\times10^{-24}$** | 1.05 (1.01,1.10) | **0.01** | 1.50 (1.38,1.63) | **$3.58\times10^{-21}$** | 1.01 (0.98,1.06) | 0.46 |
| | | | WC1 | 1.25 (1.19,1.31) | **$1.47\times10^{-17}$** | 1.06 (1.01,1.10) | **$7.81\times10^{-3}$** | 1.51 (1.38,1.64) | **$3.31\times10^{-21}$** | 0.97 (0.89,1.05) | 0.43 |

(B) Mediation effects of obesity on the associations between obesity PRS and obesity-related diseases

| Disease (Y) | Independent variable (X) | Mediator (M) | Mediator Group | Direct effect | | Indirect effect | | Prop. of med. effect | |
|---|---|---|---|---|---|---|---|---|---|
| | | | | BETA | P | BETA | P | BETA | P |
| T2D | BMI PRS | BMI25 | Case (obese) | -0.0009 | 0.47 | 0.0043 | **$<2\times10^{-16}$** | 1.2363 | **$<2\times10^{-16}$** |
| | | | Control (non-obese) | -0.0008 | 0.47 | 0.0043 | **$<2\times10^{-16}$** | 1.2479 | **$<2\times10^{-16}$** |
| | | | Average | -0.0008 | 0.47 | 0.0043 | **$<2\times10^{-16}$** | 1.2421 | **$<2\times10^{-16}$** |
| | WC PRS | WC1 | Case (obese) | 0.0059 | **$<2\times10^{-16}$** | 0.0039 | **$<2\times10^{-16}$** | 0.4049 | **$<2\times10^{-16}$** |
| | | | Control (non-obese) | 0.0057 | **$<2\times10^{-16}$** | 0.0036 | **$<2\times10^{-16}$** | 0.3799 | **$<2\times10^{-16}$** |
| | | | Average | 0.0058 | **$<2\times10^{-16}$** | 0.0037 | **$<2\times10^{-16}$** | 0.3924 | **$<2\times10^{-16}$** |
| | PRSsum1 | BMI25 | Case (obese) | 0.0039 | **$<2\times10^{-16}$** | 0.0047 | **$<2\times10^{-16}$** | 0.5597 | **$<2\times10^{-16}$** |
| | | | Control (non-obese) | 0.0037 | **$<2\times10^{-16}$** | 0.0046 | **$<2\times10^{-16}$** | 0.5373 | **$<2\times10^{-16}$** |
| | | | Average | 0.0038 | **$<2\times10^{-16}$** | 0.0046 | **$<2\times10^{-16}$** | 0.5485 | **$<2\times10^{-16}$** |
| | | WC1 | Case (obese) | 0.0037 | **$<2\times10^{-16}$** | 0.0043 | **$<2\times10^{-16}$** | 0.5474 | **$<2\times10^{-16}$** |
| | | | Control (non-obese) | 0.0036 | **$<2\times10^{-16}$** | 0.0041 | **$<2\times10^{-16}$** | 0.5251 | **$<2\times10^{-16}$** |
| | | | Average | 0.0037 | **$<2\times10^{-16}$** | 0.0042 | **$<2\times10^{-16}$** | 0.5363 | **$<2\times10^{-16}$** |
| | PRSsum2 | BMI25 | Case (obese) | 0.0047 | **$<2\times10^{-16}$** | 0.0041 | **$<2\times10^{-16}$** | 0.4764 | **$<2\times10^{-16}$** |
| | | | Control (non-obese) | 0.0045 | **$<2\times10^{-16}$** | 0.0039 | **$<2\times10^{-16}$** | 0.4533 | **$<2\times10^{-16}$** |
| | | | Average | 0.0046 | **$<2\times10^{-16}$** | 0.0040 | **$<2\times10^{-16}$** | 0.4648 | **$<2\times10^{-16}$** |
| | | WC1 | Case (obese) | 0.0044 | **$<2\times10^{-16}$** | 0.0037 | **$<2\times10^{-16}$** | 0.4684 | **$<2\times10^{-16}$** |
| | | | Control (non-obese) | 0.0042 | **$<2\times10^{-16}$** | 0.0035 | **$<2\times10^{-16}$** | 0.4463 | **$<2\times10^{-16}$** |
| | | | Average | 0.0043 | **$<2\times10^{-16}$** | 0.0036 | **$<2\times10^{-16}$** | 0.4573 | **$<2\times10^{-16}$** |
| CVD | BMI PRS | BMI25 | Case (obese) | 0.0006 | 0.47 | 0.0013 | **$<2\times10^{-16}$** | 0.6777 | **0.02** |
| | | | Control (non-obese) | 0.0006 | 0.47 | 0.0013 | **$<2\times10^{-16}$** | 0.6695 | **0.02** |
| | | | Average | 0.0006 | 0.47 | 0.0013 | **$<2\times10^{-16}$** | 0.6736 | **0.02** |
| | WC PRS | WC1 | Case (obese) | 0.0019 | **0.04** | 0.0010 | **$<2\times10^{-16}$** | 0.3640 | **$<2\times10^{-16}$** |
| | | | Control (non-obese) | 0.0018 | **0.04** | 0.0010 | **$<2\times10^{-16}$** | 0.3510 | **$<2\times10^{-16}$** |
| | | | Average | 0.0018 | **0.04** | 0.0010 | **$<2\times10^{-16}$** | 0.3580 | **$<2\times10^{-16}$** |
| | PRSsum1 | BMI25 | Case (obese) | 0.0016 | 0.06 | 0.0015 | **$<2\times10^{-16}$** | 0.4770 | **$<2\times10^{-16}$** |
| | | | Control (non-obese) | 0.0016 | 0.06 | 0.0014 | **$<2\times10^{-16}$** | 0.4630 | **$<2\times10^{-16}$** |

| | | | | | | | | | |
|---|---|---|---|---|---|---|---|---|---|
| | | | Average | 0.0016 | 0.06 | 0.0014 | **<2×10$^{-16}$** | 0.4700 | **<2×10$^{-16}$** |
| | | WC1 | Case (obese) | 0.0019 | **0.04** | 0.0013 | **<2×10$^{-16}$** | 0.4120 | **<2×10$^{-16}$** |
| | | | Control (non-obese) | 0.0018 | **0.04** | 0.0012 | **<2×10$^{-16}$** | 0.3967 | **<2×10$^{-16}$** |
| | | | Average | 0.0019 | **0.04** | 0.0013 | **<2×10$^{-16}$** | 0.4043 | **<2×10$^{-16}$** |
| | PRSsum2 | BMI25 | Case (obese) | 0.0030 | **2.0×10$^{-3}$** | 0.0012 | **<2×10$^{-16}$** | 0.2933 | **<2×10$^{-16}$** |
| | | | Control (non-obese) | 0.0029 | **2.0×10$^{-3}$** | 0.0012 | **<2×10$^{-16}$** | 0.2776 | **<2×10$^{-16}$** |
| | | | Average | 0.0029 | **2.0×10$^{-3}$** | 0.0012 | **<2×10$^{-16}$** | 0.2854 | **<2×10$^{-16}$** |
| | | WC1 | Case (obese) | 0.0030 | **<2×10$^{-16}$** | 0.0010 | **<2×10$^{-16}$** | 0.2599 | **<2×10$^{-16}$** |
| | | | Control (non-obese) | 0.0029 | **<2×10$^{-16}$** | 0.0010 | **<2×10$^{-16}$** | 0.2450 | **<2×10$^{-16}$** |
| | | | Average | 0.0029 | **<2×10$^{-16}$** | 0.0010 | **<2×10$^{-16}$** | 0.2525 | **<2×10$^{-16}$** |

Note: BMI, body mass index; BMI25, BMI ≥ 25 kg/m²; WC, waist circumference; WC1, WC ≥ 85 cm for females / 90 cm for males; T2D, type 2 diabetes; CVD, cardiovascular disease; PRSsum1, Sum of BMI and WC PRS; PRSsum2, Sum of BMI, WC, systolic blood pressure (SBP), diastolic blood pressure (DBP), blood sugar (BS), triglyceride (TG), and high-density lipoprotein (HDL) cholesterol PRSs; Prop. of med. effect, Proportion of mediation effect.

**Figure S1.** Manhattan plots constructed from GWAS results for obesity in the replication dataset of REP1$_{Kor}$

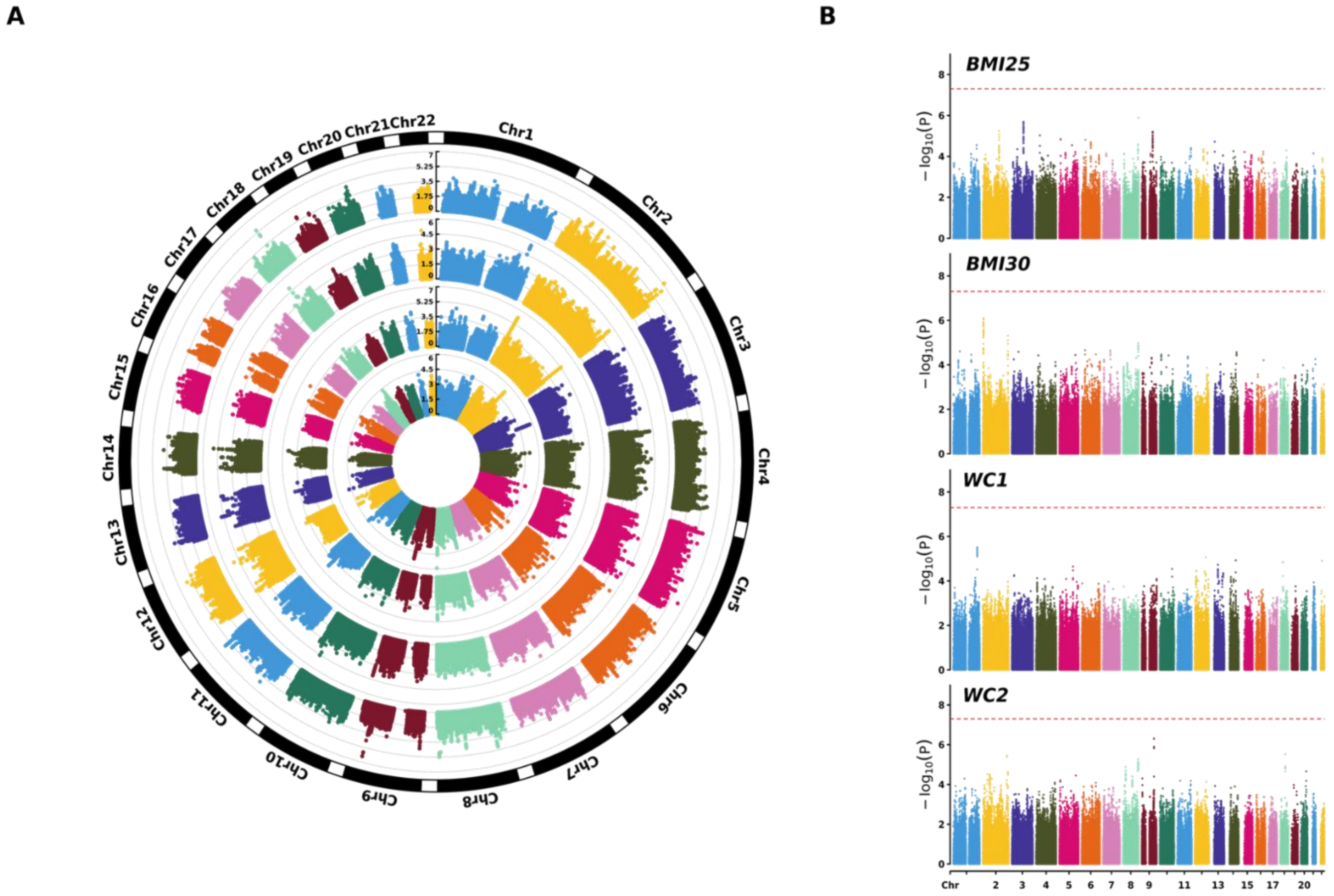


Circular and standard Manhattan plots are shown for four obesity phenotypes: BMI25 (body mass index ≥ 25 kg/m²), BMI30 (≥ 30 kg/m²), WC1 (waist circumference ≥ 85 cm for females / 90 cm for males), and WC2 (≥ 95 cm for females / 100 cm for males). The red dashed line indicates the genome-wide significance threshold. Panel A presents circular plots arranged from inner to outer rings. Panel B displays the corresponding standard Manhattan plots.

**Figure S2.** Manhattan plots constructed from GWAS results for obesity in the replication dataset of REP2$_{Chi}$

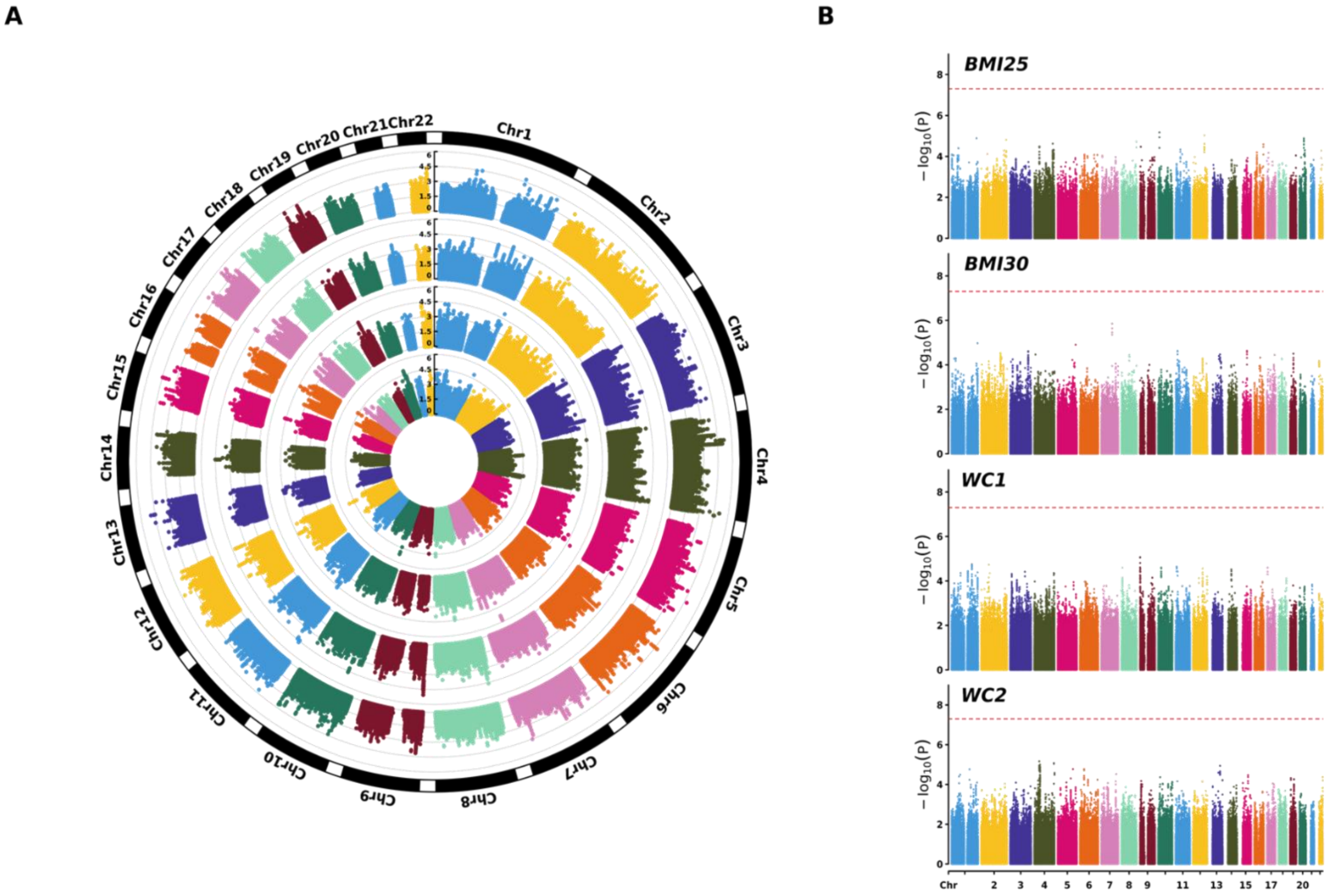


Circular and standard Manhattan plots are shown for four obesity phenotypes: BMI25 (body mass index ≥ 25 kg/m²), BMI30 (≥ 30 kg/m²), WC1 (waist circumference ≥ 85 cm for females / 90 cm for males), and WC2 (≥ 95 cm for females / 100 cm for males). The red dashed line indicates the genome-wide significance threshold. Panel A presents circular plots arranged from inner to outer rings. Panel B displays the corresponding standard Manhattan plots.

**Figure S3.** Manhattan plots constructed from GWAS results for obesity in the replication dataset of $REP3_{NHW}$

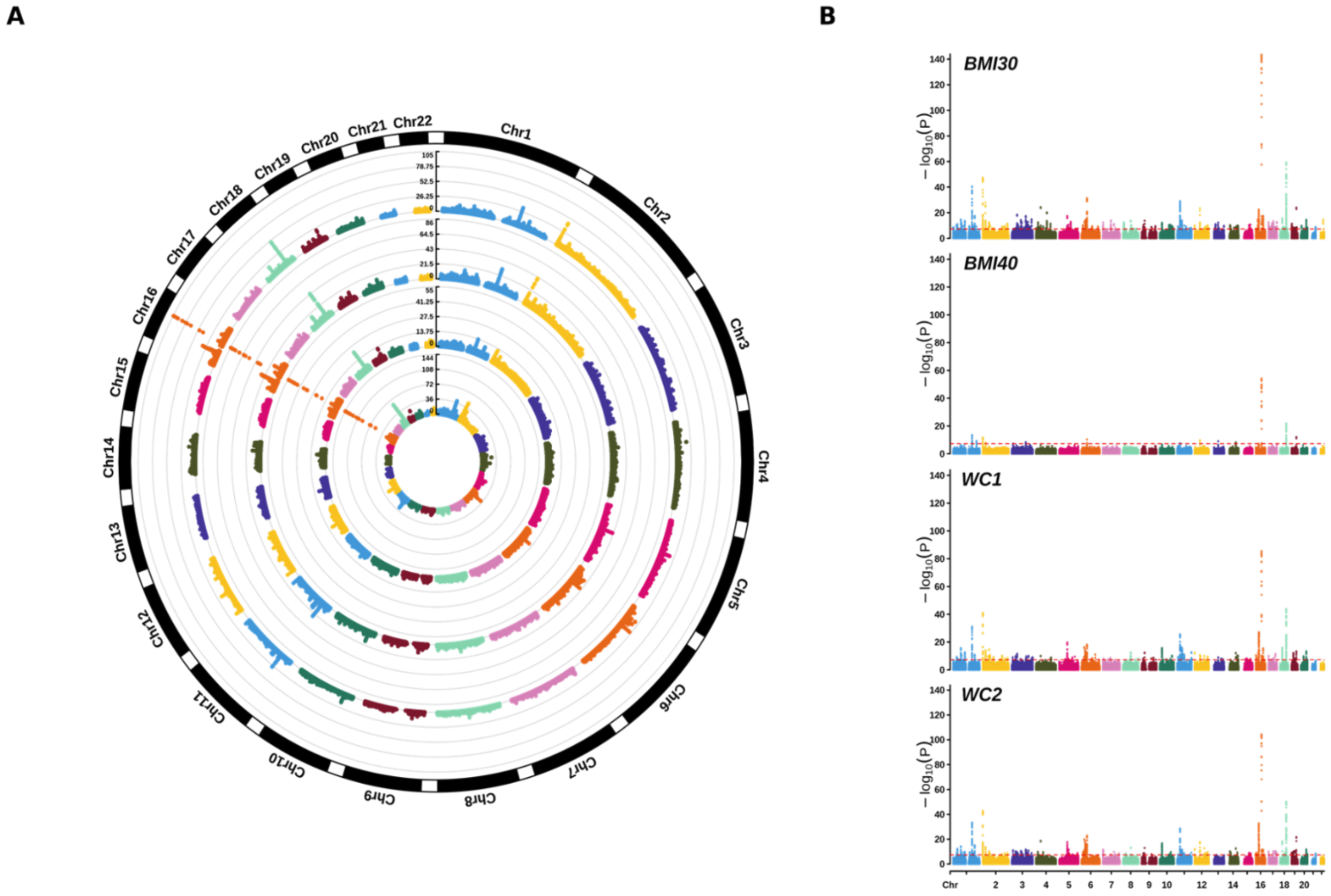


Circular and standard Manhattan plots are shown for four obesity phenotypes: BMI30 (body mass index ≥ 30 kg/m²), BMI40 (≥ 40 kg/m²), WC1 (waist circumference ≥ 85 cm for females / 90 cm for males), and WC2 (≥ 95 cm for females / 100 cm for males). The red dashed line indicates the genome-wide significance threshold. Panel A presents circular plots arranged from inner to outer rings. Panel B displays the corresponding standard Manhattan plots.

**Figure S4A.** Comparison of GWAS results for BMI and general obesity in the discovery dataset

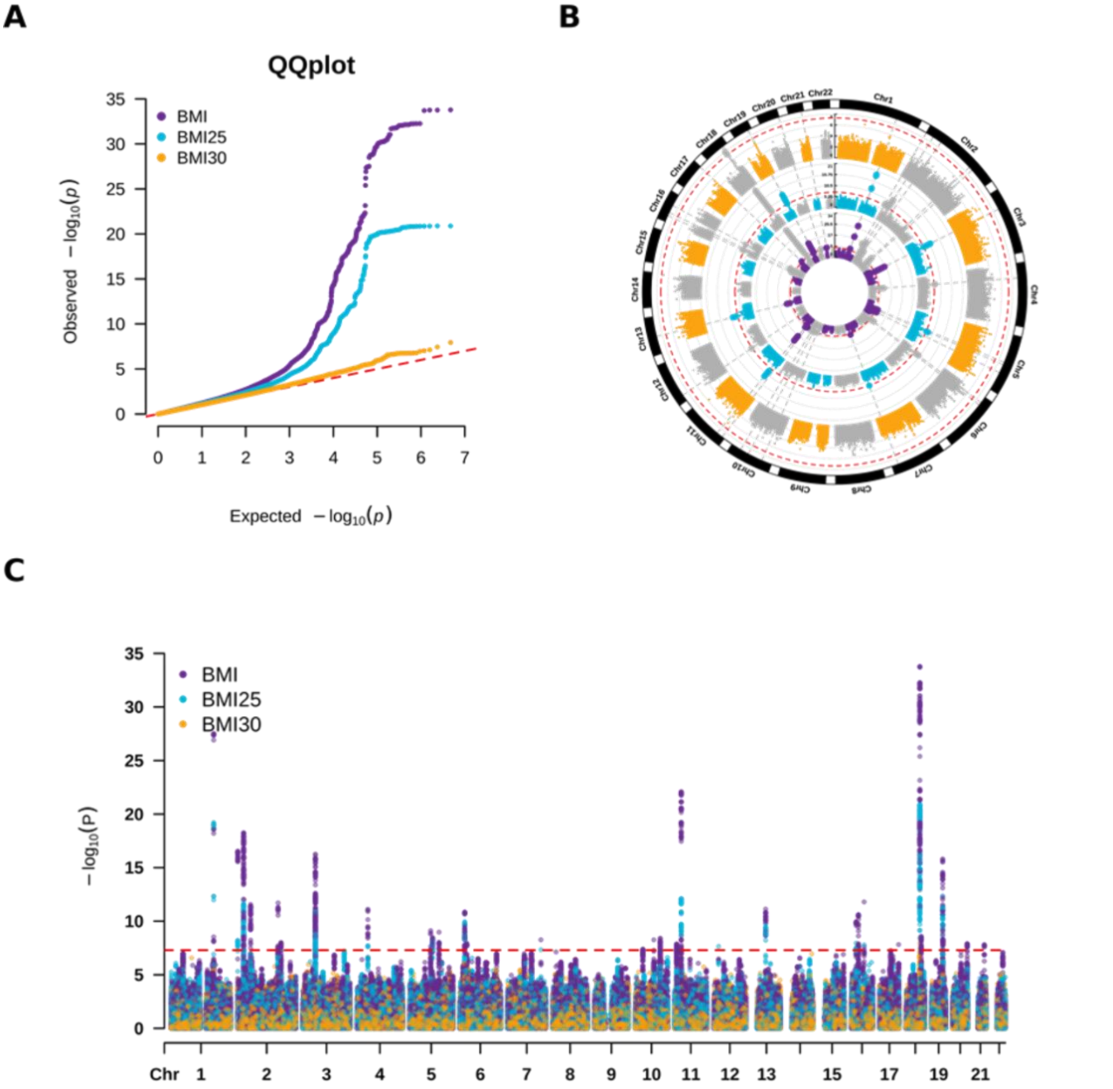


This figure compares genetic associations between continuous obesity-related indicators: BMI and dichotomous obesity definitions: BMI25 (body mass index ≥ 25 kg/m²), BMI30 (≥ 30 kg/m²). Panel A shows quantile–quantile (QQ) plots; Panel B presents circular Manhattan plots for BMI and two severity levels of general obesity; Panel C displays the corresponding standard Manhattan plots.

**Figure S4B.** Comparison of GWAS results for WC and abdominal obesity in the discovery dataset

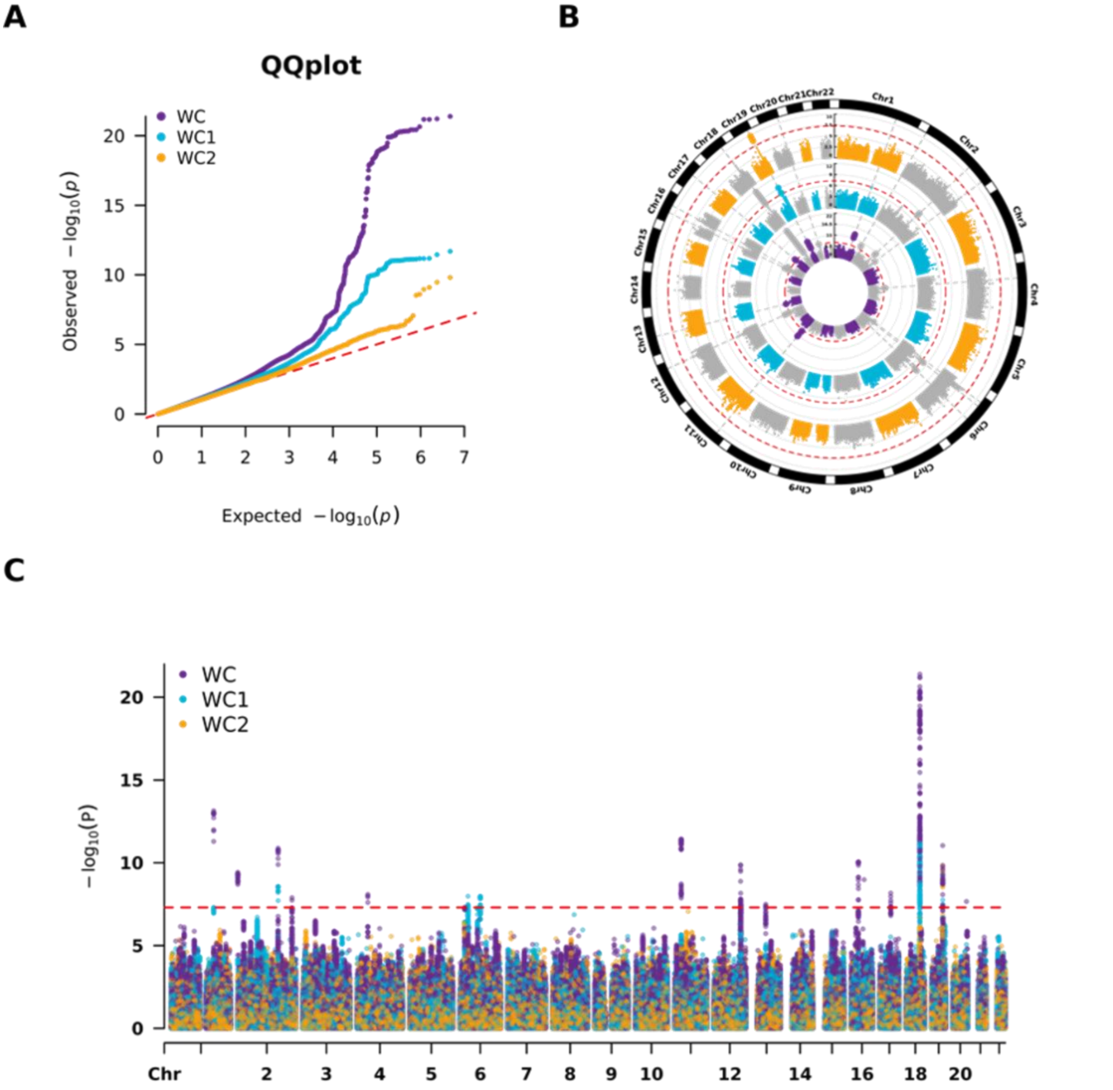


This figure compares genetic associations between continuous obesity-related indicators: WC and dichotomous obesity definitions: WC1 (waist circumference ≥ 85 cm for females / 90 cm for males) / WC2 (≥ 95 cm for females / 100 cm for males). Panel A shows quantile–quantile (QQ) plots; Panel B presents circular Manhattan plots for WC and two severity levels of abdominal obesity; Panel C displays the corresponding standard Manhattan plots.

**Figure S5A.** Manhattan plots from GWAS of obesity-related diseases in the discovery dataset

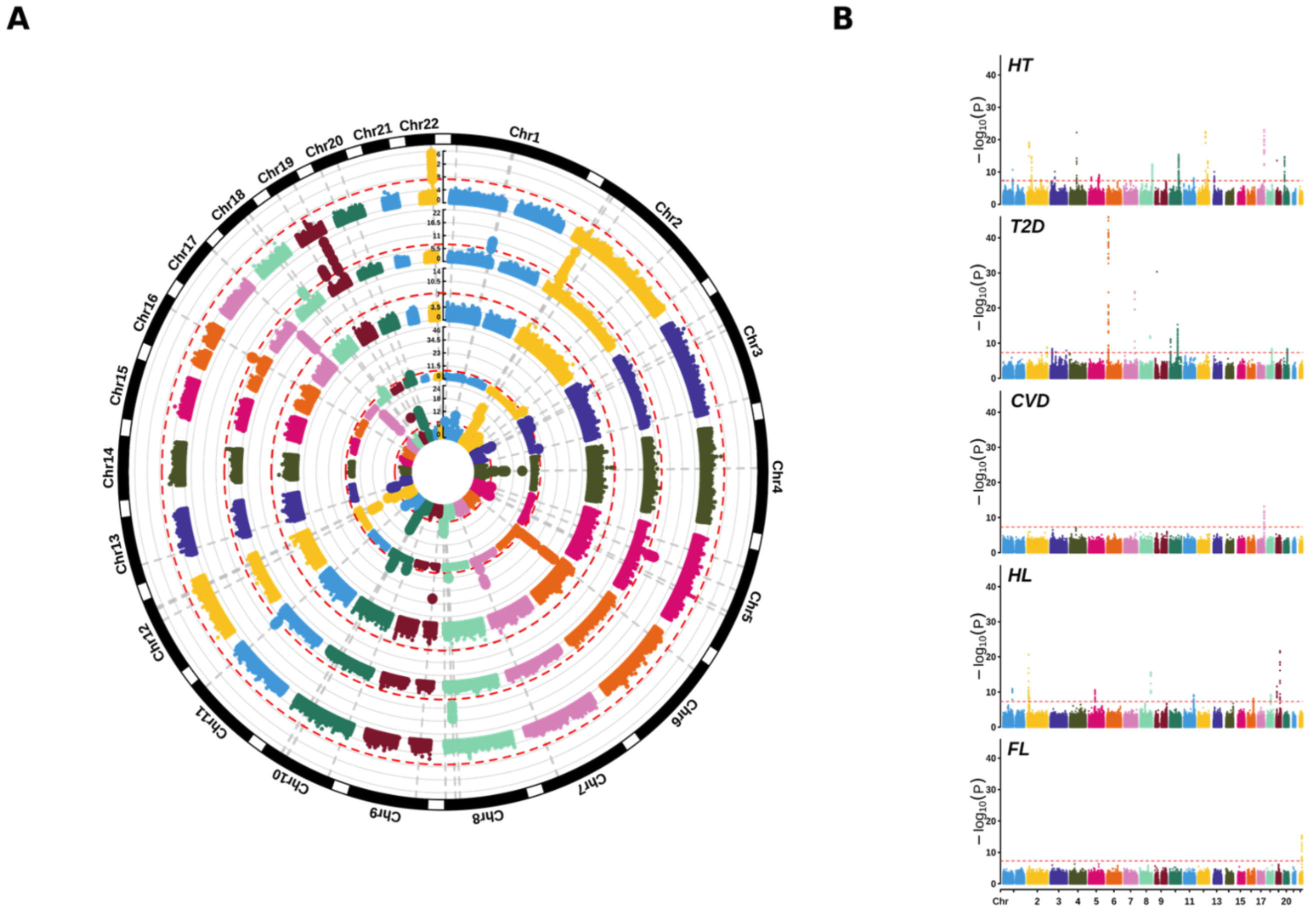


Circular and standard Manhattan plots are shown for five obesity-related disease phenotypes: hypertension (HT), type 2 diabetes (T2D), cardiovascular disease (CVD), hyperlipidemia (HL), and fatty liver (FL). The red dashed line indicates the genome-wide significance threshold. Panel A presents circular plots arranged from inner to outer rings. Panel B displays the corresponding standard Manhattan plots.

**Figure S5B.** Manhattan plots from GWAS of obesity-related diseases in the $\mathrm{REP3}_{\mathrm{NHW}}$ replication dataset

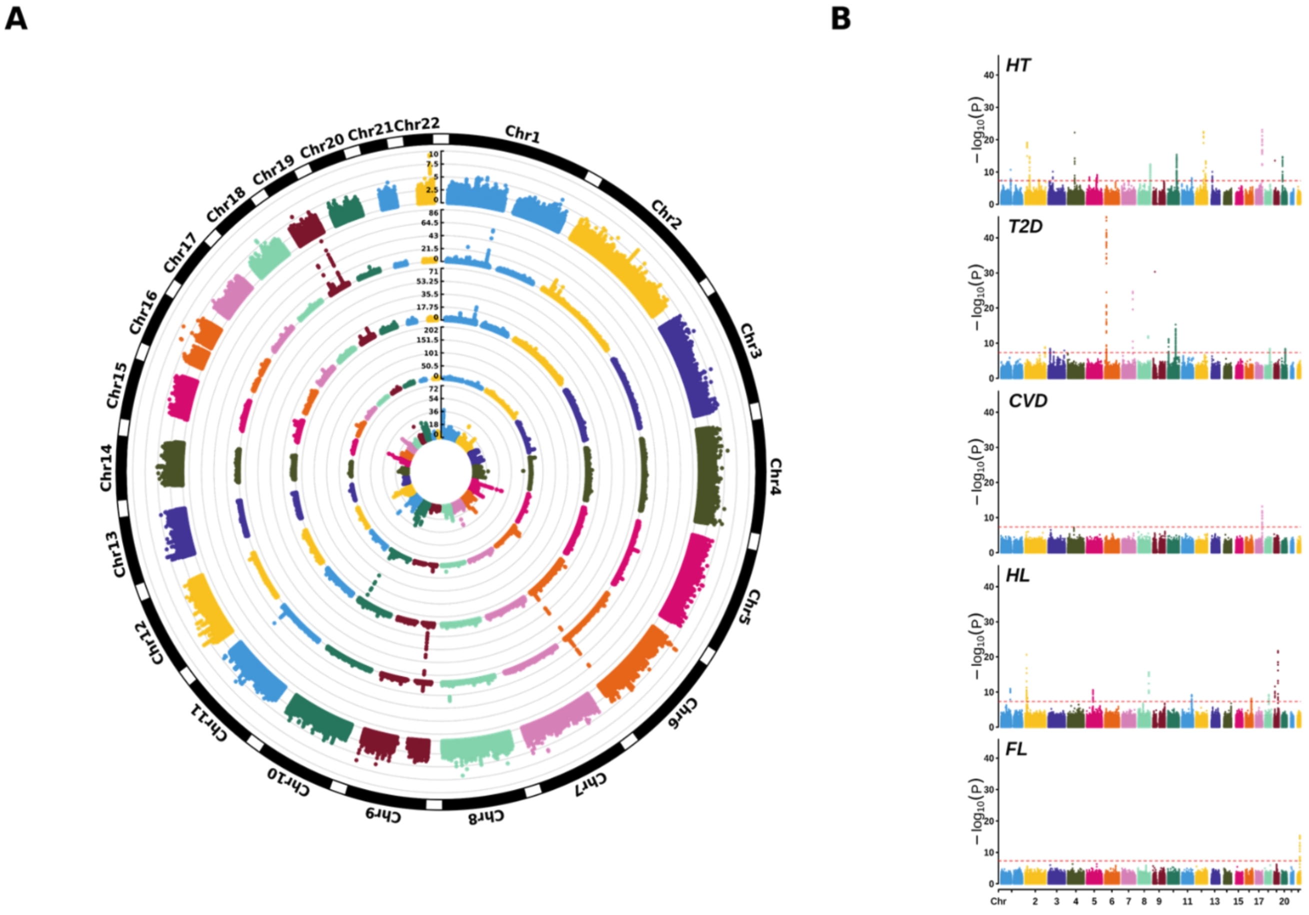


Circular and standard Manhattan plots are shown for five obesity-related disease phenotypes: hypertension (HT), type 2 diabetes (T2D), cardiovascular disease (CVD), hyperlipidemia (HL), and fatty liver (FL). The red dashed line indicates the genome-wide significance threshold. Panel A presents circular plots arranged from inner to outer rings. Panel B displays the corresponding standard Manhattan plots.

**Figure S6A.** Local genetic correlation between general obesity and obesity-related diseases

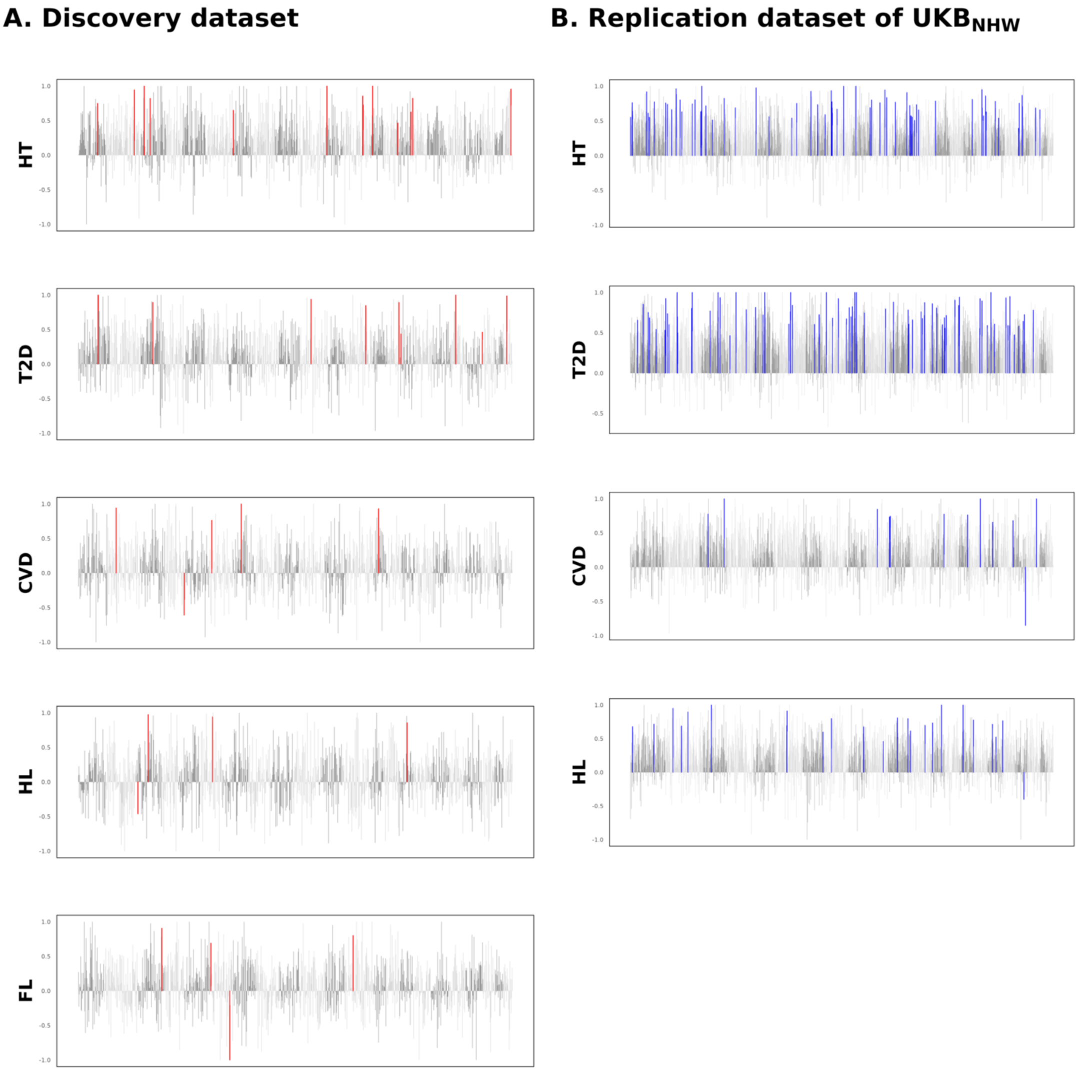


Local genetic correlation plots of general obesity (BMI25, body mass index ≥ 25 kg/m²) are shown for five obesity-related disease phenotypes: hypertension (HT), type 2 diabetes (T2D), cardiovascular disease (CVD), hyperlipidemia (HL), and fatty liver (FL). Panel A presents plots from the discovery dataset, with the red line indicating signals that meet the Bonferroni-corrected significance threshold. Panel B presents plots from the REP3$_{NHW}$ replication dataset, with the blue line indicating signals that surpass the same Bonferroni threshold.

**Figure S6B.** Local genetic correlation between abdominal obesity and obesity-related diseases

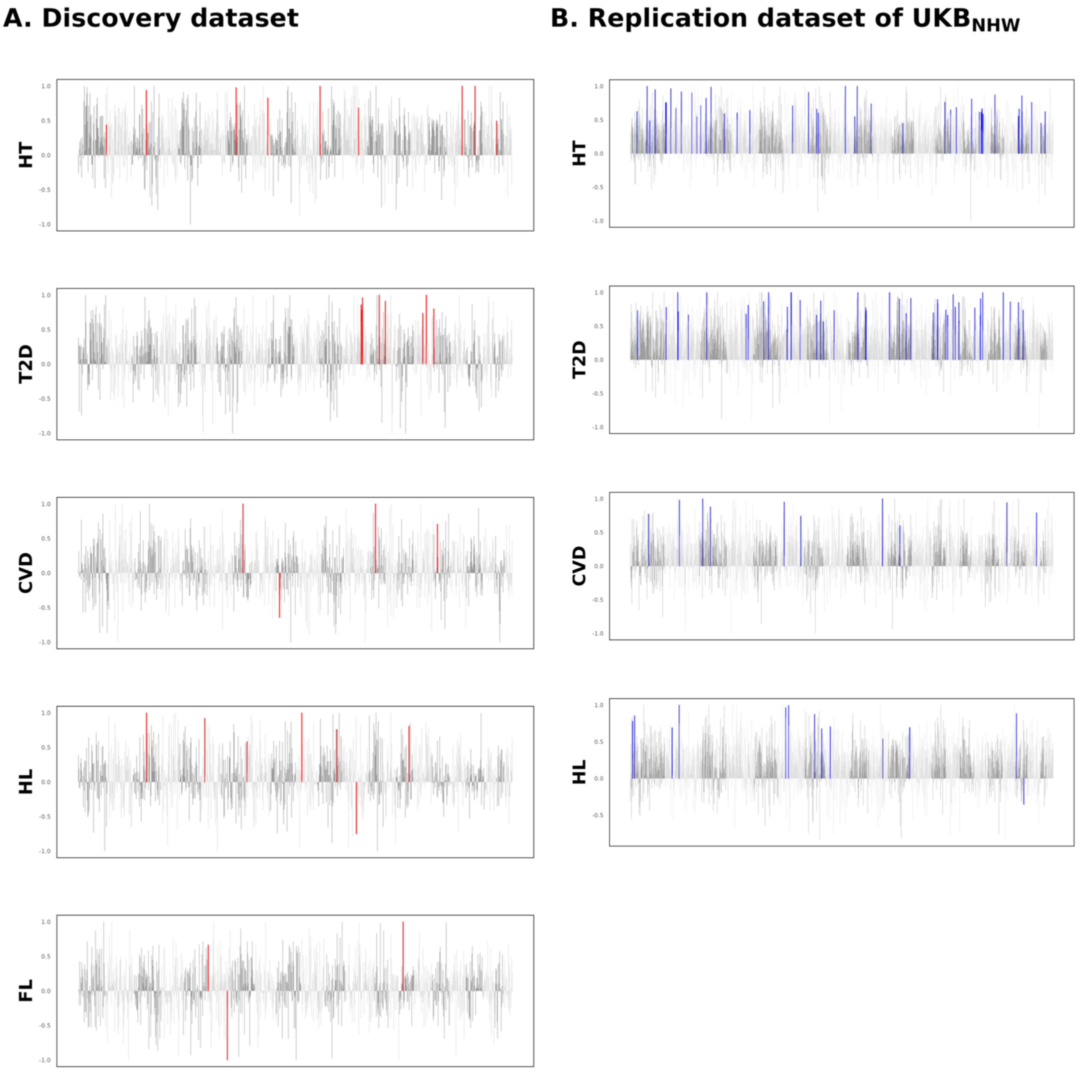


Local genetic correlation plots of abdominal obesity (WC1, waist circumference ≥ 85 cm for females / 90 cm for males) are shown for five obesity-related disease phenotypes: hypertension (HT), type 2 diabetes (T2D), cardiovascular disease (CVD), hyperlipidemia (HL), and fatty liver (FL). Panel A presents plots from the discovery dataset, with the red line indicating signals that meet the Bonferroni-corrected significance threshold. Panel B presents plots from the REP3$_{NHW}$ replication dataset, with the blue line indicating signals that surpass the same Bonferroni threshold.
